# Frequency of Variants in Mendelian Alzheimer’s Disease Genes within the Alzheimer’s Disease Sequencing Project (ADSP)

**DOI:** 10.1101/2023.10.24.23297227

**Authors:** Dongyu Wang, Alexandra Scalici, Yanbing Wang, Honghuang Lin, Achilleas Pitsillides, Nancy Heard-Costa, Carlos Cruchaga, Ellen Ziegemeier, Joshua C. Bis, Myriam Fornage, Eric Boerwinkle, Philip L De Jager, Ellen Wijsman, Josée Dupuis, Alan E. Renton, Sudha Seshadri, Alison M. Goate, Alzheimer’s Disease Neuroimaging Initiative (ADNI), The Alzheimer’s Disease Sequencing Project, Anita L. DeStefano, Gina M. Peloso

## Abstract

**Background:** Prior studies using the ADSP data examined variants within presenilin-2 (*PSEN2*), presenilin-1 (*PSEN1*), and amyloid precursor protein (*APP*) genes. However, previously-reported clinically-relevant variants and other predicted damaging missense (DM) variants have not been characterized in a newer release of the Alzheimer’s Disease Sequencing Project (ADSP).

**Objective:** To characterize previously-reported clinically-relevant variants and DM variants in *PSEN2, PSEN1, APP* within the participants from the ADSP.

**Methods:** We identified rare variants (MAF <1%) previously-reported in *PSEN2*, *PSEN1,* and *APP* in the available ADSP sample of 14,641 individuals with whole genome sequencing and 16,849 individuals with whole exome sequencing available for research-use (N_total_ = 31,490). We additionally curated variants in these three genes from ClinVar, OMIM, and Alzforum and report carriers of variants in clinical databases as well as predicted DM variants in these genes.

**Results:** We detected 31 previously-reported clinically-relevant variants with alternate alleles observed within the ADSP: 4 variants in *PSEN2*, 25 in *PSEN1*, and 2 in *APP*. The overall variant carrier rate for the 31 clinically-relevant variants in the ADSP was 0.3%. We observed that 79.5% of the variant carriers were cases compared to 3.9% were controls. In those with AD, the mean age of onset of AD among carriers of these clinically-relevant variants was 19.6 ± 1.4 years earlier compared with noncarriers (p-value=7.8×10^-57^).

**Conclusion:** A small proportion of individuals in the ADSP are carriers of a previously-reported clinically-relevant variant allele for AD and these participants have significantly earlier age of AD onset compared to noncarriers.

## INTRODUCTION

Mendelian Alzheimer’s disease (AD) makes up less than 1% of all AD cases and is characterized by an early age of onset (<65 years old) [1]. Rare mutations in the *PSEN1, PSEN2,* and *APP* genes have been previously characterized to show Mendelian AD inheritance autosomal dominant inheritance patterns within families with near complete penetrance [1]. Identification and classification of variants in these three genes has aided both in the molecular classification of pathways involved in AD pathogenesis and screening for known AD causing variants within families. Publicly available clinical databases such as ClinVar, Online Mendelian Inheritance of Man (OMIM), and Alzforum have compiled clinically-relevant variants presumed to cause AD and other types of dementia (“clinical variants”). These databases are important resources but also have limitations. For example, variants shown to be Mendelian with assumed near complete penetrance may also be found in late onset familial and sporadic AD cases, leading to conflicting interpretations of these variants due to biased ascertainment from the original highly-selected large pedigree studies [2,3].

The Alzheimer’s Disease Sequencing Project (ADSP) seeks to identify novel genetic risk factors for AD. The data collected and generated through the ADSP consist of whole genome sequence (WGS) and whole exome sequence (WES) data from family, case-control, and cohort study designs[4]. Within the ADSP, standardized variant calling and data management pipelines (VCPA), as well as an ADSP Quality Control (QC) protocol have been implemented[5,6]. Bringing together high-quality sequence data from across the AD research community has allowed for an increased sample size, overall increasing statistical power and the ability to search for rare variation associated with AD. Given that ADSP data are biased towards the selection of variants of incomplete penetrance, we aimed to leverage these data to further characterize previously-reported AD clinical variants.

Previously, two studies using a prior release of ADSP data examined variants within *APP*, *PSEN1*, *PSEN2*, and other dementia-related genes[7,8]. With a newer release and doubled sample size, we seek to examine the frequency of previously-reported clinically-relevant and conflicting clinically-relevant variants as well as predicted damaging missense (DM) variants in these three Mendelian AD genes within the ADSP WES and WGS datasets. We report on the distribution of these variants by racial and ethnic group to clarify the clinical implications of these previously-reported variants in the context of a history of disparity in representation in most studies of AD. Knowing the distribution of these variants in the ADSP datasets may also inform studies focused on gene and variant identification at other loci in the context of AD risk.

## METHODS

### ADSP study data

WES and WGS data have been generated in multiple cohorts as part of the ADSP. See **Supplementary Methods** for description of the data included in this manuscript. Study participants provided written informed consent per each study’s Institutional Review Board (IRB) approved protocol. These data were analyzed through a protocol approved by the Boston University IRB.

The release of the ADSP data used in the current study contains 16,905 samples with WGS data (NG00067.v3) and 20,504 samples with WES data (NG00067.v3). Genetically identical individuals were identified based on pairwise scaled KING kinship coefficient ≥ 0.354 as recommended by the PLINK2.0 documentation[9]. Keeping one genetically unique participant and preferentially selecting WGS over WES yielded 31,490 individuals (14,641 with WGS and 16,849 with WES) with both genotype and phenotype information. QC flags in the ADSP files were used in a filtering process to retain high quality variants as detailed in the supplemental materials.

### Phenotype determination

For participants in the ADSP case-control study, we defined AD cases as individuals with either prevalent or incident AD. ADSP case-control study participants with no prevalent or incident AD were defined as controls. Participants with a status of “NA” were recoded as “Unknown”. In the ADSP family studies, the AD status variable has the possible values of no dementia, definite AD, probable AD, possible AD, family-reported AD, other dementia, family-reported no dementia, and unknown. For ADSP family-based study participants, we defined AD cases as individuals coded with possible, probable, or definite AD. AD controls were defined as individuals coded as no dementia. Participants with a status of family-reported AD, other dementia, and unknown were all recoded as “Unknown” for AD status. The Alzheimer’s Disease Neuroimaging Initiative (ADNI) phenotype data, which is part of the ADSP augmentation study, provides information on mild cognitive impairment (MCI) in addition to AD status. Individuals with a current diagnosis of MCI were set to AD unknowns (N=313) in the current study. Age of onset was available in 98% (13,491 of 13,825) of the AD cases. Participants with age greater than 90 (“90+”) were recoded to 90 to create a continuous age variable. Individuals with reported race as Asian, Native Hawaiian or Pacific Islander, American Indian or Alaskan Native, Other, or Unknown were combined and classified as Other/Unknown (N=4,725).

### Curation of clinically-relevant variants

Variants previously-reported in *PSEN2*, *PSEN1*, *APP* were aggregated from ClinVar[10] (June 2023), Online Mendelian Inheritance in Man[11] (OMIM, June 2023), and Alzforum[12] (March, 2023), which we label as “clinical variants”. Variants were selected and classified as clinical variants if they were described as pathogenic or likely pathogenic, and related to AD, dementia, or a related disorder in any of the databases (**Supplementary Table e-1**). For variants with conflicting interpretations of pathogenicity in ClinVar, or having conflicting pathogenicity across the databases, we created a separate list and referred to the variants as “conflicting clinical variants”. Variants with minor allele frequency < 1% that met these criteria were selected within the WGS and WES using PLINK 2.0[13]. Variants were matched based on chromosome, position, reference allele, and alternate allele. Alternate allele is defined as the mutant allele as compared to the reference allele and individuals carrying an alternate allele of at least one clinical or conflicting clinical variant are termed clinical variant carriers or conflicting clinical variant carriers, respectively, in this study.

### Variant annotation

Variants in *PSEN2*, *PSEN1*, *APP* with minor allele frequency < 1% within ADSP were selected and annotated using Ensembl Variant Effect Predictor (VEP) release 108 with default parameter and human genome reference build 38[14]. Variants with missense consequence and a damaging metaSVM prediction from the database for non-synonymous functional prediction (dbNFSP) version 4.3a were classified as damaging missense (DM) variants[15,16]. Individuals who carry at least one DM variant that is not in the curated clinical variant list are classified as additional DM variant carriers. Additionally, we annotated the clinical variants within ADSP using the InterVar tool and obtained the gnomAD allele frequencies using the allele frequencies from the exomes control set as well as InterVer pathogenicity predictions[17]. We restricted to the control individuals for reporting the gnomAD allele frequencies given that ADSP contributed to gnomAD.

### Statistical analysis

The ADSP phenotype and genotype data were used to identify individuals carrying at least one alternate allele for a clinical variant or a DM variant and generate descriptive statistics based on carrier status and AD diagnosis. In order to examine the distribution of genetic variants across different populations, we defined population groups based on reported race and ethnicity. Principal components of ancestry were estimated using overlapping variants from WGS and WES with MAF > 0.1% and pairwise r^2^ < 0.1 in LD pruning. We compared the mean age of onset of AD by carrier status using a linear mixed effects regression model with carrier status as the exposure and age of onset as the outcome, adjusting for sex,the first four principal components of ancestry, an indicator variable for WGS or WES data, *APOE* ε*2*, and *APOE* ε*4* status, using an empirical Balding-Nichols kinship matrix. All statistical analyses were performed using R 4.2.1. A schematic of the analysis is shown in **Figure 1**.

**Figure 1.**
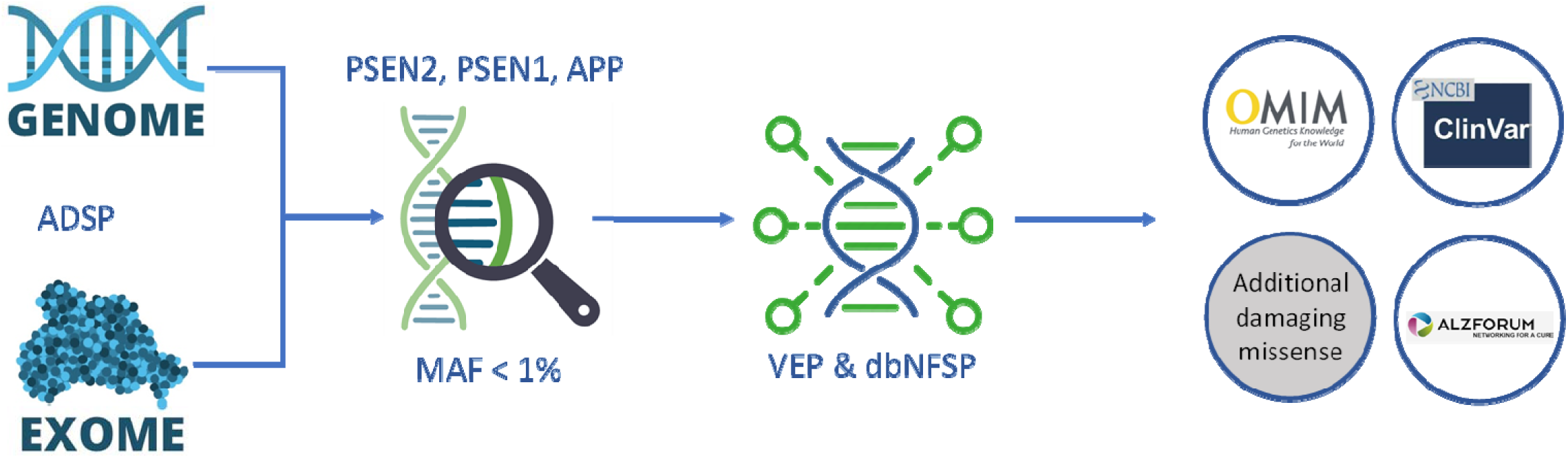
Flow Chart of the Analysis. Figure 1 represents the schematic of the analysis. ADSP whole genome sequencing data and whole exome sequencing data were combined, and variants with MAF<1% in *PSEN2*, *PENS1*, and *APP* genes were extracted. Subsequently, VEP and dbNFSP annotation was conducted to identify the additional damaging missense (DM) variants in the ADSP participants, followed by the curation of clinical variants from ClinVar, OMIM, and Alzforum databases. We then reported demographic and AD status of the carriers of these variants.

### Data availability

ADSP whole genome (NG00067.v3) and whole exome (NG00067.v3) sequencing data are available through The National Institute on Aging Genetics of Alzheimer’s Disease Data Storage Site (NIGADS) (https://www.niagads.org/).

## RESULTS

Aggregating variants in ClinVar, OMIM, and Alzforum that met our criteria yielded a list of 293 clinical variants and 51 conflicting clinical variants, totaling 344 variants: 29 in *PSEN2,* 251 in *PSEN1*, and 64 in *APP* (**Supplementary Table e-2**). After removing genetically identical individuals, we had a total of 31,490 individuals consisting of 13,825 AD cases, 14,715 controls, and 2,950 with unknown AD status (**Table 1**). We identified 78 individuals that carried at least one clinical variant, 550 individuals that carried at least one conflicting clinical variant, and 844 individuals that carried at least one additional DM variant (**Table 1**).

**Table 1.** Alzheimer Disease and Demographics by Carrier Status.

|  | **Clinical Variant Carriers** | **Conflicting Clinical Variant Carriers** | **Additional DM Variant Carriers** | **Total (%)** |
| --- | --- | --- | --- | --- |
| **N** | 78 | 550 | 844 | 31,490 |
| **AD Diagnosis** |  |  |  |  |
| Case | 62 (79.5%) | 224 (40.7%) | 365 (43.3%) | 13,825 (43.9%) |
| Control | 3 (3.9%) | 270 (49.1%) | 409 (48.5%) | 14,715 (46.7%) |
| Unknown | 13 (16.7%) | 56 (10.2%) | 70 (8.3%) | 2,950 (9.4%) |
| **Sex** |  |  |  |  |
| Male | 42 (53.9%) | 200 (36.4%) | 323 (38.3%) | 12,256 (38.9%) |
| Female | 36 (46.2%) | 350 (63.6%) | 521 (61.7%) | 19,234 (61.1%) |
| **Ethnicity/Race** |  |  |  |  |
| Non-Hispanic |  |  |  |  |
| White | 46 (59.0%) | 360 (65.5%) | 365 (43.3%) | 21,083 (67.0%) |
| Black | 3 (3.9%) | 95 (17.3%) | 295 (35.0%) | 5,286 (16.8%) |
| Other/Unknown | 0 (0%) | 0 (0%) | 4 (0.47%) | 102 (0.3%) |
| Hispanic |  |  |  |  |
| White | 12 (15.4%) | 6 (1.1%) | 7 (0.8%) | 322 (1.0%) |
| Black | 1 (1.3%) | 1 (0.2%) | 6 (0.7%) | 74 (0.2%) |
| Other/Unknown | 16 (20.5%) | 88 (16.0%) | 167 (19.8%) | 4623 (14.7%) |

Data shown as frequency (percentage).
DM: Damaging missense.

Among the 293 variants from the aggregated list of clinical variants, 31 had alternate alleles present within the ADSP data: 4 in *PSEN2,* 25 in *PSEN1,* and 2 in *APP* (**Table 2**). All 31 variants seen within the ADSP participants are coding variants and were rare within the ADSP (MAF <1%). Overall, 78 ADSP participants (0.3%) carry at least one alternate allele of the variants curated from the clinical databases (**Table 2**). The 78 clinical variant carriers consisted of 75 DM variant carriers. *PSEN1* variant carriers constituted the greatest proportion (0.22%) of the clinical variant carriers, followed by *PSEN2* variant carriers (0.02%) and *APP* variant carriers (0.01%). One individual was identified as carrying a genetic variant in *APP* and *PSEN1*, however, the genetic variant in *APP* is a conflicting clinical variant with unclear consequences.

**Table 2.**
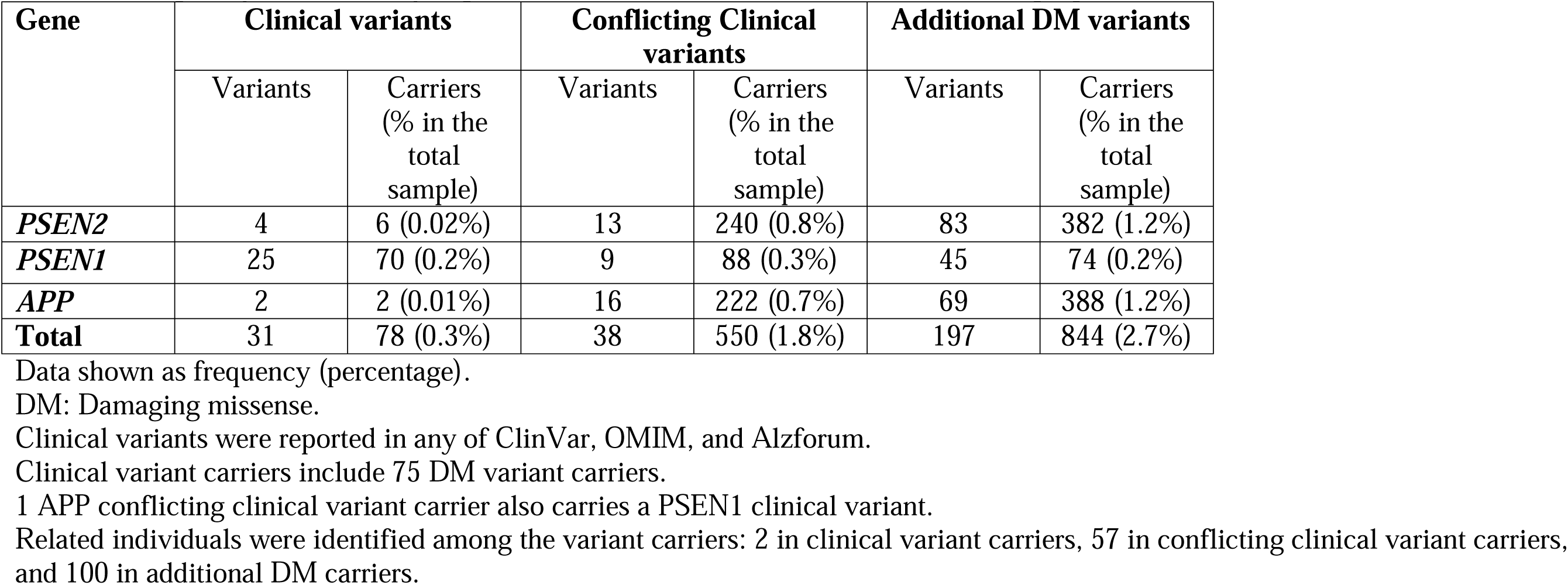
Frequency of Previously-reported Clinical Variants and Predicted Damaging Missense Variants within ADSP.

| Gene | Clinical variants |  | Conflicting Clinical variants |  | Additional DM variants |  |
| --- | --- | --- | --- | --- | --- | --- |
|  | Variants | Carriers<br>(% in the<br>total<br>sample) | Variants | Carriers<br>(% in the<br>total<br>sample) | Variants | Carriers<br>(% in the<br>total<br>sample) |
| *PSEN2* | 4 | 6 (0.02%) | 13 | 240 (0.8%) | 83 | 382 (1.2%) |
| *PSEN1* | 25 | 70 (0.2%) | 9 | 88 (0.3%) | 45 | 74 (0.2%) |
| *APP* | 2 | 2 (0.01%) | 16 | 222 (0.7%) | 69 | 388 (1.2%) |
| **Total** | 31 | 78 (0.3%) | 38 | 550 (1.8%) | 197 | 844 (2.7%) |

Data shown as frequency (percentage).
DM: Damaging missense.
Clinical variants were reported in any of ClinVar, OMIM, and Alzforum.
Clinical variant carriers include 75 DM variant carriers.
1 APP conflicting clinical variant carrier also carries a PSEN1 clinical variant.
Related individuals were identified among the variant carriers: 2 in clinical variant carriers, 57 in conflicting clinical variant carriers, and 100 in additional DM carriers.

**Table 3:**
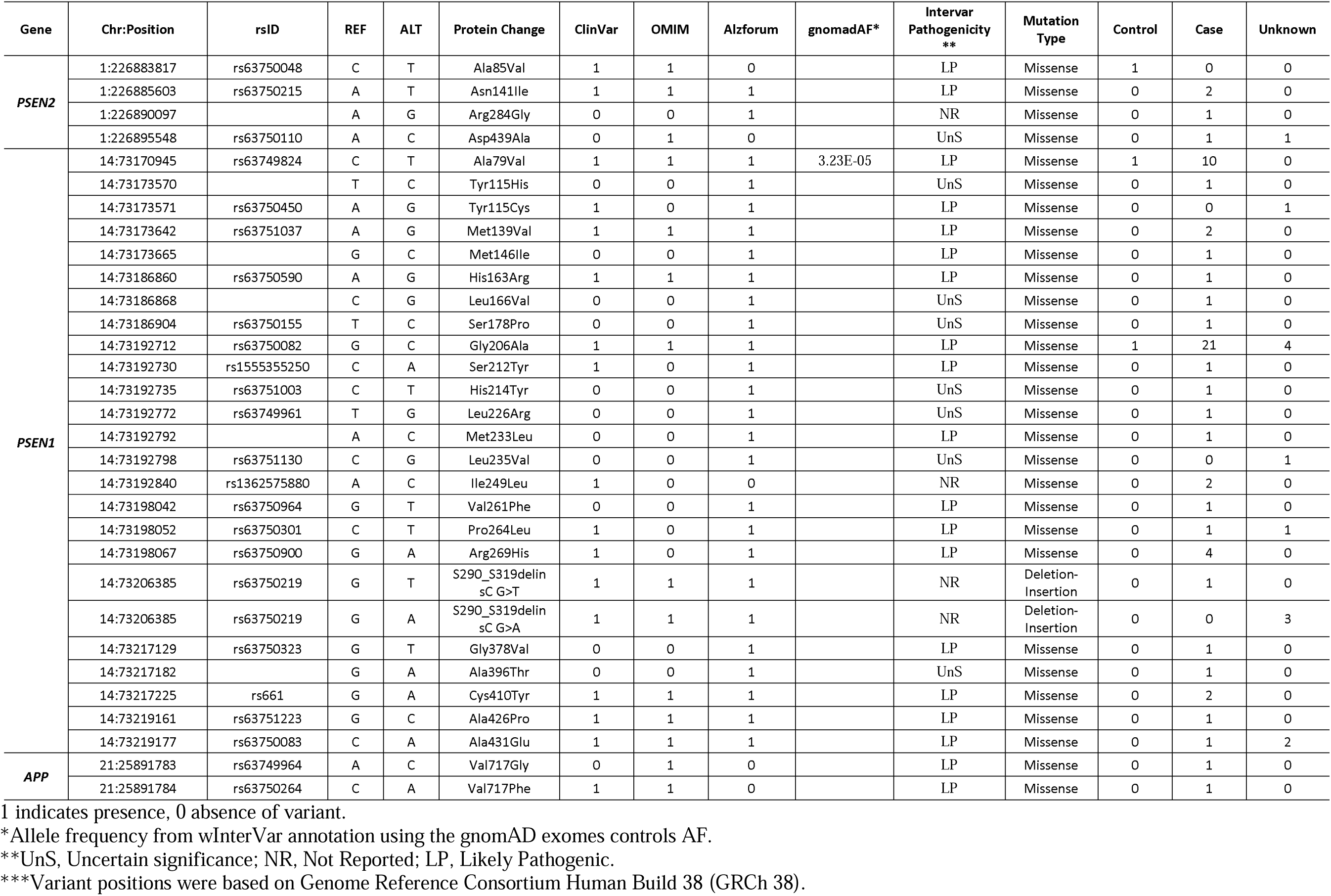
Clinical Variants with Alternative Alleles Observed within the ADSP.

| Gene | Chr:Position | rsID | REF | ALT | Protein Change | ClinVar | OMIM | Alzforum | gnomadAF* | InterVar Pathogenicity ** | Mutation Type | Control | Case | Unknown |
| --- | --- | --- | --- | --- | --- | --- | --- | --- | --- | --- | --- | --- | --- | --- |
| *PSEN2* | 1:226883817 | rs63750048 | C | T | Ala85Val | 1 | 1 | 0 |  | LP | Missense | 1 | 0 | 0 |
|  | 1:226885603 | rs63750215 | A | T | Asn141Ile | 1 | 1 | 1 |  | LP | Missense | 0 | 2 | 0 |
|  | 1:226890097 |  | A | G | Arg284Gly | 0 | 0 | 1 |  | NR | Missense | 0 | 1 | 0 |
|  | 1:226895548 | rs63750110 | A | C | Asp439Ala | 0 | 1 | 0 |  | UnS | Missense | 0 | 1 | 1 |
| *PSEN1* | 14:73170945 | rs63749824 | C | T | Ala79Val | 1 | 1 | 1 | 3.23E-05 | LP | Missense | 1 | 10 | 0 |
|  | 14:73173570 |  | T | C | Tyr115His | 0 | 0 | 1 |  | UnS | Missense | 0 | 1 | 0 |
|  | 14:73173571 | rs63750450 | A | G | Tyr115Cys | 1 | 0 | 1 |  | LP | Missense | 0 | 0 | 1 |
|  | 14:73173642 | rs63751037 | A | G | Met139Val | 1 | 1 | 1 |  | LP | Missense | 0 | 2 | 0 |
|  | 14:73173665 |  | G | C | Met146Ile | 0 | 0 | 1 |  | LP | Missense | 0 | 1 | 0 |
|  | 14:73186860 | rs63750590 | A | G | His163Arg | 1 | 1 | 1 |  | LP | Missense | 0 | 1 | 0 |
|  | 14:73186868 |  | C | G | Leu166Val | 0 | 0 | 1 |  | UnS | Missense | 0 | 1 | 0 |
|  | 14:73186904 | rs63750155 | T | C | Ser178Pro | 0 | 0 | 1 |  | UnS | Missense | 0 | 1 | 0 |
|  | 14:73192712 | rs63750082 | G | C | Gly206Ala | 1 | 1 | 1 |  | LP | Missense | 1 | 21 | 4 |
|  | 14:73192730 | rs1555355250 | C | A | Ser212Tyr | 1 | 0 | 1 |  | LP | Missense | 0 | 1 | 0 |
|  | 14:73192735 | rs63751003 | C | T | His214Tyr | 0 | 0 | 1 |  | UnS | Missense | 0 | 1 | 0 |
|  | 14:73192772 | rs63749961 | T | G | Leu226Arg | 0 | 0 | 1 |  | UnS | Missense | 0 | 1 | 0 |
|  | 14:73192792 |  | A | C | Met233Leu | 0 | 0 | 1 |  | LP | Missense | 0 | 1 | 0 |
|  | 14:73192798 | rs63751130 | C | G | Leu235Val | 0 | 0 | 1 |  | UnS | Missense | 0 | 0 | 1 |
|  | 14:73192840 | rs1362575880 | A | C | Ile249Leu | 1 | 0 | 0 |  | NR | Missense | 0 | 2 | 0 |
|  | 14:73198042 | rs63750964 | G | T | Val261Phe | 0 | 0 | 1 |  | LP | Missense | 0 | 1 | 0 |
|  | 14:73198052 | rs63750301 | C | T | Pro264Leu | 1 | 0 | 1 |  | LP | Missense | 0 | 1 | 1 |
|  | 14:73198067 | rs63750900 | G | A | Arg269His | 1 | 0 | 1 |  | LP | Missense | 0 | 4 | 0 |
|  | 14:73206385 | rs63750219 | G | T | S290_S319delin<br>sC G>T | 1 | 1 | 1 |  | NR | Deletion-Insertion | 0 | 1 | 0 |
|  | 14:73206385 | rs63750219 | G | A | S290_S319delin<br>sC G>A | 1 | 1 | 1 |  | NR | Deletion-Insertion | 0 | 0 | 3 |
|  | 14:73217129 | rs63750323 | G | T | Gly378Val | 0 | 0 | 1 |  | LP | Missense | 0 | 1 | 0 |
|  | 14:73217182 |  | G | A | Ala396Thr | 0 | 0 | 1 |  | UnS | Missense | 0 | 1 | 0 |
|  | 14:73217225 | rs661 | G | A | Cys410Tyr | 1 | 1 | 1 |  | LP | Missense | 0 | 2 | 0 |
|  | 14:73219161 | rs63751223 | G | C | Ala426Pro | 1 | 1 | 1 |  | LP | Missense | 0 | 1 | 0 |
|  | 14:73219177 | rs63750083 | C | A | Ala431Glu | 1 | 1 | 1 |  | LP | Missense | 0 | 1 | 2 |
| *APP* | 21:25891783 | rs63749964 | A | C | Val717Gly | 0 | 1 | 0 |  | LP | Missense | 0 | 1 | 0 |
|  | 21:25891784 | rs63750264 | C | A | Val717Phe | 1 | 1 | 0 |  | LP | Missense | 0 | 1 | 0 |

1 indicates presence, 0 absence of variant.
\*Allele frequency from wInterVar annotation using the gnomAD exomes controls AF.
\*\*UnS, Uncertain significance; NR, Not Reported; LP, Likely Pathogenic.
\*\*\*Variant positions were based on Genome Reference Consortium Human Build 38 (GRCh 38).

We have identified 3 families among the 78 clinical variant carriers in the family sub-study, which includes one pair of siblings both carrying a PSEN1 variant (rs63750082, p.Gly206Ala), one sporadic case without any other related individuals as clinical variant carriers, and 3 inferred cousins all carrying a PSEN1 deletion (rs63750219). Within the group of 78 individuals carrying at least one clinical variant, as expected, most were AD cases (N=62, 79%) or those with unknown AD status (N=13, 17%); very few were controls (N=3, 4%). Non-Hispanic White participants have the highest proportion of clinical variant carriers (59%) and are the largest proportion of the data (67%), whereas Non-Hispanic Blacks constitute 16.8% of the data, but are only 3.9% of the clinical variant carriers (**Table 1**).

The higher number of clinical variant carriers in cases compared to controls was consistent across the three genes. We observed the highest proportion of clinical variant carriers among cases for *PSEN1*, with 0.4% of the AD cases carrying a clinical variant as compared to 0.01% of the controls (**Figure 2 & Supplemental Table e-3**). Among *PSEN2* carriers, 0.03% of those who were diagnosed with AD carry an alternate allele in *PSEN2* variants compared to 0.01% of controls. And *APP* carrier rate is 0.01% in the AD cases as compared to 0% in the controls.

**Figure 2.**
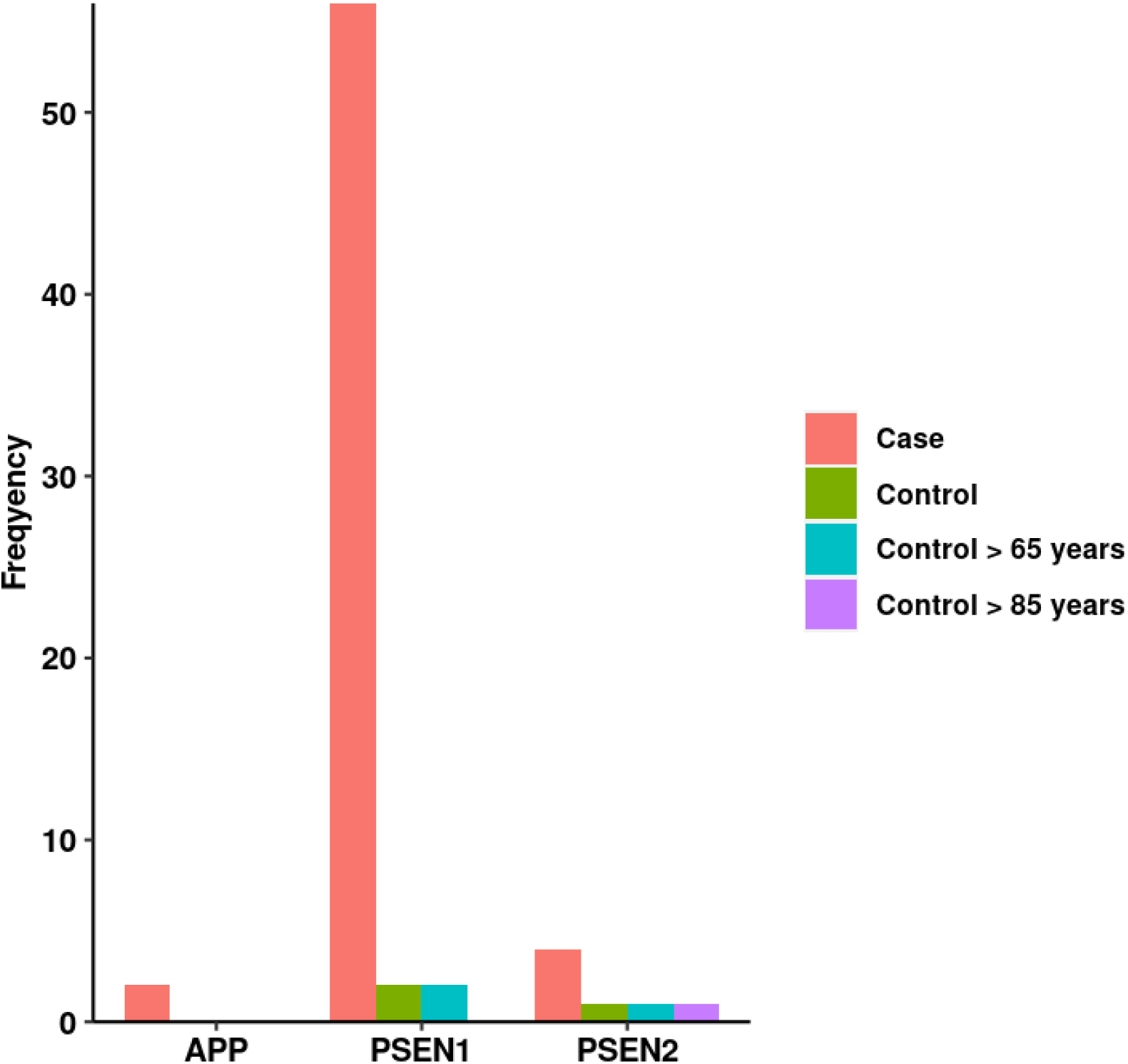
Distribution of AD Diagnosis by Gene among the Clinical variant Carriers. Bar chart represents the frequency of AD status for clinical variant carriers of APP, PSEN1, and PSEN2 genes. The Y axis is the count while the X-axis shows the specific genes. AD case and control status was shown by different colors on the legend.

Noteworthy, the higher proportion of *PSEN1* carriers among the cases are mainly due to two variants: rs63749824 (p.Ala79Val) and rs63750082 (p.Gly206Ala) (**Table 4**). When restricting controls to more advanced ages (>65 or > 85), we observed similar carrier rate differences between cases and elder controls for clinical variant carriers of all 3 genes (**Figure 2**).

We observed 38 out of 51 conflicting clinical variants, which consist of 13 *PSEN2* variants, 9 *PSEN1* variants, and 16 *APP* variants among the ADSP participants (**Table 2**). There were 550 individuals (1.75%) carrying at least one alternate allele of a conflicting clinical variant within the ADSP datasets, of which 240 are *PSEN2* variant carriers (0.76%), 88 are *PSEN1* variant carriers (0.28%), and 222 are *APP* variant carriers (0.70%) (**Table 2**). There were 57 related individuals among the conflicting clinical variant carriers. Among the conflicting clinical variant carriers, we observed a higher percentage of controls (N=270, 49.09%) than cases (N=224, 40.73%). Non-Hispanic White individuals (65.45%) constituted the largest proportion of the conflicting clinical variant carriers, followed by Non-Hispanic Black (17.27%), Non-Hispanic Other/Unknown (0%), Hispanic White (1.09%), Hispanic Black (0.18%), and Hispanic Other/Unknown individuals (16%) (**Table 1**).

We observed that the rates of conflicting clinical variant carriers are higher in controls for *PSEN2* and *APP* variants, with 0.80% and 0.76% of the controls carrying at least one conflicting clinical variant as compared to 0.69% and 0.64% of the cases, respectively. The rates for *PSEN1* conflicting clinical variant carriers were 0.27% of the controls were *PSEN1* variant carriers in contrast to 0.29% of the cases. And the rates of conflicting variant carriers increased when looking at elder controls except for *PSEN1* variant carriers (**Supplementary Table e-3**). We observed 23 variants that were seen more often in controls rather than cases: 11 in *PSEN2*, 5 in *PSEN1*, and 7 in *APP*, of which 5 variants were only seen in controls (rs201269325, p.Gly709Ser; rs63750831 p.Val94Met; rs115760359, p. Pro218Pro; rs63750227 p.Ala409Thr; rs866044092 p.Val150Met) (**Supplementary Table e-4**). Out of the 38 conflicting clinical variants, 27 of the conflicting clinical variants were all observed in both cases and controls, diminishing support for these variants (**Supplement Table e-4**).

Using VEP to annotate rare variants (MAF <1%) observed in the ADSP participants, we identified 197 rare variants (MAF < 1%) within ADSP which were not reported in known clinical databases. These variants were labeled as additional DM variants (**Supplement Table e-5**). Eighty-three out of the 197 additional DM variants were found in *PSEN2* with another 45 in *PSEN1* and 69 in *APP* (**Table 2**). The frequency of DM variant carriers is much higher than that of clinical or conflicting clinical variant carriers, with 844 participants carrying at least one alternate allele of the predicted DM variants. Specifically, 382 participants carry DM variants in *PSEN2*, 74 carry DM variants in *PSEN1*, and 388 carry DM variants in *APP.* And 100 individuals of all the additional DM variant carriers were first-degree relatives evaluated via the kinship coefficients. Among the additional DM variant carriers, 43.3% were AD cases 48.5% were controls, and the rest were with AD unknown status(**Table 1**). The frequency of additional DM variants in controls indicates that this class of variation is very diverse for identifying additional functional DM variants. Overall, the low clinical/DM variant carrier rates within ADSP participants suggest that it’s extremely unlikely for controls who are greater than 60 years old to be a carrier of a pathogenic Mendelian AD variant (**Supplementary Table e-3**). Furthermore, we observed that the carrier rate varied by NIAGADS defined study sites (**Supplementary Table e-6**), with highest carrier rates in a family study and a sample of individuals selected based on family history (the “enriched” sample within the ADSP Discovery case/control set). Because family history of AD is not available across the ADSP study, we were not able to evaluate the carrier rate by presence or absence of family history.

The ADSP samples included in this analysis are predominantly Non-Hispanic White (66.9%), followed by Non-Hispanic Black (16.85%), Hispanic White (1.02%), Hispanic Black (0.23%), Hispanic Other/Unknown (14.68%), and Non-Hispanic Other/Unknown (0.32%). None of the clinical variant carriers were of Non-Hispanic Other/Unknown population, and the lowest observed carrier rate was found among individuals reported as Hispanic Black (0.003%) compared to Non-Hispanic White (0.15%), Non-Hispanic Black (0.01%), Hispanic White age (0.04%), or Hispanic Other/Unknown (0.05%) (**Supplementary Table e-7**). Within each population, cases have a higher clinical variant carrier rate compared to the controls (**Supplementary Table e-7**).

We examined whether carrying an alternate allele of one of the curated variants is associated with the age of onset of AD. Among participants with AD and available age of onset data, we observed 62 clinical variant carriers, 224 conflicting clinical variant carriers, 365 additional DM carriers, and 13,169 noncarriers. Although *APOE* ε*2* and ε*4* are statistically associated with age of onset (p=5.59*10^-16^ and 6.13*10^-186^, separately), they do not change the association between variant carriers and age of onset in the linear mixed effects models. The mean age of onset of AD among clinical variant carriers was significantly lower (55.6 ± 10.9 range 34-76 years) than the mean age of onset in the noncarriers (75.6 ± 9.5 range 39-90 years) when accounting for relatedness among participants (score test p-value=7.8×10^-57^, **Figure 3**). Comparing between populations, the adjusted mean ages of onset are 57, 53.6, and 64.5 years for White, African American, and Unknown/other individuals, respectively (**Supplemental Table e-8)**. Individuals with Hispanic ethnicity have, on average, an AD onset 13 years later than those without Hispanic ethnicity (p=0.003). However, interpretation of these results must be placed in the context of the age-based risk score used for selection of a portion of the subjects included in the ADSP dataset.

**Figure 3.**
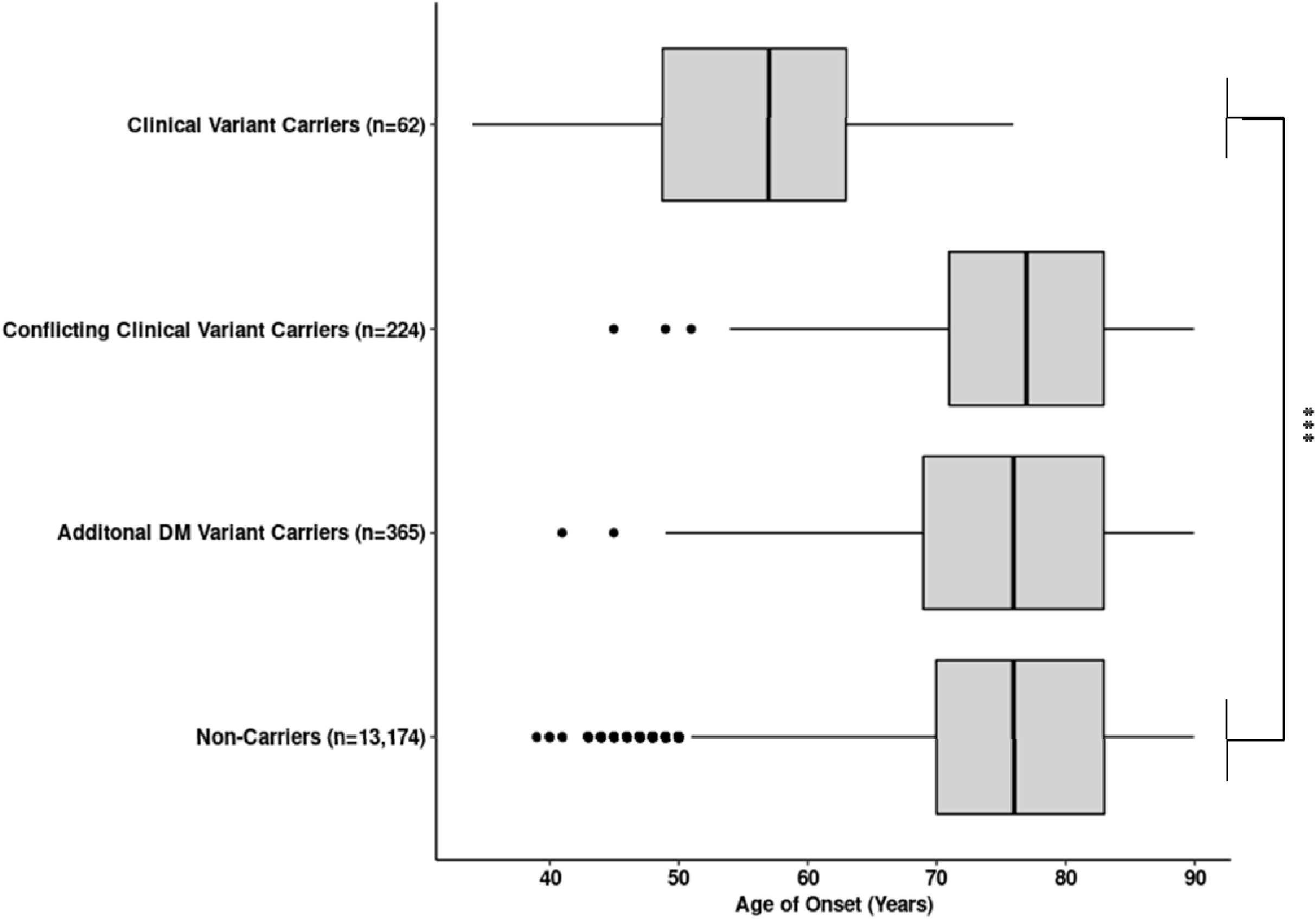
Age of AD Onset Among Carriers and Noncarriers. Boxplot showing the age of AD onset in years among the carriers and noncarriers of clinical, conflicting clinical, additional LoF, or additional DM variants. The mean and standard deviation (SD) of age of AD onset are 55.98 ± 10.88 years for clinical variant carriers, 76.30 ± 9.30 for conflicting clinical variant carriers, 75.28 ± 9.73 for additional DM variant carriers, and 75.56 ± 9.48 for non-carriers. Linear mixed models adjusted for sex, the first four principal components of ancestry, an indicator variable for WGS or WES data, APOE ε2, and APOE ε4 status, accounting for an empirical Balding-Nichols kinship matrix were used to contrast the mean age of AD onset between clinical variant carriers and noncarriers. The exact p-value for the comparison is 1.23x10^-64^. *** denotes p-value < 0.001.

When looking at the *APOE* allele distribution in clinical variant carriers and noncarriers, we found 1 control and 20 cases who carry at least one *APOE* ε4 allele among the clinical variant carriers. Six clinical variant carriers who were diagnosed with AD carry one *APOE* ε2 allele whereas none of the control clinical variant carriers were *APOE* ε2 carriers indicating they may harbor another protective variant. The *APOE* ε3/ε3 genotype is the most prevalent genotype both in cases and controls among the clinical variant carriers. There is a higher proportion of *APOE* ε2 allele carriers in the controls (15.8%) as compared to cases (8.1%) among the clinical variant noncarriers. Conversely, more cases (50.3%) were seen compared to controls (26.5%) among the *APOE* ε4 allele carriers (**Supplementary Table e-9**). *APOE* genotype was part of the case selection criterion for the WES case-control study, which can lead to an inverted association of *APOE* ε4 in some sets of individuals (i.e., cases selected without known genetic risk and controls who avoided AD despite known genetic risk). We didn’t observe any statistical evidence for potential protective or synergic effect by the *APOE* ε alleles with the clinical variant carriers of the 3 Mendelian genes, however, this might be due to limited clinical carrier counts hence we lack the power to formally test the gene by gene interaction on AD.

## DISCUSSION

Two studies using a prior release of ADSP data examined variants within *APP*, *PSEN1*, *PSEN2*, and other dementia-related genes[7,8]. Blue et al. examined 578 WGS and 10,836 WES samples and Fernandez et al. examined 143 WGS and 10,280 WES samples from the ADSP. The current study, using a more recent data release, doubles the sample size over the prior studies and increases the diversity among the study participants. We identified variants in the *PSEN2, PSEN1,* and *APP* genes previously implicated in Mendelian AD within the ADSP WES and WGS datasets. Among clinical variant carriers overall, a higher proportion were cases compared to controls. However, some previously-reported clinical variants were observed in control individuals. This may reflect the age of the carrier. Clinical variant carriers who were controls had ages of 73, 74, and 90 and some of these individuals may manifest AD in the future. Our observation of clinical variants in controls may also reflect reduced penetrance or the misclassification of a variant as clinically relevant for AD. Within the ADSP, AD cases were more likely to be carrying clinical variants in *PSEN2*, *PSEN1* or *APP* (0.03%, 0.41% and 0.01%, respectively) compared to controls (**Supplementary Table e-3**). However, we observed higher proportions of controls carrying conflicting clinical variants in *PSEN2* and *APP* (**Supplementary Table e-3**). This could be explained by varying penetrance of the variants or that these variants are potentially not causal for AD.

Four conflicting clinical variants were labeled as pathogenic/likely pathogenic in either OMIM or Alzforum but not in ClinVar (rs63750197 *PSEN2* p.Ser130Leu, rs63750831 *PSEN1* p.Val94Met, rs63750847 *APP* p.Ala673Thr, rs63750363 *APP* p. Glu665Asp) (**Supplementary Table e-**4). Arboleda-Velasquez et al. reported the protective effect of the APOE3ch mutation (rs121918393.p.Arg154Ser) against a *PSEN1* variant (rs63750231.p. Glu280Ala)[18]. However, both the *APOE3ch* and *PSEN1* variants were not present in our ADSP sample. In a recent report, Xian et al. re-evaluated 452 pathogenic variants in *PSEN1*, *PSEN2*, and *APP* genes from PubMed and Alzforum according to the ACMG-AMP guidelines and found 5.09% of the variants were of uncertain significance[19]. Altogether, these studies support the hypothesis that there is misclassification or reduced penetrance of the pathogenic/likely pathogenic variants from ClinVar, OMIM, and Alzforum, and that our data can aide in reclassifying these variants.

We observed that the mean age of onset for clinical variant carriers was significantly earlier than that for noncarriers. The adjusted mean ages of onset for clinical variant carriers with reported African American and Unknown/Other race is 53.6 years and 64.5 years, respectively (**Supplemental Table e<u>-8</u>**). Clinical variant carriers with reported Hispanic ethnicity on average have AD onset at 67.1 years as compared to 54.1 years for those with non-Hispanic ethnicity, and the difference is statistically significant (p=0.003). However, this result should be interpreted with caution given distinct ascertainment schemes in different phases of the ADSP, some of which included exclusion thresholds for age at onset. Onset in the 4^th^ and 5^th^ decade of life was seen in individuals who did not carry an alternate allele at an identified clinical variant, suggesting that additional variants, potentially in other genes, driving early onset AD have yet to be identified.

Although the ADSP participants used in this analysis are predominantly Non-Hispanic White, a substantial proportion (33.1%) of the subjects are reported as other races/ethnicities, primarily Non-Hispanic Black and Hispanic. The carrier rates for the clinical variants were not similar among defined populations. The highest carrier rate was present among Non-Hispanic White individuals and the lowest carrier rate among participants from the Non-Hispanic Other/Unknown population (0.15% and 0%, respectively) (**Supplementary Table e-8**). Variant level disparities among race/ethnicity groups were not reported due to the fact that the clinical variants are very rare in the ADSP with only 1 or 2 variants present in each non-White population. When looking at the clinical variant count by AD status among race/ethnicity groups, we identified one variant that is disproportionally presented in AD cases with Hispanic ethnicity (rs63750082 PSEN1 p. Gly206Ala, Supplemental Table e-10). Additionally, rs63750083 PSEN1 p. Ala431Glu and rs63751130 PSEN1 p. Leu235Val were only observed in one AD case and three AD unknown individuals with Hispanic ethnicity. These variants may have unique pathogenicity mechanism which requires careful interpretations and further investigations.

Previous studies have shown that disparities in inclusion in genomic sequencing may result in misclassification or a higher rate of variants of uncertain significance for patients in underrepresented populations[20]. A study conducted on assessing the underlying genetic risk for hypertrophic cardiomyopathy showed that variants that had been classified as pathogenic were actually common variants in the African American population[21]. These variants were misclassified as pathogenic due to the lack of African American controls in previous studies, emphasizing the importance of including diverse ancestry groups in order to avoid health disparities as a result of misdiagnosis[21]. Another study showed that the sample sizes of most non-White populations within The Cancer Genome Atlas were not sufficient to detect common variants across several cancer types specific to different racial and ethnic groups[22]. These studies demonstrate the importance of including diverse populations in genetic studies in order to aid in accurate disease diagnosis and treatment. Further investigation is needed to confirm the differences in clinical variant carrier rates among populations within the ADSP. These differences could be due to a true difference in the frequency of AD causal PSEN2, PSEN1, APP variants across populations, or may reflect study design including ascertainment criteria, higher propensity for clinical variant screening for these three genes in White participants, or limitations due to populations historically included in genetic studies of AD. And the emphasis of the ADSP on increased diversity of study participants will help to answer some of these questions.

There were 2,950 individuals with unknown AD status in our study sample, out of which we identified 13 clinical variant carriers, 56 conflicting clinical variant carriers, and 70 additional DM variant carriers (**Supplementary Table e-11**). 16.7% of the clinical variant carriers had unknown AD status compared to 3.85% being as controls, indicating a mixture of potential AD cases and controls among the AD unknown individuals (**Table 1**). Upon further examination of the phenotypic data, 775 individuals out of the 2,950 do not have any phenotypic data, 313 individuals from the ADNI study had mild cognitive impairment (MCI), 549 individuals were diagnosed with progressive supranuclear palsy (PSP) while 332 individuals had corticobasal degeneration (CBD), an additional 202 individuals were from family studies, with another 779 individuals enrolled from ADSP case-control studies (**Supplementary Table e-12**).

In conclusion, we identified variants in the *PSEN2, PSEN1, APP* genes within the ADSP WES and WGS datasets. These data suggest conflicting clinical variants that may not be causal for AD. New DM variants which are potentially associated with AD were also reported via dbNFSP prediction. Additional studies are needed to determine whether these are functional variants. The ADSP generates WES and WGS datasets that are jointly called and QC’ed across studies. These genomic files, along with phenotype files that include AD status, are available to qualified researchers to facilitate the discovery of variants that are protective for or confer risk for AD. There are over 145 approved ADSP data access requests from academic and industry investigators across the US and internationally. Understanding the presence of known Mendelian AD variants within the ADSP data will have broad impact by informing analyses of variants at other loci in relation to AD risk.

## Data Availability

All data produced in the present study are available upon formal request to the Alzheimer's Disease Sequencing Project (ADSP) and The National Institute on Aging Genetics of Alzheimer's Disease Data Storage Site (NIGADS)

https://www.niagads.org/

## Acknowledgement

We thank Alzforum for providing the details of the variants in mendelian AD genes from the Alzforum database. ADSP data for this study were prepared, archived, and distributed by the National Institute on Aging Alzheimer’s Disease Data Storage Site (NIAGADS) at the University of Pennsylvania (U24-AG041689), funded by the National Institute on Aging (accession NG00067). The full acknowledgement statement for the ADSP can be found at: https://dss.niagads.org/datasets/ng00067/. ADNI data used in preparation of this article were obtained through NIAGADS. The investigators within the ADNI contributed to the design and implementation of ADNI and/or provided data but did not participate in analysis or writing of this report. A complete listing of ADNI investigators can be found at: http://adni.loni.usc.edu/wp-content/uploads/how_to_apply/ADNI_Acknowledgement_List.pdf

## Author Contributions

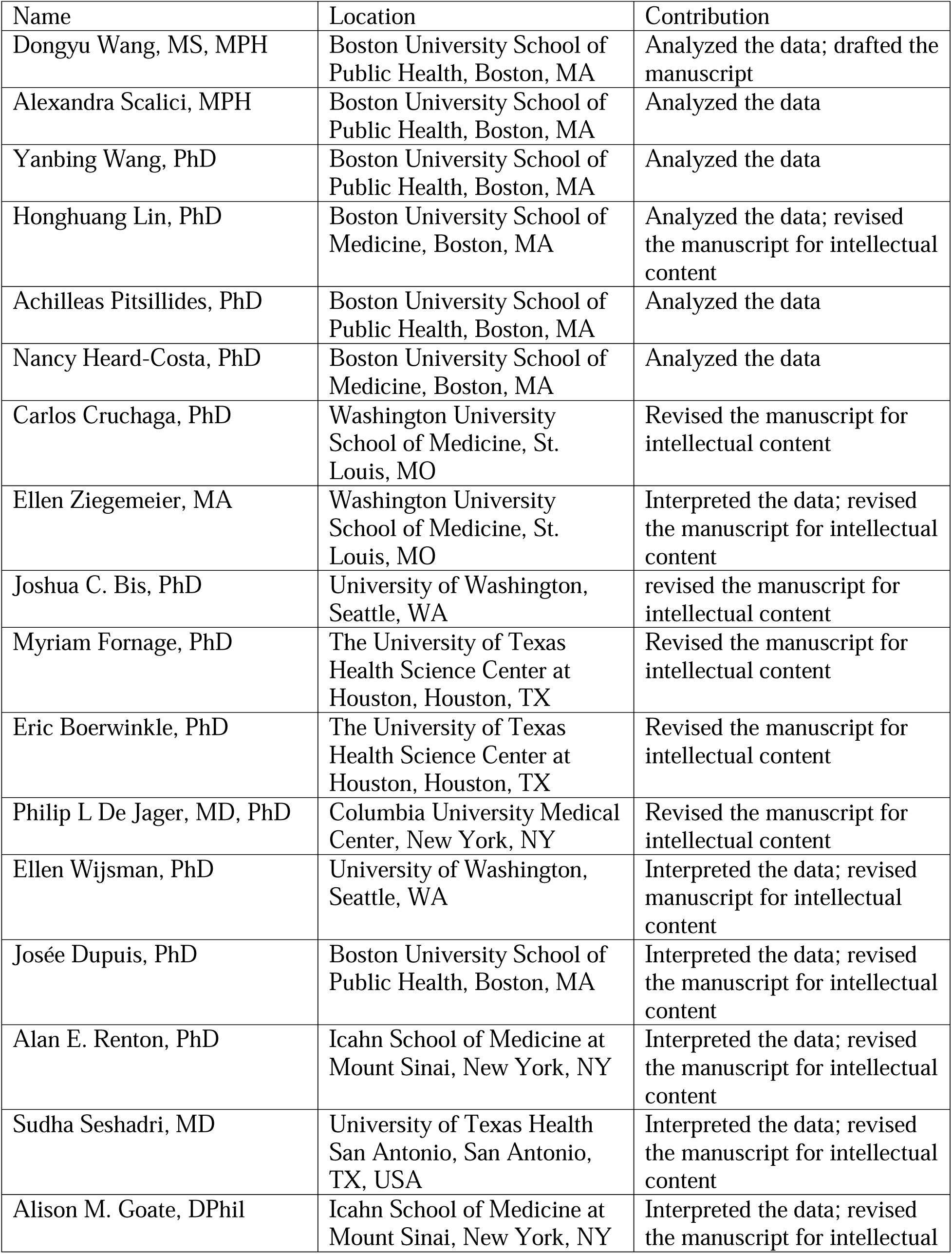
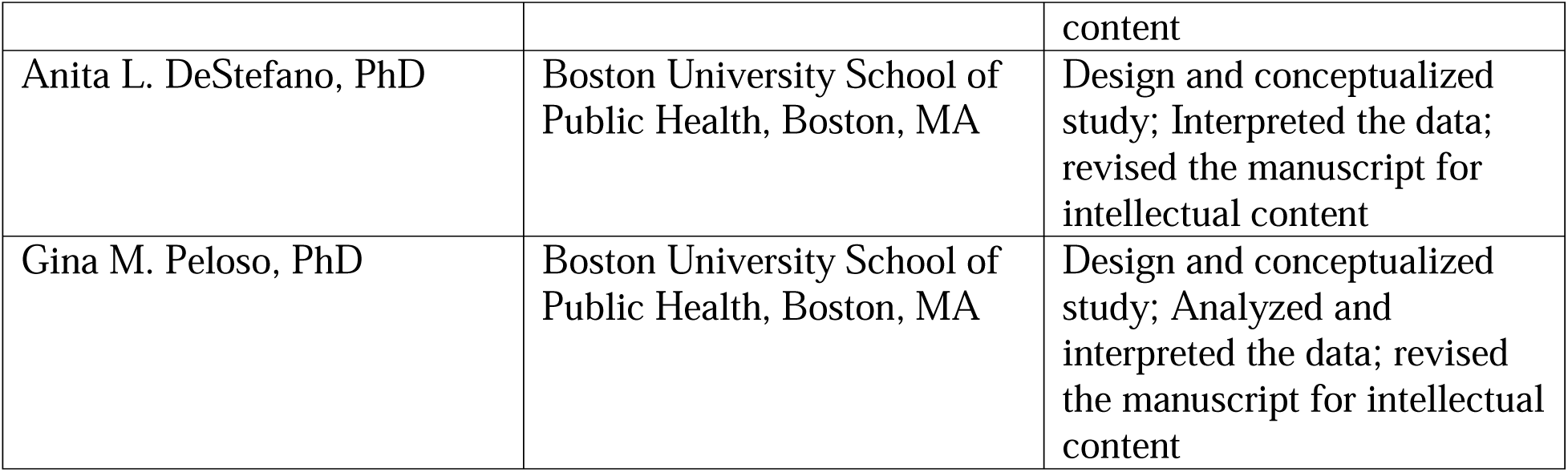

