## Supplemental Material for "Frequency of Variants in Mendelian Alzheimer’s Disease Genes within the Alzheimer’s Disease Sequencing Project (ADSP)"

**Supplemental Methods**

**ADSP data**

The ADSP data included in this study are comprised of distinct phases including the Discovery, Discovery Extension, Augmentation phases, and the Follow Up Study (FUS) phase. In the Discovery phase, as previously described,^1,2^ the majority of study participants for WES were selected based on computing a risk score that included dosage of the *APOE* ε2/ε3/ε4 alleles, sex and either onset age (for cases) or age at last exam for controls (or pathology-based adjusted age at death for neuropathology controls). The goal was to select cases with age at onset greater than 60 whose AD risk did not appear to be explained by age, sex, or *APOE* ε allele count and to select controls who were older than 60 years of age at last exam and whose risk score indicated they were least likely to develop AD by age 85. Additional “enriched” cases were selected from families with at least three family members with AD and no known *APP, PSEN1/2* mutations. The Discovery phase WGS was generated in individuals from multiplex AD families as previously described.^1^ The Discovery Extension phase, which consisted of WGS, included both a family and a case-control component. The Discovery Extension family WGS was generated on additional members of selected families from the Discovery phase as well as members of 77 additional families. A set of 114 Hispanic control individuals were also sequenced in this phase. A focus of the Discovery Extension case control component was to increase diversity among the ADSP samples. The ADSP Discovery Extension WGS was generated on 3,082 samples, with approximately one third from individuals of European, African, and Hispanic ancestry. The Augmentation phase comprises sequence data generated outside of the ADSP to boost the sample size for identifying rare mutations related to AD. Participants in the Discovery Extension and Augmentation phases were not selected using the risk score implemented in the Discovery phase. The Follow Up Study collects and sequences existing ethnically diverse and unique cohorts with clinical data to expand the utility of new discoveries for individuals from all populations. The first release of FUS data 3,250 AD cases, 4,149 cognitively normal individuals, 194 individuals with mild cognitive impairment, and 567 with unknown or other dementia from seven datasets (PR1066, ADC Autopsy, ADGC African American (release 1), HIHG Brain Bank, ADNI-WGS-2, APOE Extremes, and StEP AD). The current study includes the ADSP Discovery, Discovery Extension phases, Follow Up Study phase with additional participants via the Augmentation Phase. A detailed list of the cohorts included in our analysis can be found in the supplement (**Supplementary Table e-6**).

The ADSP data coordinating center, GCAD, produced a jointly called and QC’ed WGS dataset that included the ADSP WGS Discovery, Discovery Extension, Follow UP Study, and from the Alzheimer’s Disease Neuroimaging Initiative (ADNI) in the Augmentation phase. For WES, the GCAD jointly called and QC’ed dataset includes the ADSP Discovery phase WES and Augmentation phase WES from the following studies: Alzheimer’s Disease Genetics Consortium (ADGC) African American study, Familial Alzheimer Sequencing (FASe) project, Brkanac Families, Miami Families, Columbia Washington Heights/Inwood Columbia Aging Project (WHICAP), Charles F. and Joanne Knight Alzheimer’s Disease Research Center (Knight ADRC), Corticobasal degeneration (CBD) and Progressive supranuclear palsy (PSP) studies. AD phenotype data were not collected for CBD or PSP participants; hence these individuals were excluded from the current analysis. Details of studies included in the ADSP can be found at The National Institute on Aging Genetics of Alzheimer’s Disease Data Storage Site (NIAGADS; <https://dss.niagads.org/>) under dataset: NG00067 ADSP Umbrella Study (<https://dss.niagads.org/datasets/ng00067/>).

In the ADSP Discovery phase, families carrying known Mendelian mutations in *APP*, *PSEN1*, or *PSEN2* based on screening at least one affected individual per family were excluded from sequencing. Screening may have occurred in participants in other contributing studies. However, data on who was screened and details of screening (e.g. technology, which mutations) are not in the ADSP datasets. In the ADSP Discovery and Discovery Extension phases sequencing was performed at three sequencing centers via the National Human Genome Research Institute (NHGRI). Sequence data for ADSP Augmentation Studies are generated under NIA and private funding and are shared with the research community through NIAGADS.

**Quality Control filtering**

The Genomic Center for Alzheimer Disease (GCAD) performs single nucleotide and insertion deletion variant calling for the ADSP. They deliver project level vcf files that include all variants called with flags indicating quality control (QC) metrics that can be used for filtering. For the current study the GCAD provided QC flags were used to identify and retain high quality variants within the ADSP pvcfs. Specifically, for WGS data variants that did not receive a GATK pass, that were monomorphic across all samples, or had low call rate across all studies were excluded. Additionally, genotypes were set to missing for all individuals within a study if the variant had high mean depth or an ABhet ratio outside of 0.25 to 0.75. For WES data, the same filtering was implemented with the following modifications: genotypes were set to missing within a study if a low call rate was observed within that study and if variants were outside of the target region defined as the intersection of the target capture kits used by all contributing studies. Details of the GCAD provided QC flags can be found in the readme files at NIAGADS.

**Supplementary Table e-1: Mutation Selection Criteria in ClinVar, OMIM, and Alzforum.**

| Criteria | Search Terms |
| --- | --- |
| Pathogenicity | "Pathogenic"  "Likely pathogenic"  "Pathogenic/Likely pathogenic"  "Conflicting interpretations of pathogenicity" |
| Alzheimer’s, Dementia  or Other Related Disorder | "Alzheimer disease, early-onset, with cerebral amyloid angiopathy"  "Alzheimer disease familial 3, with spastic paraparesis"  "Alzheimer disease, familial, 3, with spastic paraparesis and apraxia"  "Alzheimer disease, familial, 3, with spastic paraparesis and unusual plaques"  "Alzheimer disease, familial, 3, with unusual plaques"  "Alzheimer disease, familial, with spastic paraparesis and unusual plaques"  "Alzheimer disease, protection against"  "Alzheimer disease, type 1"  "Alzheimer disease, type 3"  "Alzheimer disease, type 4"  “Alzheimer disease familial type 4”  "Alzheimer's disease"  "Cerebral amyloid angiopathy, APP-related"  "CEREBRAL AMYLOID ANGIOPATHY, APP-RELATED, PIEDMONT VARIANT"  "Dementia"  "Early-Onset Familial Alzheimer Disease"  "Frontotemporal dementia"  "Huntington disease-like syndrome"  "Inborn genetic diseases"  "Mental deterioration"  "Pick's diseases" |

**Supplementary Table e-2: List of Mutations Meeting the Criteria Listed in Table e-1 (Clinical Variants or Conflicting Clinical Variants) from ClinVar, OMIM, and Alzforum.**

| **CHROM:POS:REF:ALT** | **ClinVar** | **OMIM** | **Alzforum** |
| --- | --- | --- | --- |
| 1:226881888:: | 1 | 0 | 0 |
| 1:226882007:G:A | 1 | 0 | 0 |
| 1:226883729:G:A | 1 | 0 | 0 |
| 1:226883771:G:A | 1 | 0 | 0 |
| 1:226883817:C:T | 1 | 1 | 0 |
| 1:226883863:C:T | 1 | 0 | 0 |
| 1:226883899:C:T | 1 | 0 | 0 |
| 1:226885545:A:C | 1 | 1 | 1 |
| 1:226885546:C:G | 1 | 1 | 0 |
| 1:226885570:C:T | 0 | 1 | 0 |
| 1:226885596:G:A | 1 | 0 | 0 |
| 1:226885602:A:T | 0 | 0 | 1 |
| 1:226885603:A:T | 1 | 1 | 1 |
| 1:226885629:G:A | 1 | 0 | 0 |
| 1:226888952:C:G | 1 | 0 | 0 |
| 1:226888973:G:A | 1 | 0 | 0 |
| 1:226888974:C:T | 1 | 0 | 0 |
| 1:226888977:A:G | 1 | 1 | 1 |
| 1:226888979:G:A | 1 | 1 | 1 |
| 1:226889016:G:A | 1 | 0 | 0 |
| 1:226890097:A:G | 0 | 0 | 1 |
| 1:226890135:TGA: | 1 | 0 | 0 |
| 1:226891284:T:C | 0 | 0 | 1 |
| 1:226891345:C:T | 1 | 0 | 0 |
| 1:226894073:C:A | 1 | 0 | 0 |
| 1:226895521:C:T | 0 | 1 | 0 |
| 1:226895548:A:C | 0 | 1 | 0 |
| 1:226895650:C:A | 0 | 0 | 1 |
| 1:226895699:G:A | 1 | 0 | 0 |
| 14:73170813:G:A | 1 | 0 | 0 |
| 14:73170854:C:G | 1 | 0 | 0 |
| 14:73170943:C:T | 1 | 0 | 0 |
| 14:73170945:C:T | 1 | 1 | 1 |
| 14:73170953:G:C | 0 | 0 | 1 |
| 14:73170957:T:C | 0 | 0 | 1 |
| 14:73170959:A:G | 0 | 0 | 1 |
| 14:73170961:G:C | 1 | 0 | 0 |
| 14:73170963:T:C | 1 | 1 | 1 |
| 14:73170972:C:G | 1 | 0 | 0 |
| 14:73170972:C:T | 0 | 0 | 1 |
| 14:73170974:G:C | 0 | 0 | 1 |
| 14:73170974:G:T | 0 | 0 | 1 |
| 14:73170983:T:A | 1 | 0 | 0 |
| 14:73170984:G:C | 0 | 1 | 1 |
| 14:73170989:G:A | 0 | 0 | 1 |
| 14:73170995:G:T | 0 | 0 | 1 |
| 14:73170998:G:T | 0 | 0 | 1 |
| 14:73171007:A:T | 0 | 0 | 1 |
| 14:73171017:T:G | 1 | 0 | 0 |
| 14:73171017:TCA: | 0 | 0 | 1 |
| 14:73171022:T:A | 0 | 0 | 1 |
| 14:73171022:T:G | 0 | 0 | 1 |
| 14:73171023:T:G | 1 | 0 | 1 |
| 14:73171024:T:G | 0 | 0 | 1 |
| 14:73171040:G:T | 0 | 0 | 1 |
| 14:73171041:G:T | 0 | 0 | 1 |
| 14:73171047:T:C | 1 | 1 | 1 |
| 14:73171048:: | 0 | 1 | 0 |
| 14:73171048:G: | 0 | 0 | 1 |
| 14:73171054:A:G | 1 | 0 | 0 |
| 14:73173570:T:C | 0 | 0 | 1 |
| 14:73173571:A:G | 1 | 0 | 1 |
| 14:73173574:C:A | 1 | 0 | 1 |
| 14:73173574:C:T | 1 | 0 | 1 |
| 14:73173576:C:A | 0 | 0 | 1 |
| 14:73173576:C:G | 0 | 0 | 1 |
| 14:73173576:C:T | 1 | 0 | 1 |
| 14:73173577:C:A | 0 | 0 | 1 |
| 14:73173577:C:G | 0 | 0 | 1 |
| 14:73173577:C:T | 1 | 0 | 1 |
| 14:73173583:C:T | 1 | 0 | 1 |
| 14:73173585:G:A | 0 | 0 | 1 |
| 14:73173586:A:G | 0 | 0 | 1 |
| 14:73173587:A:C | 0 | 0 | 1 |
| 14:73173587:A:T | 0 | 1 | 1 |
| 14:73173591:A:G | 1 | 0 | 0 |
| 14:73173619:A:G | 0 | 0 | 1 |
| 14:73173630:A:G | 1 | 0 | 1 |
| 14:73173631:A:G | 1 | 0 | 1 |
| 14:73173636:G:A | 1 | 0 | 0 |
| 14:73173642:A:G | 1 | 1 | 1 |
| 14:73173642:A:T | 0 | 0 | 1 |
| 14:73173643:T:A | 0 | 0 | 1 |
| 14:73173643:T:C | 0 | 0 | 1 |
| 14:73173644:G:A | 0 | 0 | 1 |
| 14:73173644:G:C | 0 | 0 | 1 |
| 14:73173651:G:A | 1 | 0 | 1 |
| 14:73173651:G:T | 0 | 0 | 1 |
| 14:73173654:A:G | 1 | 0 | 0 |
| 14:73173654:A:T | 0 | 0 | 1 |
| 14:73173655:T:A | 0 | 0 | 1 |
| 14:73173655:T:C | 1 | 0 | 1 |
| 14:73173656:T:G | 0 | 0 | 1 |
| 14:73173663:A:C | 1 | 1 | 1 |
| 14:73173663:A:C | 1 | 1 | 1 |
| 14:73173663:A:G | 1 | 1 | 1 |
| 14:73173663:A:G | 1 | 1 | 1 |
| 14:73173663:A:T | 1 | 1 | 1 |
| 14:73173663:A:T | 1 | 1 | 1 |
| 14:73173665:G:A | 0 | 0 | 1 |
| 14:73173665:G:C | 0 | 0 | 1 |
| 14:73173665:G:T | 1 | 1 | 0 |
| 14:73173667:C:T | 0 | 0 | 1 |
| 14:73173684:C:G | 0 | 0 | 1 |
| 14:73173687:T:A | 0 | 0 | 1 |
| 14:73173691:AT:ATTTATAT | 1 | 1 | 0 |
| 14:73173693::TTATAT | 0 | 0 | 1 |
| 14:73173702:T:C | 1 | 0 | 0 |
| 14:73173703:A:C | 0 | 0 | 1 |
| 14:73173703:A:T | 1 | 0 | 1 |
| 14:73186859:C:T | 1 | 1 | 1 |
| 14:73186860:A:G | 1 | 1 | 1 |
| 14:73186867:G:C | 0 | 0 | 1 |
| 14:73186867:G:T | 0 | 0 | 1 |
| 14:73186868:C:G | 0 | 0 | 1 |
| 14:73186869:T:C | 1 | 1 | 1 |
| 14:73186869:T:G | 0 | 0 | 1 |
| 14:73186872:TTA: | 0 | 0 | 1 |
| 14:73186876:ATCATC:ATC | 1 | 0 | 0 |
| 14:73186877:T:C | 0 | 0 | 1 |
| 14:73186878:C:T | 1 | 0 | 1 |
| 14:73186879:ATC: | 0 | 0 | 1 |
| 14:73186881:C:T | 0 | 1 | 1 |
| 14:73186881:T:TTAT | 1 | 0 | 0 |
| 14:73186884:T:C | 1 | 0 | 1 |
| 14:73186890:T:C | 0 | 0 | 1 |
| 14:73186890:T:G | 0 | 0 | 1 |
| 14:73186891:G:C | 0 | 0 | 1 |
| 14:73186891:G:T | 0 | 0 | 1 |
| 14:73186892:C:A | 0 | 1 | 1 |
| 14:73186893:T:G | 0 | 0 | 1 |
| 14:73186901:T:C | 0 | 0 | 1 |
| 14:73186901:T:G | 0 | 0 | 1 |
| 14:73186902:T:C | 1 | 0 | 1 |
| 14:73186904:T:C | 0 | 0 | 1 |
| 14:73186920:G:T | 1 | 1 | 0 |
| 14:73192646:A:G | 0 | 0 | 1 |
| 14:73192647:A:C | 1 | 0 | 1 |
| 14:73192699:A:T | 0 | 0 | 1 |
| 14:73192709:TTG: | 0 | 0 | 1 |
| 14:73192711:G:A | 0 | 0 | 1 |
| 14:73192712:G:A | 0 | 0 | 1 |
| 14:73192712:G:C | 1 | 1 | 1 |
| 14:73192712:G:T | 0 | 0 | 1 |
| 14:73192720:G:A | 0 | 0 | 1 |
| 14:73192721:G:A | 1 | 0 | 1 |
| 14:73192721:G:C | 0 | 0 | 1 |
| 14:73192721:G:T | 1 | 0 | 1 |
| 14:73192730:C:A | 1 | 0 | 1 |
| 14:73192732:A:C | 0 | 0 | 1 |
| 14:73192732:A:T | 0 | 0 | 1 |
| 14:73192733:T:C | 0 | 0 | 1 |
| 14:73192735:C:A | 1 | 0 | 0 |
| 14:73192735:C:T | 0 | 0 | 1 |
| 14:73192736:A:G | 0 | 0 | 1 |
| 14:73192744:G:C | 1 | 1 | 1 |
| 14:73192745:G:A | 0 | 0 | 1 |
| 14:73192749:A:G | 1 | 0 | 0 |
| 14:73192760:A:C | 1 | 0 | 0 |
| 14:73192760:A:G | 0 | 0 | 1 |
| 14:73192761:G:C | 0 | 0 | 1 |
| 14:73192763:A:G | 0 | 0 | 1 |
| 14:73192771:C:T | 0 | 0 | 1 |
| 14:73192772:T:G | 0 | 0 | 1 |
| 14:73192784:G:A | 0 | 0 | 1 |
| 14:73192786:G:A | 1 | 0 | 1 |
| 14:73192792:A:C | 0 | 0 | 1 |
| 14:73192792:A:G | 1 | 0 | 1 |
| 14:73192793:T:C | 1 | 0 | 1 |
| 14:73192794:G:A | 0 | 0 | 1 |
| 14:73192798:C:G | 0 | 0 | 1 |
| 14:73192799:T:C | 0 | 0 | 1 |
| 14:73192804:T:C | 1 | 0 | 1 |
| 14:73192809:C:G | 0 | 0 | 1 |
| 14:73192818:CCC:CC | 1 | 0 | 0 |
| 14:73192831:G:C | 0 | 0 | 1 |
| 14:73192832:C:A | 1 | 1 | 1 |
| 14:73192838:T:C | 0 | 0 | 1 |
| 14:73192840:A:C | 1 | 0 | 0 |
| 14:73192840:A:T | 1 | 0 | 0 |
| 14:73192843:T:G | 0 | 0 | 1 |
| 14:73192844:T:C | 0 | 1 | 1 |
| 14:73192845:G:T | 1 | 0 | 1 |
| 14:73192861:T:A | 1 | 0 | 1 |
| 14:73198040:C:T | 0 | 0 | 1 |
| 14:73198042:G:T | 0 | 0 | 1 |
| 14:73198043:T:C | 1 | 0 | 0 |
| 14:73198045:T:G | 0 | 0 | 1 |
| 14:73198047:G:C | 0 | 0 | 1 |
| 14:73198048:T:C | 0 | 0 | 1 |
| 14:73198049:G:T | 0 | 0 | 1 |
| 14:73198050:T:G | 0 | 0 | 1 |
| 14:73198052:C:T | 1 | 0 | 1 |
| 14:73198053:G:T | 1 | 0 | 0 |
| 14:73198057:G:A | 1 | 1 | 1 |
| 14:73198060:C:G | 0 | 0 | 1 |
| 14:73198060:C:T | 1 | 1 | 1 |
| 14:73198061:C:T | 1 | 0 | 0 |
| 14:73198066:C:G | 0 | 0 | 1 |
| 14:73198067:G:A | 1 | 0 | 1 |
| 14:73198072:C:G | 1 | 1 | 1 |
| 14:73198076:T:C | 0 | 0 | 1 |
| 14:73198079:A:C | 0 | 0 | 1 |
| 14:73198079:A:G | 0 | 0 | 1 |
| 14:73198085:C:T | 0 | 0 | 1 |
| 14:73198094:G:C | 1 | 1 | 0 |
| 14:73198094:G:C | 1 | 1 | 0 |
| 14:73198094:G:T | 1 | 1 | 1 |
| 14:73198094:G:T | 1 | 1 | 1 |
| 14:73198099:G:A | 1 | 0 | 1 |
| 14:73198100:A:C | 1 | 1 | 1 |
| 14:73198100:A:C | 1 | 1 | 1 |
| 14:73198100:A:G | 1 | 1 | 1 |
| 14:73198100:A:G | 1 | 1 | 1 |
| 14:73198105:C:G | 0 | 0 | 1 |
| 14:73198105:C:T | 0 | 0 | 1 |
| 14:73198106:T:C | 1 | 0 | 0 |
| 14:73198106:T:G | 0 | 0 | 1 |
| 14:73198110:T:G | 0 | 0 | 1 |
| 14:73198111:C:T | 0 | 0 | 1 |
| 14:73198112:C:T | 0 | 0 | 1 |
| 14:73198115:C:T | 1 | 0 | 1 |
| 14:73198117:C:G | 0 | 1 | 1 |
| 14:73198118:T:C | 0 | 0 | 1 |
| 14:73204386* | 0 | 0 | 1 |
| 14:73204758:: | 1 | 0 | 0 |
| 14:73204762** | 0 | 0 | 1 |
| 14:73206384:A:G | 0 | 0 | 1 |
| 14:73206384:A:T | 1 | 0 | 0 |
| 14:73206385:G:A | 1 | 1 | 1 |
| 14:73206385:G:T | 1 | 1 | 1 |
| 14:73206388:A:C | 0 | 0 | 1 |
| 14:73211811:A:G | 1 | 0 | 0 |
| 14:73211815:C:T | 1 | 0 | 0 |
| 14:73211830:A:G | 1 | 0 | 0 |
| 14:73211942:A:T | 1 | 0 | 1 |
| 14:73217129:G:A | 1 | 0 | 1 |
| 14:73217129:G:T | 0 | 0 | 1 |
| 14:73217137:C:G | 1 | 1 | 1 |
| 14:73217137:C:T | 1 | 1 | 1 |
| 14:73217147:G:C | 0 | 0 | 1 |
| 14:73217152:T:A | 0 | 0 | 1 |
| 14:73217153:T:C | 0 | 0 | 1 |
| 14:73217154:C:A | 0 | 0 | 1 |
| 14:73217159:T:C | 0 | 0 | 1 |
| 14:73217160:C:G | 0 | 0 | 1 |
| 14:73217161:T:C | 0 | 0 | 1 |
| 14:73217167:G:T | 0 | 0 | 1 |
| 14:73217168:T:G | 0 | 0 | 1 |
| 14:73217170:C:G | 1 | 0 | 1 |
| 14:73217171:T:C | 1 | 0 | 0 |
| 14:73217173:G:T | 1 | 0 | 0 |
| 14:73217177:G:T | 1 | 0 | 1 |
| 14:73217180:A:T | 0 | 0 | 1 |
| 14:73217182:G:A | 0 | 0 | 1 |
| 14:73217221:G:A | 1 | 0 | 0 |
| 14:73217225:G:A | 1 | 1 | 1 |
| 14:73217243:T:C | 0 | 0 | 1 |
| 14:73219139:G:C | 1 | 0 | 0 |
| 14:73219139:G:T | 0 | 0 | 1 |
| 14:73219144:T:G | 0 | 0 | 1 |
| 14:73219155:C:G | 1 | 0 | 0 |
| 14:73219155:C:T | 0 | 0 | 1 |
| 14:73219156:T:A | 0 | 0 | 1 |
| 14:73219156:T:C | 0 | 0 | 1 |
| 14:73219156:T:G | 0 | 0 | 1 |
| 14:73219161:G:C | 1 | 1 | 1 |
| 14:73219164:A:G | 1 | 0 | 0 |
| 14:73219177:C:A | 1 | 1 | 1 |
| 14:73219177:C:T | 1 | 1 | 1 |
| 14:73219182:C:T | 1 | 0 | 1 |
| 14:73219185:G:A | 0 | 0 | 1 |
| 14:73219185:GC:TG | 1 | 1 | 1 |
| 14:73219186:C:T | 1 | 0 | 0 |
| 14:73219188:C:T | 0 | 0 | 1 |
| 14:73219192:C:A | 1 | 1 | 1 |
| 14:73219194:A:G | 1 | 0 | 0 |
| 14:73219202:CACC:C | 1 | 1 | 0 |
| 14:73219262:A:G | 1 | 0 | 0 |
| 14:73221156:ACAGACAG:ACAG | 1 | 0 | 0 |
| 21:25881338:TA: | 0 | 0 | 1 |
| 21:25881777:AAACAA:AA | 1 | 0 | 0 |
| 21:25891765:T:C | 0 | 0 | 1 |
| 21:25891768:T:A | 0 | 0 | 1 |
| 21:25891777:C:A | 0 | 0 | 1 |
| 21:25891783:A:C | 0 | 1 | 0 |
| 21:25891783:T:G | 0 | 0 | 1 |
| 21:25891784:C:A | 1 | 1 | 0 |
| 21:25891784:C:A | 1 | 1 | 0 |
| 21:25891784:C:A | 1 | 1 | 0 |
| 21:25891784:C:T | 1 | 1 | 0 |
| 21:25891784:C:T | 1 | 1 | 0 |
| 21:25891784:C:T | 1 | 1 | 0 |
| 21:25891784:G:A | 0 | 0 | 1 |
| 21:25891784:G:C | 0 | 0 | 1 |
| 21:25891784:G:T | 0 | 0 | 1 |
| 21:25891787:A:G | 0 | 1 | 0 |
| 21:25891787:A:T | 0 | 0 | 1 |
| 21:25891787:TC:CA | 1 | 0 | 0 |
| 21:25891789:T:C | 0 | 0 | 1 |
| 21:25891790:C:T | 0 | 1 | 0 |
| 21:25891790:G:A | 0 | 0 | 1 |
| 21:25891792:C:T | 0 | 0 | 1 |
| 21:25891792:G:A | 1 | 1 | 0 |
| 21:25891793:A:G | 0 | 0 | 1 |
| 21:25891793:T:C | 1 | 1 | 0 |
| 21:25891796:C:T | 1 | 1 | 0 |
| 21:25891808:C:T | 1 | 0 | 0 |
| 21:25891820:C:G | 0 | 0 | 1 |
| 21:25891820:G:C | 1 | 1 | 0 |
| 21:25891848:TTCTTTGCAGAAGATGTG: | 0 | 0 | 1 |
| 21:25891853:C:T | 1 | 1 | 0 |
| 21:25891853:G:A | 0 | 0 | 1 |
| 21:25891854:GAA: | 0 | 0 | 1 |
| 21:25891855:A:G | 0 | 0 | 1 |
| 21:25891855:T:C | 1 | 1 | 0 |
| 21:25891856:C:G | 1 | 1 | 0 |
| 21:25891856:C:G | 1 | 1 | 0 |
| 21:25891856:G:A | 0 | 0 | 1 |
| 21:25891856:G:C | 0 | 0 | 1 |
| 21:25891858:C:G | 0 | 0 | 1 |
| 21:25891858:G:C | 1 | 1 | 0 |
| 21:25897575:T:G | 0 | 0 | 1 |
| 21:25897578:A:C | 0 | 0 | 1 |
| 21:25897605:G:A | 0 | 0 | 1 |
| 21:25897605:G:C | 0 | 0 | 1 |
| 21:25897619:G:A | 1 | 1 | 0 |
| 21:25897620:C:T | 0 | 1 | 0 |
| 21:25897626:TC:GA | 1 | 1 | 0 |
| 21:25897627:G:T | 0 | 0 | 1 |
| 21:25897642:C:G | 0 | 1 | 0 |
| 21:25911840:C:T | 1 | 0 | 0 |
| 21:25911961:A:G | 1 | 0 | 0 |
| 21:25975060:C:T | 1 | 0 | 0 |
| 21:26000066:G:A | 1 | 0 | 0 |
| 21:26000131:C:T | 1 | 0 | 0 |
| 21:26000163:G:A | 1 | 0 | 0 |
| 21:26021865:GTGGTGGTGGTGGTGGTGGTGG:GTGGTGGTGGTGGTGGTGGTGGTGG | 1 | 0 | 0 |
| 21:26021963:C:T | 1 | 0 | 0 |
| 21:26022001:G:A | 1 | 0 | 0 |
| 21:26022051:G:T | 1 | 0 | 0 |
| 21:26051044:C:T | 1 | 0 | 0 |
| 21:26051070:A:G | 1 | 0 | 0 |
| 21:26171246:G:A | 1 | 0 | 0 |

*14:73204386:TAAAAAAATATTATTTTAGGACTATTTAACAAATATACTATTTATGATAATTAACATGGGGTTAGCAAACTACTGCTTGAAGCCAAATATGGCCCATGGCTTGTTTTTGTACAGCTTATAAGCTAAAAATGCTTTTTACATTTAAAAAAAAAAAAAAAGAACAAGGAAGAATATGTGACACACAAAACCTGAAATGTTTACTCCCTGGCCATTTACAGAAAAAGTTAGCTGGACATTGATTTTCATGTTACTATTAACTGCTAAAACAAAGGAGCTTGTAGCTATATTTTTTCATACTTGCATAAAGAAAGAAAACCTAATAAATCGCTGGTGGCTTCATTTCCAAAAGGCTATGGAAATCCATAAACAGGCTGGGTGCAGTGGCTCATGCCTGTAATCCTAGCACTTTGGGAGGCTGAGGCGGGAGGATTGCTTGAGCCCAGGAGTTTGAGTCTGGCCTGGGCAACGTAGTGAGACCCTGTTTTTAGAAAAAATGAAGATAAAAAACTAATCCATAAACCTCATACTTGGTGGTTGCCCTTCAGTTTTTCTTCTTTAGATCTGGATTTAGTTAAACAGTGTATTGCCTGCCTGGTTTCTGTTAACTTTTGATCATTTGGTTGGGTTATCCTATAATGAAACTTGGTTGACCTCAGAAATAACAAGGTGGTCGTTCAGCTTCTTTGATTGTGTGTTTCTTTCATAAGTTCTTCAAGAAGCCTGAGTATTAGAAACATGGATAAAATTATTGTGATAAAAAGCCAGAGAGACATAAATGTCAAGTATCTTGTTTAAAATTACTGTGACCAGGCCGGGCGTGGTGGCTCACGCCTGTAATCCCAGCACTTTGGGAGGCTGAGGCGGGCGGATCACGAGGTCAGGAGATCGAGACCATCCTGGCTAACACAGTGAAACCCCACCTCTACTAAAAAAAATACAAAAAATTAGCCAGGCATGGTGGCGGGCGCCTGTAGTCCCAGCTACTCGGGAGGCTGAGGCAGGAGAATGGCGTGAACCCAGGAGACGGAGCTTGCAGTGAGCCGAGATCATGCCACTGTACTCCAGCCTGGGTGACAGAGCAAGACTCTGTCTCAAAAAAAAAAAAAAAAAAAAAAATTACTGTGACCAGATTGGACTCAGCTTGCTGCTGGCATTTGGTTCCCACCATAACCTCATATGTCATGTGTTTGCTTATATGTACTTTTGTGTTATTGTTGGTGTATCTTCAGGATAAGTTCCAAAATGTAATATTGCTGGGTTAAAGGATTAATGCACATGTAGTTTTATTAGATGTTACCAAATTTCCCTCCAGTGGGGATTATACCATTTTTCATTCCTGCCTGAAATATATGAGAGAAGCACCATTTTAAAGTTTTCACAATGTCTCTGAACTAAAATGTGGTAGAGATGCACATGTGTATATCTAGATCTAGATTGATATATTGATATACATGTATTTTTGTAGAAGGGGCACAGAGGAGGCCTCTTGACTATCCCTATTATGTTTATTGCTTTTAACGTTTTTTCTCACGCCGGGTTTAAAACATGGTTTAGCATTATTGAACATTCTAAAAATGAGACATAATATGAAGGAAATTTACTCTGTGCATCTTTTGAATTATAATCACCATCTGAGGCTTTTGTGAGCTCCAGTTTGTCCTGGAATTTAGAACATTCAACTAGTCCAGCTATTGTTTCAGTGGAATCTTGCTGGCCTGAACAGTTTTCCTTCTGGTTCCTTTTCAGGGACTAAAAATGATGAAGAGTTTTGTGATAGCAGGTGCAGTTTGAGTACTACAGTAAACATTCAGTTTCAGAACTTCTTCTCTTACCTGCTAAAACCAAAGAGAACCTTTTTTTTATTTTTACTTCTGATTGTTGAACAGTCTTAAGGCAGCATTAGGAAGACTGGCGATTTGTGTGGAGAAATGATGGCTTGTTGTTGTCTATGCATACTTTGTGTGTCCAGTGCTTACCTGGAATTTTGTCTTTCCCAACAGCAACAATGGTGTGGTTGGTGAATATGGCAGAAGGAGACCCGGAAGCTCAAAGGAGAGTATCCAAAAATTCCAAGTATAATGCAGAAAGTAGGTAACTTTTATTAGATAATATCTTGATTTTTCAGGGTCACTGTTATAAGCTAACAGTATAGCAATGTTTTTATCGTCTTTCTTTGGTCATAGACTCCTTTGAGAATCTCTTGAGAACTATGATAATGCCCAGTAAATACACAGATAAGTATTTAAGGAGTTCAGATACTCAAAACCCAACAATACAGTCAAAGCATCCTAGGTTAAGACACCTCCCATTAAATACAGAATACCAGCATGGAAAGGTTCAGGCTGAGGTTATGATTTTGTTTTTGTTTTTTGTTTGTTTTTTATAAGTCATGATTTTAAAAAGAAAAAATAAACTCTCTCCAAACATGTAAAAGTAAGAATCTCCTAAAAGAAACAAAAAAGAAACAGACAATAAAGGAAAAATAAGTAAACAAAAAAAGCAAAATATAAACAACATAAAAATGAGAACCTCTTGTCTAACATGGCCAGAATAACTGAAATATGATTTGAGATGTGCTGATGTGTATACTGAGAAGATCAAATATTCTAGGTGGATATGACTTTTTAGAAGAGGATAAGAATGACTTAGGATGAGCTGGGTGCAGTGATGTGTGCCTGTAGTCCCAGCTACATGGCCAAGGCAAGAGGATCACTTGAGCTAAGGAGTTCCGGATTACCCTGGGCAACTTATACCTTACCTCAAAAATAATAATAATAATGAATAATAATAATGACTTAGGATAAGAGAGTGAAAGGCCCTCTAAGGATACTACAGAACATATTAAGGAGAACACAAGTTAGAAGTTGTTATTAGATGGAGTTGAGGGGGAGCTTTCATTCAGTTTTCTGGAATGGAGGCTACCTTCCCACACACACTAAAAAAAAAAAGAAAGGGTTGAGGAATAACAGCACAATATAAATAGTCTAAAAAAAGGGCTAACTTTTATGAGGAATCTGAAAAACAACAGGCACAGTAAATATTGGTGAGGAAAAAAAGGCCTTAATTGAGTTGCTGTTTGTCAGGAAAGGGAGAGTTTATGTTTTATAAAGCCAGGAAATCTACAGAGTTAGCATTTAGGCAACTGCAAGGAAGATACTTTCAACCAGGAATTCCTATTTGGAACTCAGAGATGTGAGTCTTTATTATACAGTAAGCAAGGCAGCAGAACAAGGTCATGTTAGACTCTCACGGGATCCTTAGGGTGTGACTTTGCTAGCCAGAAACCTCTGTGGCTGGTGGCACCTTTTCCTGAGATTTGCTCAGGCCCACTGGGCTCATTCCACCTACTCAGCCTAGCTGGCTATGCTTGGCTTTCTCTACCAGCTGGACCCCATGCCTGCCAAAGGCAAGCCAGGCATGGAGCAGCGAGGAGTGCATGAGCAAGTGAGTGCAGGGTCCAGCCACTGCACACAGCCAGGCATGCTGGCTGCAGCAGGGCAGGCAGCCTCAGGTACTAGCTCCCTGCTGCAGCTAGACCAGGCATTACCTAAGCAGCTTTGACTACAGGCCACTACCACTAGTAAACCTGATAGAAAAGTAAGAAATATACTGGAAAGAATTGCCACAAGCCTGGCTCTTGTGTTAACCTTGTGTTAACCTGGGGAACACAGGGGCACCCAGAAGCTTGGAGATGCCAGGAACTGCAGAACCCCAAAGAGGGGGGTCATAGCCCTGGCTTAGGGAACTCCTGAGTCTGGACTCCCCGAAGGGCCACATCTCTTCACTCCTCTCTTCTCTCCTTCTCGCCACCTGCAACGTGGCAAGTGGGGGGCACGTTTCAGCCCTGTTTGTCTTACGGTTCTTTCAATCCTGCCATTCAATGGGTCCCAAGTTCTTGTCCTGCATCCAGGAAGAATGAAGTACGTGGACAGCTGGAGGGAGAGCAAGATGAAAAGGTGCTTTATTGAGCAACAGTACAGCTCTCAGGAGACCCTATCTGCAGGCAGGTCGTCCCATTGTCTCTGCAGCTCTCAGTGGAGAGGAGACCTGGAGTGGGTAGCTCCTGTCTGCAGGCAGGTCATCCTGATGTCTGTAATCCTCAGTGGAGAGGAGACCCGGAATGGGTATCTCCTATCCACAAGCAGGTCATCCCATCATCTCTGCAGCCCTCAGTGGAGAGGAGACCCGGAGTGGGTAGCTTCTTTCTGCAGGCAGGTTGTCCCTTCATCTGCCCAAGTCTGACTGAGTCTGGGGTTTTTATGGGCTTCAGAGGGGAGGAAGTACATGCTGATGGGTCCATGGGTGGCCATGAGCAGGCCCAGAAAATGCACCATAAGTTCTCACTGTGGTCTGTGGAACTGGGAGCTTGGCCTCCAGGCTTCAGGCTTTCCCTGACTTGAGGTGGTGCTTCACCAGGGAACTTCCCCTTTCCGCCTAGGAGCCTGTCTGCCTCCTGCTGCCATCAACCTGCTGTCTATAGCTCCCATGGCACCCAGGCTGTTCATGCCAAAGGGTGCCTGCAGGCCCACGGTAAGCTGCCTTCAGCCCCTCCTCAGTCTCCCTCCCATGCTTGTCAGCACCCAAAGTCCAGAGGGGGCCAAGGTGGCAGGGGGCTGGCATGTCAGTGCTGCCCCAAGCGCCCGCACACCCGGCCAGGTCATGACAGCACCCAGCCTCAGCCACAACTTTGCTCTGAAATCGGAGCCTGCAGACGCAAGGGGCTTCCTGGACCCCTGAGAGCACAGGGATGCCCAGGTCTACAGCCACAGCTGGGAAGCTGCAGCTGTGGGAGTGCGGGACTTCTGCCCCACCAACTCAGAAGGGGGCAGGACTCCCGCCTGTTCCTGGCTCCTGCCAGCTCACAGAGCATGCTGCCCCAGCTGCACCTCCCCTGCTGCAGCTGCCATCTTTGCAGCAGCCACTCCAGATGGGCCGCTGCTGCCATCAAGACCACTAGTAAACCTGATAGAAAAGTAAGAAATATACTGGAAAGAATTGCCACAAGCCTGGCTCTTGTGTTAACCTGTGTTAAGCTAAAGAAAATCAAATGATTGTCTGTGAGCATGTAGGTATATATGTATGTGAGTGAGTGATACAACAGAATTTCATTCATTTTACAGATGTTGACTGAGCACCTGACTATGTGCTAGGCCCTGGGGATATAGCACTGAACAAGATTTCCCCTGCCCTTGTGGAGCTTATAGTCTATTTGGAGAGATAGATGGTCAACAAATTATTACATAAATAATTCATACAGTTGTGATAGGTACTACAAAGAAGACGTATAAGTTGCTATGAAAGTTTATAATAGGGGAATTTTACGTATCCTGGAAAGTCAAGGGGTGCTTCCCTGAGGAAGTGGTAATAGGGGACGGCCCGCTGAAGGATGGAGGAAGAGCTTTCCAAGAGAGGGACAACATGAGCAAGGGCTTTGAAATGAGAAGGCTGGATGAACTGCAGGCTTCCTCAGTGAGAATGCTGCTGGTATTTTGGGGGGCACAGTTCTTTGTGGGACTTTCCCTCATATTGCAAGATATTTAACATCCCTGGCCCCTCACCCACTAAATGCCAATAATGGCTTTAAGGCTTTGCAATAATCAAAAACTCCTCCATCCTAGTTATTTCCAAACACTCCCCGGAAGGGAGGTGCTATCCCTAGTTGAAAAATCACTGTGTTAACGGAACTAGAAGTTACATTGGAACAAAAGGGCATAGGGCTCCAAGAGGGATATCTGTGTAAGGAAAAAACAAACAAAAGAACTGAGAGATTACCTGATGTGGTTGAGCTCTGTCAGAGCATTTCAGGATAAATTAGTCATAGATAATAATATAAAATTCATCAGTGGAAAAAAATGAGG:None

**14:73204762:TGCAGTGGCTCATGCCTGTAATCCTAGCACTTTGGGAGGCTGAGGCGGGAGGATTGCTTGAGCCCAGGAGTTTGAGTCTGGCCTGGGCAACGTAGTGAGACCCTGTTTTTAGAAAAAATGAAGATAAAAAACTAATCCATAAACCTCATACTTGGTGGTTGCCCTTCAGTTTTTCTTCTTTAGATCTGGATTTAGTTAAACAGTGTATTGCCTGCCTGGTTTCTGTTAACTTTTGATCATTTGGTTGGGTTATCCTATAATGAAACTTGGTTGACCTCAGAAATAACAAGGTGGTCGTTCAGCTTCTTTGATTGTGTGTTTCTTTCATAAGTTCTTCAAGAAGCCTGAGTATTAGAAACATGGATAAAATTATTGTGATAAAAAGCCAGAGAGACATAAATGTCAAGTATCTTGTTTAAAATTACTGTGACCAGGCCGGGCGTGGTGGCTCACGCCTGTAATCCCAGCACTTTGGGAGGCTGAGGCGGGCGGATCACGAGGTCAGGAGATCGAGACCATCCTGGCTAACACAGTGAAACCCCACCTCTACTAAAAAAAATACAAAAAATTAGCCAGGCATGGTGGCGGGCGCCTGTAGTCCCAGCTACTCGGGAGGCTGAGGCAGGAGAATGGCGTGAACCCAGGAGACGGAGCTTGCAGTGAGCCGAGATCATGCCACTGTACTCCAGCCTGGGTGACAGAGCAAGACTCTGTCTCAAAAAAAAAAAAAAAAAAAAAAATTACTGTGACCAGATTGGACTCAGCTTGCTGCTGGCATTTGGTTCCCACCATAACCTCATATGTCATGTGTTTGCTTATATGTACTTTTGTGTTATTGTTGGTGTATCTTCAGGATAAGTTCCAAAATGTAATATTGCTGGGTTAAAGGATTAATGCACATGTAGTTTTATTAGATGTTACCAAATTTCCCTCCAGTGGGGATTATACCATTTTTCATTCCTGCCTGAAATATATGAGAGAAGCACCATTTTAAAGTTTTCACAATGTCTCTGAACTAAAATGTGGTAGAGATGCACATGTGTATATCTAGATCTAGATTGATATATTGATATACATGTATTTTTGTAGAAGGGGCACAGAGGAGGCCTCTTGACTATCCCTATTATGTTTATTGCTTTTAACGTTTTTTCTCACGCCGGGTTTAAAACATGGTTTAGCATTATTGAACATTCTAAAAATGAGACATAATATGAAGGAAATTTACTCTGTGCATCTTTTGAATTATAATCACCATCTGAGGCTTTTGTGAGCTCCAGTTTGTCCTGGAATTTAGAACATTCAACTAGTCCAGCTATTGTTTCAGTGGAATCTTGCTGGCCTGAACAGTTTTCCTTCTGGTTCCTTTTCAGGGACTAAAAATGATGAAGAGTTTTGTGATAGCAGGTGCAGTTTGAGTACTACAGTAAACATTCAGTTTCAGAACTTCTTCTCTTACCTGCTAAAACCAAAGAGAACCTTTTTTTTATTTTTACTTCTGATTGTTGAACAGTCTTAAGGCAGCATTAGGAAGACTGGCGATTTGTGTGGAGAAATGATGGCTTGTTGTTGTCTATGCATACTTTGTGTGTCCAGTGCTTACCTGGAATTTTGTCTTTCCCAACAGCAACAATGGTGTGGTTGGTGAATATGGCAGAAGGAGACCCGGAAGCTCAAAGGAGAGTATCCAAAAATTCCAAGTATAATGCAGAAAGTAGGTAACTTTTATTAGATAATATCTTGATTTTTCAGGGTCACTGTTATAAGCTAACAGTATAGCAATGTTTTTATCGTCTTTCTTTGGTCATAGACTCCTTTGAGAATCTCTTGAGAACTATGATAATGCCCAGTAAATACACAGATAAGTATTTAAGGAGTTCAGATACTCAAAACCCAACAATACAGTCAAAGCATCCTAGGTTAAGACACCTCCCATTAAATACAGAATACCAGCATGGAAAGGTTCAGGCTGAGGTTATGATTTTGTTTTTGTTTTTTGTTTGTTTTTTATAAGTCATGATTTTAAAAAGAAAAAATAAACTCTCTCCAAACATGTAAAAGTAAGAATCTCCTAAAAGAAACAAAAAAGAAACAGACAATAAAGGAAAAATAAGTAAACAAAAAAAGCAAAATATAAACAACATAAAAATGAGAACCTCTTGTCTAACATGGCCAGAATAACTGAAATATGATTTGAGATGTGCTGATGTGTATACTGAGAAGATCAAATATTCTAGGTGGATATGACTTTTTAGAAGAGGATAAGAATGACTTAGGATGAGCTGGGTGCAGTGATGTGTGCCTGTAGTCCCAGCTACATGGCCAAGGCAAGAGGATCACTTGAGCTAAGGAGTTCCGGATTACCCTGGGCAACTTATACCTTACCTCAAAAATAATAATAATAATGAATAATAATAATGACTTAGGATAAGAGAGTGAAAGGCCCTCTAAGGATACTACAGAACATATTAAGGAGAACACAAGTTAGAAGTTGTTATTAGATGGAGTTGAGGGGGAGCTTTCATTCAGTTTTCTGGAATGGAGGCTACCTTCCCACACACACTAAAAAAAAAAAGAAAGGGTTGAGGAATAACAGCACAATATAAATAGTCTAAAAAAAGGGCTAACTTTTATGAGGAATCTGAAAAACAACAGGCACAGTAAATATTGGTGAGGAAAAAAAGGCCTTAATTGAGTTGCTGTTTGTCAGGAAAGGGAGAGTTTATGTTTTATAAAGCCAGGAAATCTACAGAGTTAGCATTTAGGCAACTGCAAGGAAGATACTTTCAACCAGGAATTCCTATTTGGAACTCAGAGATGTGAGTCTTTATTATACAGTAAGCAAGGCAGCAGAACAAGGTCATGTTAGACTCTCACGGGATCCTTAGGGTGTGACTTTGCTAGCCAGAAACCTCTGTGGCTGGTGGCACCTTTTCCTGAGATTTGCTCAGGCCCACTGGGCTCATTCCACCTACTCAGCCTAGCTGGCTATGCTTGGCTTTCTCTACCAGCTGGACCCCATGCCTGCCAAAGGCAAGCCAGGCATGGAGCAGCGAGGAGTGCATGAGCAAGTGAGTGCAGGGTCCAGCCACTGCACACAGCCAGGCATGCTGGCTGCAGCAGGGCAGGCAGCCTCAGGTACTAGCTCCCTGCTGCAGCTAGACCAGGCATTACCTAAGCAGCTTTGACTACAGGCCACTACCACTAGTAAACCTGATAGAAAAGTAAGAAATATACTGGAAAGAATTGCCACAAGCCTGGCTCTTGTGTTAACCTTGTGTTAACCTGGGGAACACAGGGGCACCCAGAAGCTTGGAGATGCCAGGAACTGCAGAACCCCAAAGAGGGGGGTCATAGCCCTGGCTTAGGGAACTCCTGAGTCTGGACTCCCCGAAGGGCCACATCTCTTCACTCCTCTCTTCTCTCCTTCTCGCCACCTGCAACGTGGCAAGTGGGGGGCACGTTTCAGCCCTGTTTGTCTTACGGTTCTTTCAATCCTGCCATTCAATGGGTCCCAAGTTCTTGTCCTGCATCCAGGAAGAATGAAGTACGTGGACAGCTGGAGGGAGAGCAAGATGAAAAGGTGCTTTATTGAGCAACAGTACAGCTCTCAGGAGACCCTATCTGCAGGCAGGTCGTCCCATTGTCTCTGCAGCTCTCAGTGGAGAGGAGACCTGGAGTGGGTAGCTCCTGTCTGCAGGCAGGTCATCCTGATGTCTGTAATCCTCAGTGGAGAGGAGACCCGGAATGGGTATCTCCTATCCACAAGCAGGTCATCCCATCATCTCTGCAGCCCTCAGTGGAGAGGAGACCCGGAGTGGGTAGCTTCTTTCTGCAGGCAGGTTGTCCCTTCATCTGCCCAAGTCTGACTGAGTCTGGGGTTTTTATGGGCTTCAGAGGGGAGGAAGTACATGCTGATGGGTCCATGGGTGGCCATGAGCAGGCCCAGAAAATGCACCATAAGTTCTCACTGTGGTCTGTGGAACTGGGAGCTTGGCCTCCAGGCTTCAGGCTTTCCCTGACTTGAGGTGGTGCTTCACCAGGGAACTTCCCCTTTCCGCCTAGGAGCCTGTCTGCCTCCTGCTGCCATCAACCTGCTGTCTATAGCTCCCATGGCACCCAGGCTGTTCATGCCAAAGGGTGCCTGCAGGCCCACGGTAAGCTGCCTTCAGCCCCTCCTCAGTCTCCCTCCCATGCTTGTCAGCACCCAAAGTCCAGAGGGGGCCAAGGTGGCAGGGGGCTGGCATGTCAGTGCTGCCCCAAGCGCCCGCACACCCGGCCAGGTCATGACAGCACCCAGCCTCAGCCACAACTTTGCTCTGAAATCGGAGCCTGCAGACGCAAGGGGCTTCCTGGACCCCTGAGAGCACAGGGATGCCCAGGTCTACAGCCACAGCTGGGAAGCTGCAGCTGTGGGAGTGCGGGACTTCTGCCCCACCAACTCAGAAGGGGGCAGGACTCCCGCCTGTTCCTGGCTCCTGCCAGCTCACAGAGCATGCTGCCCCAGCTGCACCTCCCCTGCTGCAGCTGCCATCTTTGCAGCAGCCACTCCAGATGG:None

**Supplementary Table e-3. Distribution of AD Diagnosis by Gene Among the Carriers.**

| Gene |  | Case  (%) | Control  (%) | Control  > 65 years (%) | Control  >85 years (%) |
| --- | --- | --- | --- | --- | --- |
| *PSEN2* | Clinical Variant Carriers | 4 (0.03%) | 1 (0.01%) | 1 (0.01%) | 1 (0.02%) |
|  | Conflicting Clinical Variant Carriers | 95 (0.7%) | 118 (0.8%) | 115 (0.8%) | 50 (1.1%) |
|  | Additional DM Variant Carriers | 167 (1.2%) | 180 (1.2%) | 169 (1.2%) | 54 (1.1%) |
| *PSEN1* | Clinical Variant Carriers | 56 (0.4%) | 2 (0.01%) | 2 (0.01%) | 0 (0%) |
|  | Conflicting Clinical Variant Carriers | 40 (0.3%) | 40 (0.3%) | 52 (0.3%) | 10 (0.2%) |
|  | Additional DM Variant Carriers | 39 (0.3%) | 29 (0.2%) | 25 (0.2%) | 3 (0.06%) |
| *APP* | Clinical Variant Carriers | 2 (0.01%) | 0 (0%) | 0 (0%) | 0 (0%) |
|  | Conflicting Clinical Variant Carriers | 89 (0.6%) | 112 (0.8%) | 110 (0.8%) | 38 (0.8%) |
|  | Additional DM Variant Carriers | 159 (1.2%) | 199 (1.4%) | 174 (1.3%) | 45 (1.0%) |
| Total |  | 13,825 | 14,715 | 13,644 | 4722 |

Data shown as frequency (percentage).

DM: Damaging missense.

**Supplementary Table e-4: List of Conflicting Clinical Variants with Alternative Alleles Observed within the ADSP.**

| **Gene** | **Chr:Position** | **rsID** | **REF** | **ALT** | **Protein Change** | **ClinVar** | **OMIM** | **Alzforum** | **gnomadAF*** | **Intervar Pathogenicity**** | **Control** | **Case** | **Unknown** |
| --- | --- | --- | --- | --- | --- | --- | --- | --- | --- | --- | --- | --- | --- |
| ***PSEN2*** | 1:226883729 | rs188598190 | G | A | Gly56Ser | 1 | 0 | 0 | 2.00E-04 | UnS | 5 | 1 | 0 |
|  | 1:226883771 | rs139972151 | G | A | Gly70Arg | 1 | 0 | 0 | 3.23E-05 | UnS | 0 | 0 | 1 |
|  | 1:226883863 | rs200801915 | C | T | Ile100= | 1 | 0 | 0 | 3.24E-05 | LB | 0 | 2 | 1 |
|  | 1:226883899 | rs200610057 | C | T | Tyr112= | 1 | 0 | 0 | 2.00E-04 | LB | 12 | 11 | 1 |
|  | 1:226885570 | rs63750197 | C | T | Ser130Leu | 0 | 1^#^ | 0 | 5.00E-04 | UnS | 26 | 28 | 6 |
|  | 1:226885596 | rs202178897 | G | A | Val139Met | 1 | 0 | 0 | 3.24E-05 | UnS | 8 | 3 | 2 |
|  | 1:226885629 | rs866044092 | G | A | Val150Met | 1 | 0 | 0 | 3.24E-05 | UnS | 1 | 0 | 0 |
|  | 1:226888952 | rs145010538 | C | G | Ala230= | 1 | 0 | 0 | 2.00E-04 | LB | 13 | 20 | 5 |
|  | 1:226888973 | rs202003390 | G | A | Ala237= | 1 | 0 | 0 |  | LB | 1 | 1 | 1 |
|  | 1:226888974 | rs367855127 | C | T | Leu238Phe | 1 | 0 | 0 | 3.23E-05 | UnS | 0 | 1 | 0 |
|  | 1:226889016 | rs138836272 | G | A | Ala252Thr | 1 | 0 | 0 | 5.00E-04 | UnS | 11 | 1 | 2 |
|  | 1:226891345 | rs199587016 | C | T | Pro318= | 1 | 0 | 0 | 2.00E-04 | LB | 14 | 9 | 4 |
|  | 1:226895699 | rs143059995 | G | A |  | 1 | 0 | 0 |  | NR | 28 | 18 | 4 |
| ***PSEN1*** | 14:73170989 | rs63750831 | G | A | Val94Met | 0 | 0 | 1^#^ |  | UnS | 1 | 0 | 0 |
|  | 14:73192749 | rs115760359 | A | G | Pro218= | 1 | 0 | 0 | 2.00E-04 | LB | 1 | 0 | 1 |
|  | 14:73211811 | rs121917809 | A | G | Asp333Gly | 1 | 0 | 0 | 3.00E-04 | LP | 5 | 7 | 0 |
|  | 14:73211815 | rs116640707 | C | T | Gly334= | 1 | 0 | 0 | 3.00E-04 | LB | 10 | 5 | 1 |
|  | 14:73211830 | rs201776669 | A | G | Glu339= | 1 | 0 | 0 | 2.00E-04 | LB | 19 | 25 | 6 |
|  | 14:73217221 | rs63750227 | G | A | Ala409Thr | 1 | 0 | 0 |  | UnS | 1 | 0 | 0 |
|  | 14:73219164 |  | A | G | Ile427Val | 1 | 0 | 0 |  | UnS | 0 | 1 | 0 |
|  | 14:73219194 | rs764971634 | A | G | Ile437Val | 1 | 0 | 0 | 2.00E-04 | UnS | 0 | 1 | 0 |
|  | 14:73219262 | rs201496204 | A | G | Gln459= | 1 | 0 | 0 | 3.23E-05 | LB | 3 | 2 | 0 |
|  | 21:25891796 | rs63750066 | C | T | Ala713Thr | 1 | 1 | 0 | 3.23E-05 | UnS | 0 | 1 | 0 |
| ***APP*** | 21:25891808 | rs201269325 | C | T | Gly709Ser | 1 | 0 | 0 | 6.47E-05 | LP | 1 | 0 | 0 |
|  | 21:25897620 | rs63750847 | C | T | Ala673Thr | 0 | 1^#^ | 0 | 8.00E-04 | UnS | 5 | 1 | 0 |
|  | 21:25897642 | rs63750363 | C | G | Glu665Asp | 0 | 1^#^ | 0 |  | UnS | 2 | 3 | 1 |
|  | 21:25911840 | rs199887707 | C | T | Val604Met | 1 | 0 | 0 | 1.10E-03 | UnS | 4 | 1 | 1 |
|  | 21:25911961 | rs137865262 | A | G | Asp563= | 1 | 0 | 0 | 6.46E-05 | LB | 9 | 5 | 3 |
|  | 21:25975060 | rs201290605 | C | T |  | 1 | 0 | 0 |  | NR | 5 | 2 | 3 |
|  | 21:26000066 | rs200978018 | G | A | Arg328Trp | 1 | 0 | 0 | 1.00E-04 | UnS | 4 | 6 | 4 |
|  | 21:26000131 | rs202218688 | C | T | Arg306His | 1 | 0 | 0 | 3.23E-05 | UnS | 2 | 1 | 1 |
|  | 21:26000163 | rs199890425 | G | A | Ala295= | 1 | 0 | 0 |  | LB | 3 | 1 | 0 |
|  | 21:26021963 | rs200103591 | C | T | Asp248Asn | 1 | 0 | 0 | 4.00E-04 | UnS | 5 | 1 | 0 |
|  | 21:26022001 | rs139819006 | G | A | Ala235Val | 1 | 0 | 0 |  | UnS | 3 | 1 | 0 |
|  | 21:26022051 | rs199587668 | G | T |  | 1 | 0 | 0 |  | NR | 9 | 9 | 1 |
|  | 21:26051044 | rs201022619 | C | T | Ser206= | 1 | 0 | 0 | 3.23E-05 | LB | 3 | 1 | 0 |
|  | 21:26051070 | rs145081708 | A | G | Ser198Pro | 1 | 0 | 0 | 2.00E-04 | UnS | 23 | 22 | 3 |
|  | 21:26171246 |  | G | A |  | 1 | 0 | 0 |  | NR | 35 | 36 | 4 |

*Allele frequency from wInterVar annotation using the gnomAD exomes controls AF.

**UnS, Uncertain significance; NR, Not Reported; LB, Likely Benign, LP, Likely, Pathogenic.

^#^Variants have conflicting pathogenicity interpretation among the 3 databases.

**Supplementary Table e-5: List of Damaging Missense Variants with Alternative Alleles Observed within the ADSP**

| **Gene** | **Chr:Position** | **rsID** | **REF** | **ALT** | **Protein Change** | **gnomadAF*** | **Control** | **Case** | **Unknown** |
| --- | --- | --- | --- | --- | --- | --- | --- | --- | --- |
| ***PSEN2*** | 1:226881915 | rs1261261196 | C | A | Thr3Lys | 7.95E-06 | 0 | 2 | 0 |
|  | 1:226881945 | rs766853710 | T | C | Val13Ala |  | 1 | 0 | 0 |
|  | 1:226881956 | rs199644116 | C | T | Arg17Trp | 2.39E-05 | 1 | 0 | 0 |
|  | 1:226881960 | rs143061887 | C | T | Thr18Met | 1.59E-05 | 4 | 1 | 0 |
|  | 1:226881977 | rs757152552 | G | A | Glu24Lys | 1.19E-05 | 1 | 0 | 0 |
|  | 1:226881990 | rs748504448 | C | T | Pro28Leu | 3.98E-06 | 1 | 1 | 1 |
|  | 1:226881993 | rs771375988 | G | A | Arg29His | 1.20E-05 | 1 | 0 | 0 |
|  | 1:226882017 | rs1231154581 | G | A | Gly37Asp | 3.99E-06 | 1 | 1 | 1 |
|  | 1:226882026 | rs1306395711 | A | T | Asp40Val | 3.99E-06 | 1 | 0 | 0 |
|  | 1:226883708 |  | A | G | Ser49Gly |  | 0 | 1 | 0 |
|  | 1:226883712 | rs143501870 | A | G | Gln50Arg | 0.0002237 | 1 | 2 | 1 |
|  | 1:226883725 | rs146894466 | G | C | Glu54Asp | 0.0002233 | 24 | 11 | 1 |
|  | 1:226883729 | rs188598190 | G | A | Gly56Ser | 0.0001874 | 5 | 1 | 0 |
|  | 1:226883747 | rs150400387 | C | T | Arg62Cys | 0.0002111 | 6 | 8 | 0 |
|  | 1:226883757 | rs766446160 | G | A | Cys65Tyr | 4.38E-05 | 0 | 0 | 1 |
|  | 1:226883768 | rs202133351 | C | G | Pro69Ala | 9.15E-05 | 2 | 1 | 0 |
|  | 1:226883771 | rs139972151 | G | A | Gly70Arg | 6.37E-05 | 0 | 0 | 1 |
|  | 1:226883774 | rs140501902 | C | T | Arg71Trp | 0.003573 | 65 | 79 | 17 |
|  | 1:226883775 | rs769031756 | G | A | Arg71Gln | 2.39E-05 | 1 | 0 | 1 |
|  | 1:226883799 | rs760961297 | T | C | Leu79Pro | 1.19E-05 | 1 | 1 | 0 |
|  | 1:226883804 | rs1405799988 | C | T | Leu81Phe | 3.98E-06 | 0 | 1 | 0 |
|  | 1:226883817 | rs63750048 | C | T | Ala85Val | 3.98E-06 | 1 | 0 | 0 |
|  | 1:226883855 |  | T | G | Cys98Gly |  | 0 | 0 | 0 |
|  | 1:226883862 |  | T | A | Ile100Asn |  | 1 | 0 | 0 |
|  | 1:226883904 |  | A | G | Glu114Gly |  | 1 | 0 | 0 |
|  | 1:226885543 | rs1325639403 | A | G | Tyr121Cys | 3.99E-06 | 0 | 1 | 0 |
|  | 1:226885551 | rs1345586077 | T | C | Phe124Leu | 3.98E-06 | 0 | 1 | 0 |
|  | 1:226885564 | rs752002301 | C | T | Thr128Ile | 7.97E-06 | 0 | 1 | 0 |
|  | 1:226885570 | rs63750197 | C | T | Ser130Leu | 0.000677 | 26 | 28 | 6 |
|  | 1:226885596 | rs202178897 | G | A | Val139Met | 0.0001155 | 8 | 3 | 2 |
|  | 1:226885603 | rs63750215 | A | T | Asn141Ile |  | 0 | 2 | 0 |
|  | 1:226885617 | rs1004970794 | A | T | Ile146Phe | 3.98E-06 | 1 | 1 | 2 |
|  | 1:226885629 | rs866044092 | G | A | Val150Met | 0 | 1 | 0 | 0 |
|  | 1:226885635 | rs1410810723 | A | G | Met152Val | 4.00E-06 | 0 | 2 | 0 |
|  | 1:226885668 | rs200931244 | C | T | Arg163Cys | 8.09E-06 | 0 | 2 | 0 |
|  | 1:226888112 | rs61757781 | A | G | Met174Val | 0.0006164 | 12 | 5 | 2 |
|  | 1:226888114 |  | G | A | Met174Ile |  | 0 | 1 | 0 |
|  | 1:226888136 | rs1343351444 | C | T | Leu182Phe |  | 1 | 0 | 0 |
|  | 1:226888851 | rs770238439 | G | A | Val197Met | 1.20E-05 | 1 | 0 | 0 |
|  | 1:226888854 | rs757273954 | G | T | Ala198Ser | 1.20E-05 | 0 | 1 | 0 |
|  | 1:226888867 | rs750266631 | C | T | Pro202Leu | 7.96E-06 | 1 | 0 | 0 |
|  | 1:226888877 |  | G | C | Leu205Phe |  | 3 | 1 | 0 |
|  | 1:226888917 |  | A | G | Ile219Val |  | 1 | 0 | 0 |
|  | 1:226888927 | rs1163620977 | A | C | Lys222Thr |  | 0 | 0 | 1 |
|  | 1:226888972 | rs200670135 | C | T | Ala237Val | 5.17E-05 | 1 | 0 | 0 |
|  | 1:226888974 | rs367855127 | C | T | Leu238Phe | 1.59E-05 | 0 | 1 | 0 |
|  | 1:226888975 | rs1211631545 | T | C | Leu238Pro | 1.19E-05 | 0 | 4 | 0 |
|  | 1:226888996 | rs1213166860 | A | G | Lys245Arg |  | 1 | 0 | 0 |
|  | 1:226889016 | rs138836272 | G | A | Ala252Thr | 0.0001915 | 11 | 1 | 2 |
|  | 1:226889034 | rs148238688 | G | A | Ala258Thr | 0.0001245 | 8 | 3 | 2 |
|  | 1:226890040 | rs199707432 | G | A | Val265Met | 7.95E-06 | 0 | 1 | 0 |
|  | 1:226890079 |  | G | T | Val278Leu |  | 0 | 1 | 0 |
|  | 1:226890097 | rs1208742830 | A | G | Arg284Gly | 3.98E-06 | 0 | 1 | 0 |
|  | 1:226890098 |  | G | A | Arg284Lys |  | 1 | 0 | 0 |
|  | 1:226890106 |  | C | T | Pro287Ser |  | 0 | 0 | 1 |
|  | 1:226890119 | rs374055800 | C | T | Ala291Val | 7.95E-06 | 0 | 1 | 0 |
|  | 1:226891283 | rs1254068585 | A | G | Met298Val | 3.99E-06 | 1 | 0 | 0 |
|  | 1:226891293 | rs144277432 | C | T | Thr301Met | 5.99E-05 | 2 | 0 | 0 |
|  | 1:226891298 |  | G | A | Gly303Ser |  | 0 | 1 | 0 |
|  | 1:226891319 | rs781436525 | T | C | Ser310Pro | 1.60E-05 | 0 | 1 | 0 |
|  | 1:226891331 |  | G | A | Ala314Thr |  | 0 | 1 | 0 |
|  | 1:226891347 | rs547494670 | A | G | Tyr319Cys | 3.22E-05 | 7 | 4 | 0 |
|  | 1:226891349 | rs565698726 | G | A | Asp320Asn | 1.21E-05 | 4 | 0 | 0 |
|  | 1:226891353 | rs1311206219 | C | T | Pro321Leu | 4.06E-06 | 1 | 0 | 0 |
|  | 1:226891754 | rs994711686 | T | C | Tyr328His | 7.95E-06 | 1 | 1 | 0 |
|  | 1:226891755 |  | A | G | Tyr328Cys |  | 0 | 1 | 0 |
|  | 1:226891772 | rs777639021 | C | G | Pro334Ala | 2.78E-05 | 2 | 1 | 0 |
|  | 1:226891773 | rs63750207 | C | G | Pro334Arg | 4.37E-05 | 3 | 3 | 0 |
|  | 1:226891805 |  | A | G | Thr345Ala |  | 1 | 0 | 0 |
|  | 1:226891809 | rs1365789341 | G | A | Gly346Asp | 3.98E-06 | 1 | 0 | 0 |
|  | 1:226891817 | rs759669954 | G | A | Gly349Arg | 1.19E-05 | 1 | 0 | 0 |
|  | 1:226891842 |  | A | G | Glu357Gly |  | 0 | 1 | 0 |
|  | 1:226894007 | rs940775081 | G | C | Arg358Thr |  | 0 | 1 | 0 |
|  | 1:226894027 | rs1449580232 | G | A | Gly365Arg | 3.98E-06 | 0 | 1 | 0 |
|  | 1:226894030 | rs1412191218 | G | A | Asp366Asn | 3.98E-06 | 1 | 0 | 0 |
|  | 1:226894064 | rs199714082 | C | T | Ala377Val | 4.38E-05 | 2 | 2 | 1 |
|  | 1:226894082 | rs145671982 | G | A | Gly383Glu | 3.98E-06 | 1 | 0 | 0 |
|  | 1:226894094 | rs867544084 | C | T | Thr387Ile | 7.96E-06 | 0 | 1 | 0 |
|  | 1:226894111 | rs142690225 | G | A | Val393Met | 0.0001075 | 2 | 6 | 3 |
|  | 1:226895442 | rs1250032273 | C | G | Leu404Val | 3.99E-06 | 0 | 1 | 0 |
|  | 1:226895490 | rs1558155801 | A | G | Ile420Val | 3.98E-06 | 0 | 0 | 1 |
|  | 1:226895526 | rs199676786 | A | C | Asn432His | 7.98E-06 | 1 | 0 | 0 |
|  | 1:226895535 | rs142772982 | C | T | Arg435Trp | 0.0001359 | 2 | 0 | 0 |
|  | 1:226895536 | rs201922151 | G | A | Arg435Gln | 3.60E-05 | 0 | 3 | 0 |
|  | 1:226895539 | rs749755334 | C | T | Pro436Leu | 1.20E-05 | 0 | 1 | 0 |
|  | 1:226895548 | rs63750110 | A | C | Asp439Ala | 3.61E-05 | 0 | 1 | 1 |
|  | 1:226895562 | rs151048692 | C | T | His444Tyr | 5.65E-05 | 5 | 4 | 0 |
| ***PSEN1*** | 14:73170945 | rs63749824 | C | T | Ala79Val | 1.19E-05 | 1 | 10 | 0 |
|  | 14:73170986 | rs146855665 | A | G | Met93Val | 3.98E-06 | 0 | 1 | 0 |
|  | 14:73170988 |  | G | T | Met93Ile |  | 0 | 2 | 0 |
|  | 14:73170989 | rs63750831 | G | A | Val94Met | 7.95E-06 | 1 | 0 | 0 |
|  | 14:73171007 | rs1335033402 | A | C | Ile100Leu |  | 0 | 1 | 0 |
|  | 14:73173570 |  | T | C | Tyr115His |  | 0 | 1 | 0 |
|  | 14:73173571 | rs63750450 | A | G | Tyr115Cys |  | 0 | 0 | 1 |
|  | 14:73173642 | rs63751037 | A | G | Met139Val |  | 0 | 2 | 0 |
|  | 14:73173665 | rs63750391 | G | C | Met146Ile |  | 0 | 1 | 0 |
|  | 14:73173669 | rs747363386 | A | G | Ile148Val | 7.95E-06 | 1 | 2 | 0 |
|  | 14:73186860 | rs63750590 | A | G | His163Arg |  | 0 | 1 | 0 |
|  | 14:73186868 | rs1555354564 | C | G | Leu166Val |  | 0 | 1 | 0 |
|  | 14:73186904 | rs63750155 | T | C | Ser178Pro |  | 0 | 1 | 0 |
|  | 14:73186910 |  | A | C | Ile180Leu |  | 0 | 1 | 0 |
|  | 14:73192661 | rs556147068 | A | G | Tyr189Cys | 1.19E-05 | 4 | 0 | 1 |
|  | 14:73192667 | rs112451138 | T | C | Val191Ala |  | 0 | 0 | 1 |
|  | 14:73192673 | rs780461228 | T | G | Val193Gly | 1.19E-05 | 0 | 1 | 0 |
|  | 14:73192679 | rs200065583 | A | G | Tyr195Cys |  | 0 | 1 | 0 |
|  | 14:73192681 | rs1162159737 | A | G | Ile196Val | 3.98E-06 | 0 | 1 | 0 |
|  | 14:73192687 | rs201375628 | G | C | Val198Leu | 3.98E-06 | 1 | 2 | 0 |
|  | 14:73192704 | rs1384308168 | G | C | Trp203Cys | 3.98E-06 | 1 | 0 | 0 |
|  | 14:73192712 | rs63750082 | G | C | Gly206Ala | 7.95E-06 | 1 | 21 | 4 |
|  | 14:73192717 | rs543391977 | G | T | Val208Leu | 7.95E-06 | 0 | 1 | 0 |
|  | 14:73192730 | rs1555355250 | C | A | Ser212Tyr |  | 0 | 1 | 0 |
|  | 14:73192735 | rs63751003 | C | T | His214Tyr | 3.98E-06 | 0 | 1 | 0 |
|  | 14:73192748 | rs140064975 | C | T | Pro218Leu | 7.95E-06 | 0 | 1 | 0 |
|  | 14:73192754 | rs763831389 | G | A | Arg220Gln | 1.59E-05 | 0 | 3 | 0 |
|  | 14:73192772 | rs63749961 | T | G | Leu226Arg |  | 0 | 1 | 0 |
|  | 14:73192774 | rs199842082 | A | G | Ile227Val | 1.19E-05 | 2 | 0 | 0 |
|  | 14:73192792 | CM004071 | A | C | Met233Leu |  | 0 | 1 | 0 |
|  | 14:73192798 | rs63751130 | C | G | Leu235Val |  | 0 | 0 | 1 |
|  | 14:73192828 | rs63750888 | A | C | Thr245Pro |  | 0 | 2 | 0 |
|  | 14:73192840 | rs1362575880 | A | C | Ile249Leu | 3.98E-06 | 0 | 2 | 0 |
|  | 14:73198042 | rs63750964 | G | T | Val261Phe |  | 0 | 1 | 0 |
|  | 14:73198052 | rs63750301 | C | T | Pro264Leu | 4.00E-06 | 0 | 1 | 1 |
|  | 14:73198067 | rs63750900 | G | A | Arg269His | 3.99E-06 | 0 | 4 | 0 |
|  | 14:73198078 |  | G | A | Glu273Lys |  | 0 | 1 | 0 |
|  | 14:73198123 |  | T | C | Tyr288His |  | 0 | 0 | 1 |
|  | 14:73206388 | rs63750298 | A | G | Thr291Ala | 3.98E-06 | 3 | 1 | 0 |
|  | 14:73206393 |  | G | C | Met292Ile |  | 1 | 0 | 0 |
|  | 14:73206421 |  | G | A | Asp302Asn |  | 0 | 1 | 0 |
|  | 14:73206421 |  | G | T | Asp302Tyr |  | 0 | 1 | 0 |
|  | 14:73206434 |  | A | C | Gln306Pro |  | 0 | 1 | 0 |
|  | 14:73206449 | rs115865530 | A | G | Lys311Arg | 0.0001392 | 3 | 1 | 0 |
|  | 14:73211811 | rs121917809 | A | G | Asp333Gly | 7.56E-05 | 5 | 7 | 0 |
|  | 14:73211816 | rs138871096 | G | A | Gly335Arg | 3.58E-05 | 2 | 1 | 0 |
|  | 14:73211847 | rs1018763711 | A | G | Asp345Gly | 7.95E-06 | 2 | 0 | 0 |
|  | 14:73211874 | rs63751164 | C | T | Thr354Ile | 7.95E-06 | 0 | 0 | 1 |
|  | 14:73211876 | rs376433615 | C | T | Pro355Ser | 3.18E-05 | 2 | 5 | 1 |
|  | 14:73211891 | rs199715992 | G | A | Ala360Thr | 3.18E-05 | 0 | 2 | 1 |
|  | 14:73211900 | rs996227958 | G | C | Glu363Gln | 1.19E-05 | 0 | 1 | 0 |
|  | 14:73211906 | rs200888596 | T | G | Ser365Ala | 1.59E-05 | 1 | 0 | 0 |
|  | 14:73211907 | rs63750941 | C | A | Ser365Tyr |  | 1 | 0 | 0 |
|  | 14:73217129 | rs63750323 | G | T | Gly378Val |  | 0 | 1 | 0 |
|  | 14:73217162 |  | A | G | Tyr389Cys |  | 0 | 1 | 0 |
|  | 14:73217164 |  | A | C | Ser390Arg |  | 1 | 1 | 0 |
|  | 14:73217165 |  | G | A | Ser390Asn |  | 0 | 1 | 0 |
|  | 14:73217182 |  | G | A | Ala396Thr |  | 0 | 1 | 0 |
|  | 14:73217221 | rs63750227 | G | A | Ala409Thr |  | 1 | 0 | 0 |
|  | 14:73217225 | rs661 | G | A | Cys410Tyr |  | 0 | 2 | 0 |
|  | 14:73219161 | rs63751223 | G | C | Ala426Pro |  | 0 | 1 | 0 |
|  | 14:73219164 | rs1398951357 | A | G | Ile427Val | 7.95E-06 | 0 | 1 | 0 |
|  | 14:73219177 | rs63750083 | C | A | Ala431Glu |  | 0 | 1 | 2 |
|  | 14:73219194 | rs764971634 | A | G | Ile437Val |  | 0 | 1 | 0 |
|  | 14:73219248 | rs140169796 | C | T | Pro455Ser | 3.98E-06 | 1 | 0 | 0 |
|  | 14:73219254 | rs1430581353 | A | G | Met457Val |  | 0 | 1 | 0 |
|  | 14:73219266 | rs1186921147 | G | T | Ala461Ser | 7.96E-06 | 0 | 2 | 0 |
|  | 14:73219273 | rs780119074 | A | G | His463Arg | 1.59E-05 | 3 | 0 | 0 |
| ***APP*** | 21:25881698 | rs373482247 | T | C | Tyr762Cys | 3.98E-06 | 0 | 1 | 0 |
|  | 21:25881730 | rs1381242897 | C | G | Lys751Asn | 3.98E-06 | 1 | 0 | 0 |
|  | 21:25881744 | rs1244541473 | G | A | Arg747Cys | 3.98E-06 | 1 | 0 | 0 |
|  | 21:25881770 | rs1353342006 | A | T | Val738Asp |  | 1 | 0 | 0 |
|  | 21:25881771 | rs756759593 | C | T | Val738Ile | 7.97E-06 | 1 | 0 | 0 |
|  | 21:25891783 | rs63749964 | A | C | Val717Gly |  | 0 | 1 | 0 |
|  | 21:25891784 | rs63750264 | C | A | Val717Phe |  | 0 | 1 | 0 |
|  | 21:25891787 | rs63750399 | T | C | Ile716Val |  | 0 | 1 | 1 |
|  | 21:25891796 | rs63750066 | C | T | Ala713Thr | 9.55E-05 | 0 | 1 | 0 |
|  | 21:25891808 | rs201269325 | C | T | Gly709Ser | 3.98E-05 | 1 | 0 | 0 |
|  | 21:25891854 | rs768446605 | T | A | Glu693Asp | 3.98E-06 | 0 | 1 | 0 |
|  | 21:25897586 | rs1424019513 | T | G | His684Pro | 3.98E-06 | 0 | 1 | 1 |
|  | 21:25897607 | rs63749953 | T | C | His677Arg | 3.98E-06 | 0 | 1 | 0 |
|  | 21:25897610 | rs559207950 | C | T | Arg676Gln | 2.39E-05 | 1 | 0 | 0 |
|  | 21:25897620 | rs63750847 | C | T | Ala673Thr | 0.0004455 | 5 | 1 | 0 |
|  | 21:25897626 | rs572842823 | T | A | Met671Leu |  | 0 | 1 | 0 |
|  | 21:25897626 | rs572842823 | T | G | Met671Leu |  | 0 | 1 | 0 |
|  | 21:25897627 | rs371425292 | C | A | Lys670Asn |  | 0 | 1 | 0 |
|  | 21:25897632 | rs1259157720 | C | T | Val669Met | 3.98E-06 | 1 | 0 | 0 |
|  | 21:25897641 | rs200084346 | T | A | Ile666Phe | 2.39E-05 | 0 | 1 | 0 |
|  | 21:25897642 | rs63750363 | C | G | Glu665Asp | 1.19E-05 | 2 | 3 | 1 |
|  | 21:25897644 | rs201945946 | C | T | Glu665Lys | 0.0002506 | 1 | 0 | 0 |
|  | 21:25897649 | rs200260102 | G | A | Thr663Met | 1.59E-05 | 1 | 0 | 0 |
|  | 21:25897658 |  | T | G | Asn660Thr |  | 0 | 0 | 1 |
|  | 21:25897659 |  | T | A | Asn660Tyr |  | 0 | 1 | 0 |
|  | 21:25897668 | rs1305531440 | C | T | Gly657Arg |  | 0 | 1 | 0 |
|  | 21:25905029 |  | C | T | Arg653Gln |  | 1 | 0 | 0 |
|  | 21:25905044 | rs765412723 | C | T | Arg648Gln | 1.20E-05 | 2 | 0 | 0 |
|  | 21:25905048 | rs368159818 | C | T | Asp647Asn | 7.98E-06 | 2 | 0 | 1 |
|  | 21:25911778 | rs780774625 | A | C | Phe624Leu | 3.98E-06 | 1 | 0 | 0 |
|  | 21:25911791 | rs759926843 | G | A | Pro620Leu | 1.99E-05 | 3 | 0 | 0 |
|  | 21:25911801 | rs201479601 | C | T | Asp617Asn | 7.95E-06 | 2 | 0 | 0 |
|  | 21:25911804 | rs776739799 | C | T | Asp616Asn | 2.39E-05 | 0 | 2 | 0 |
|  | 21:25911810 | rs112263157 | T | C | Ser614Gly | 0.001097 | 120 | 88 | 12 |
|  | 21:25911840 | rs199887707 | C | T | Val604Met | 0.0003778 | 4 | 1 | 1 |
|  | 21:25911851 | rs200088099 | G | A | Thr600Met | 0.0002028 | 1 | 1 | 1 |
|  | 21:25911855 | rs140304729 | C | T | Glu599Lys | 0.001372 | 15 | 21 | 2 |
|  | 21:25911860 | rs765301301 | A | C | Leu597Trp | 1.19E-05 | 0 | 3 | 0 |
|  | 21:25911861 | rs773379255 | A | C | Leu597Val | 3.98E-06 | 1 | 0 | 0 |
|  | 21:25911899 | rs145371658 | G | C | Pro584Arg | 3.18E-05 | 4 | 1 | 1 |
|  | 21:25911906 | rs1249646636 | T | C | Ser582Gly |  | 0 | 2 | 0 |
|  | 21:25911907 |  | A | C | Ile581Met |  | 1 | 0 | 0 |
|  | 21:25911924 | rs200769792 | C | T | Val576Ile | 5.57E-05 | 2 | 1 | 0 |
|  | 21:26000072 | rs754207226 | C | T | Gly326Ser | 2.39E-05 | 1 | 1 | 0 |
|  | 21:26021846 |  | C | A | Val287Phe |  | 0 | 0 | 0 |
|  | 21:26021879 | rs202198008 | T | A | Thr276Ser | 0.0004186 | 24 | 25 | 5 |
|  | 21:26021921 | rs779428437 | A | G | Tyr262His | 1.23E-05 | 0 | 0 | 1 |
|  | 21:26022001 | rs139819006 | G | A | Ala235Val | 0.0001368 | 3 | 1 | 0 |
|  | 21:26051045 | rs376138420 | G | A | Ser206Leu | 7.95E-06 | 1 | 0 | 0 |
|  | 21:26051070 | rs145081708 | A | G | Ser198Pro | 0.0006841 | 23 | 22 | 3 |
|  | 21:26051100 | rs199744129 | G | A | Pro188Ser | 3.98E-06 | 2 | 0 | 0 |
|  | 21:26053291 | rs1451050785 | T | C | Gln138Arg |  | 1 | 0 | 0 |
|  | 21:26053303 | rs576712021 | T | C | Lys134Arg | 1.19E-05 | 1 | 1 | 0 |
|  | 21:26053319 | rs563647959 | C | T | Val129Ile | 3.98E-06 | 0 | 0 | 1 |
|  | 21:26089993 | rs777260127 | C | T | Arg102His | 1.59E-05 | 1 | 0 | 0 |
|  | 21:26089999 | rs199741007 | C | T | Arg100Gln | 3.98E-06 | 1 | 0 | 0 |
|  | 21:26090000 | rs200347552 | G | A | Arg100Trp | 1.59E-05 | 2 | 0 | 0 |
|  | 21:26090015 | rs950592627 | G | C | Gln95Glu |  | 2 | 0 | 0 |
|  | 21:26090047 | rs777179682 | T | C | Asn84Ser | 7.95E-06 | 0 | 2 | 0 |
|  | 21:26090056 |  | T | C | Gln81Arg |  | 1 | 0 | 0 |
|  | 21:26090072 | rs151188448 | C | T | Val76Ile | 9.55E-05 | 6 | 3 | 0 |
|  | 21:26111992 | rs757264249 | T | C | Gln71Arg | 3.98E-06 | 0 | 1 | 0 |
|  | 21:26112006 | rs1411114284 | C | G | Lys66Asn | 3.98E-06 | 1 | 0 | 0 |
|  | 21:26112016 | rs200648330 | A | G | Ile63Thr | 3.98E-06 | 0 | 0 | 1 |
|  | 21:26112074 | rs774281569 | G | A | His44Tyr | 7.95E-06 | 0 | 0 | 1 |
|  | 21:26112143 | rs1482948530 | G | A | Pro21Ser | 3.98E-06 | 0 | 1 | 0 |
|  | 21:26170574 | rs955517095 | C | T | Arg16Gln | 2.96E-05 | 2 | 1 | 0 |
|  | 21:26170584 | rs1206506558 | A | T | Trp13Arg | 7.38E-06 | 0 | 2 | 0 |
|  | 21:26170589 |  | G | C | Ala11Gly |  | 1 | 1 | 0 |
|  | 21:26170602 | rs767211549 | G | C | Leu7Val | 1.49E-05 | 0 | 0 | 1 |
|  | 21:26170608 | rs1446208112 | A | C | Leu5Val | 1.50E-05 | 0 | 1 | 0 |

*Allele frequency from VEP annotation using the gnomADe_AF.

**Variant positions were based on Genome Reference Consortium Human Build 38 (GRCh 38).

**Supplementary Table e-6: AD Diagnosis Distribution and Alternative Allele Clinical Variant Carriers by NIAGADS Defined Study Sites.**

|  | AD Diagnosis | | |  |  |
| --- | --- | --- | --- | --- | --- |
| NIAGADS Defined Study Site | **Control** | **Case** | **Unknown** | **Total** | **% Clinical Variant Carriers** |
| Whole Exome Sequencing (WES) |  |  |  |  |  |
| ADSP | 4,115 | 5,379 | 341 | 9,835 | 0.2% |
| ADGC | 537 | 516 | 209 | 1,262 | 0% |
| FASe | 265 | 635 | 134 | 1,034 | 0% |
| Brkanac families | 0 | 26 | 16 | 42 | 7.1% |
| Miami families | 16 | 40 | 3 | 59 | 0% |
| WHICAP | 2,657 | 527 | 47 | 3,231 | 0.1% |
| Knight ADRC | 273 | 183 | 46 | 502 | 0.2% |
| CBD | 0 | 0 | 335 | 335 | 0% |
| PSP | 0 | 0 | 549 | 549 | 0% |
| Whole Genome Sequencing (WGS) |  |  |  |  |  |
| ADNI | 221 | 227 | 313 | 761 | 0% |
| ADSP Discovery Family | 74 | 456 | 31 | 561 | 0.4% |
| ADSP Extension Family | 338 | 68 | 17 | 423 | 0.2% |
| ADSP Extension Case-Control | 1,410 | 1,411 | 115 | 2,936 | 0.1% |
| ADSP Follow-up ADC | 1,227 | 1,398 | 98 | 2,723 | 0.1% |
| ADSP Follow-up ADGC AA | 1,190 | 686 | 31 | 1,907 | 0.1% |
| ADSP Follow-up ADNI | 190 | 346 | 194 | 730 | 0.1% |
| ADSP Follow-up APOE extreme | 259 | 370 | 231 | 860 | 3.8% |
| ADSP Follow-up HIHGBB | 2 | 78 | 5 | 85 | 0% |
| ADSP Follow-up PR1066 | 1,201 | 138 | 178 | 1,517 | 0.2% |
| ADSP Follow-up SteEPAD | 46 | 158 | 4 | 208 | 3.4% |
| AMPAD Mayo | 38 | 88 | 38 | 164 | 0% |
| AMPAD MSSM | 92 | 247 | 0 | 339 | 0.3% |
| AMPAD ROSMA | 336 | 391 | 0 | 727 | 0% |
| Cache County | 207 | 0 | 0 | 207 | 0.5% |
| FASe families | 9 | 73 | 4 | 86 | 0% |
| Knight ADRC | 10 | 53 | 7 | 70 | 0% |
| NACC Genetech | 2 | 122 | 2 | 126 | 2.4% |
| UPITT Kamboh | 0 | 209 | 0 | 209 | 0% |
| PSP | 0 | 0 | 2 | 2 | 0% |
| Total | 14,715 | 13,825 | 2,950 | 31,490 | 0.3% |

**Supplementary Table e-7: Percentage of Alternative Allele Carriers in Clinical Variants by Race and Ethnicity and AD Diagnosis.**

|  |  | AD Diagnosis | | |  |
| --- | --- | --- | --- | --- | --- |
| Ethnicity/Race | **Control (n=3)** | **Case**  **(n=62)** | **Unknown**  **(n=13)** | **Total Sample Size for each ethnicity/race group (%)** | **Clinical Variant Carrier % in Total** |
| Non-Hispanic |  |  |  |  |  |
| White | 2  (0.01%) | 38  (0.2%) | 6  (0.03%) | 21,083 (66.9%) | 0.2% |
| Black | 0  (0%) | 3  (0.1%) | 0  (0%) | 5,286  (16.9%) | 0.01% |
| Other/Unknown | 0  (0%) | 0  (0%) | 0  (0%) | 102  (0.3%) | 0% |
| Hispanic |  |  |  |  |  |
| White | 0  (0%) | 9  (0.3%) | 3  (0.9%) | 322  (1.0%) | 0.04% |
| Black | 0  (0%) | 0  (0%) | 1  (1.4%) | 74  (0.2%) | 0.003% |
| Other/Unknown | 1  (0.02%) | 12  (0.3%) | 3  (0.1%) | 4623  (14.7%) | 0.05% |

Data shown as frequency and percentage in each ethnicity/race group for AD controls, cases, and unknowns. Column percentage is shown for total sample size for each ethnicity/race group. Clinical carrier percentage in total sample is calculated as sum of carrier numbers in each ethnicity/race group divided by total sample size(n=31,490).

**Supplemental Table e-8 Adjusted Mean Age of Onset of Clinical Variant Carriers between Populations.**

|  | **Adjusted Mean** | **Beta Coefficient** | **Standard Error** | **p-value** |
| --- | --- | --- | --- | --- |
| **Race** | | | | |
| White (n=45) | 57.0 | N/A | N/A | N/A |
| African American (n=3) | 53.6 | -3.40 | 11.88 | 0.77 |
| Unknown/Other (n=12) | 64.5 | 7.49 | 3.72 | 0.04 |
| **Ethnicity** | | | | |
| Hispanic (n=21) | 67.1 | 13.04 | 4.36 | 0.003 |
| Non-Hispanic (n=39) | 54.1 | N/A | N/A | N/A |

Linear mixed effect model adjusted for sex, PC1-4, *APOE ε2, APOE ε4* and WGS/WES indicator.

**Supplementary Table e-9: APOE Status by AD Diagnosis among Clinical Variant Carriers and Noncarriers.**

|  | Clinical Variant Carriers | | | | | Noncarriers | | | |
| --- | --- | --- | --- | --- | --- | --- | --- | --- | --- |
|  | | **Control (N=3)** | **Case (N=62)** | **Unknown**  **(N=13)** | **Total (N=78)** | **Control (N=14,031)** | **Case (N=13,169)** | **Unknown**  **(N=2,807)** | **Total (N=30,007)** |
| APOE genotype | |  |  |  |  |  |  |  |  |
| ε2/ε2 | | 0 (0%) | 0 (0%) | 0 (0%) | 0 (0%) | 96 (0.7%) | 38 (0.3%) | 10 (0.4%) | 144 (0.5%) |
| ε2/ε3 | | 0 (0%) | 5 (8.1%) | 0 (0%) | 5 (6.4%) | 1,819 (13.0%) | 714 (5.4%) | 141 (5.0%) | 2,674 (8.9%) |
| ε2/ε4 | | 0 (0%) | 1 (1.6%) | 0 (0%) | 1 (1.3%) | 304 (2.2%) | 314 (2.4%) | 35 (1.2%) | 653 (2.2%) |
| ε3/ε3 | | 2 (66.7%) | 37 (59.7%) | 8 (61.5%) | 47 (60.3%) | 8,312 (59.2%) | 5,779 (43.9%) | 1,008 (35.9%) | 15,099 (50.4%) |
| ε3/ε4 | | 1 (33.3%) | 18 (29.0%) | 2 (15.4%) | 21 (26.9%) | 3,159 (22.5%) | 5,408 (41.1%) | 579 (20.7%) | 9,146 (30.5%) |
| ε4/ε4 | | 0 (0%) | 1 (1.6%) | 0 (0%) | 1 (1.3%) | 244 (1.7%) | 894 (6.8%) | 112 (4.0%) | 1,250 (4.1%) |
| NA | | 0 (0%) | 0 (0%) | 3 (23.1%) | 3 (3.8%) | 97 (0.7%) | 22 (0.2%) | 922 (32.8%) | 1,041 (3.5%) |
| APOE ε2 allele | |  |  |  |  |  |  |  |  |
| 0 | | 3 (100%) | 56 (90.3%) | 10 (76.9%) | 69 (88.5%) | 11,715 (83.5%) | 12,081 (91.7%) | 1,699 (60.5%) | 25,495 (85.0%) |
| 1 | | 0 (0%) | 6 (9.7%) | 0 (0%) | 6 (7.7%) | 2,123 (15.1%) | 1,038 (7.8%) | 176 (6.3%) | 3,327 (11.1%) |
| 2 | | 0 (0%) | 0 (0%) | 0 (0%) | 0 (0%) | 96 (0.7%) | 38 (0.3%) | 10 (0.4%) | 144 (0.5%) |
| NA | | 0 (0%) | 0 (0%) | 3 (23.1%) | 3 (3.8%) | 97 (0.7%) | 23 (0.2%) | 922 (32.8%) | 1,041 (3.5%) |
| APOE ε4 allele | |  |  |  |  |  |  |  |  |
| 0 | | 2 (66.7%) | 42 (67.7%) | 8 (61.5%) | 52 (66.7%) | 10,227 (72.9%) | 6,531 (49.6%) | 1,159 (41.3%) | 17,917 (59.7%) |
| 1 | | 1 (33.3%) | 19 (30.6%) | 2 (15.4%) | 22 (28.2%) | 3,463 (24.7%) | 5,722 (43.5%) | 614 (21.9%) | 9,799 (32.7%) |
| 2 | | 0 (0%) | 1 (1.6%) | 0 (0%) | 1 (1.3%) | 244 (1.8%) | 894 (6.8%) | 112 (4.0%) | 1,250 (4.1%) |
| NA | | 0 (0%) | 0 (0%) | 3 (23.1%) | 3 (3.8%) | 97 (0.7%) | 22 (0.2%) | 922 (32.8%) | 1,041 (3.5%) |

**Supplemental Table e-10. Clinical Variants by AD status Among Race/Ethnicity Groups.**

| **Gene** | **Chr:Position** | **REF** | **ALT** | **Non-Hispanic White** | **Non-Hispanic Black** | **Hispanic White** | **Hispanic Black** | **Hispanic Other/Unknow** |
| --- | --- | --- | --- | --- | --- | --- | --- | --- |
| ***PSEN2*** | 1:226883817 | C | T | 1/0/0 | 0/0/0 | 0/0/0 | 0/0/0 | 0/0/0 |
|  | 1:226885603 | A | T | 0/2/0 | 0/0/0 | 0/0/0 | 0/0/0 | 0/0/0 |
|  | 1:226890097 | A | G | 0/1/0 | 0/0/0 | 0/0/0 | 0/0/0 | 0/0/0 |
|  | 1:226895548 | A | C | 0/1/1 | 0/0/0 | 0/0/0 | 0/0/0 | 0/0/0 |
| ***PSEN1*** | 14:73170945 | C | T | 1/10/0 | 0/0/0 | 0/0/0 | 0/0/0 | 0/0/0 |
|  | 14:73173570 | T | C | 0/1/0 | 0/0/0 | 0/0/0 | 0/0/0 | 0/0/0 |
|  | 14:73173571 | A | G | 0/0/1 | 0/0/0 | 0/0/0 | 0/0/0 | 0/0/0 |
|  | 14:73173642 | A | G | 0/2/0 | 0/0/0 | 0/0/0 | 0/0/0 | 0/0/0 |
|  | 14:73173665 | G | C | 0/1/0 | 0/0/0 | 0/0/0 | 0/0/0 | 0/0/0 |
|  | 14:73186860 | A | G | 0/1/0 | 0/0/0 | 0/0/0 | 0/0/0 | 0/0/0 |
|  | 14:73186868 | C | G | 0/1/0 | 0/0/0 | 0/0/0 | 0/0/0 | 0/0/0 |
|  | 14:73186904 | T | C | 0/1/0 | 0/0/0 | 0/0/0 | 0/0/0 | 0/0/0 |
|  | 14:73192712 | G | C | 0/1/0 | 0/0/0 | 0/8/1 | 0/0/1 | 1/12/2 |
|  | 14:73192730 | C | A | 0/1/0 | 0/0/0 | 0/0/0 | 0/0/0 | 0/0/0 |
|  | 14:73192735 | C | T | 0/1/0 | 0/0/0 | 0/0/0 | 0/0/0 | 0/0/0 |
|  | 14:73192772 | T | G | 0/1/0 | 0/0/0 | 0/0/0 | 0/0/0 | 0/0/0 |
|  | 14:73192792 | A | C | 0/1/0 | 0/0/0 | 0/0/0 | 0/0/0 | 0/0/0 |
|  | 14:73192798 | C | G | 0/0/0 | 0/0/0 | 0/0/1 | 0/0/0 | 0/0/0 |
|  | 14:73192840 | A | C | 0/1/0 | 0/1/0 | 0/0/0 | 0/0/0 | 0/0/0 |
|  | 14:73198042 | G | T | 0/1/0 | 0/0/0 | 0/0/0 | 0/0/0 | 0/0/0 |
|  | 14:73198052 | C | T | 0/1/1 | 0/0/0 | 0/0/0 | 0/0/0 | 0/0/0 |
|  | 14:73198067 | G | A | 0/2/0 | 0/2/0 | 0/0/0 | 0/0/0 | 0/0/0 |
|  | 14:73206385 | G | T | 0/1/0 | 0/0/0 | 0/0/0 | 0/0/0 | 0/0/0 |
|  | 14:73206385 | G | A | 0/0/3 | 0/0/0 | 0/0/0 | 0/0/0 | 0/0/0 |
|  | 14:73217129 | G | T | 0/1/0 | 0/0/0 | 0/0/0 | 0/0/0 | 0/0/0 |
|  | 14:73217182 | G | A | 0/1/0 | 0/0/0 | 0/0/0 | 0/0/0 | 0/0/0 |
|  | 14:73217225 | G | A | 0/2/0 | 0/0/0 | 0/0/0 | 0/0/0 | 0/0/0 |
|  | 14:73219161 | G | C | 0/1/0 | 0/0/0 | 0/0/0 | 0/0/0 | 0/0/0 |
|  | 14:73219177 | C | A | 0/0/0 | 0/0/0 | 0/1/1 | 0/0/0 | 0/0/1 |
| ***APP*** | 21:25891783 | A | C | 0/1/0 | 0/0/0 | 0/0/0 | 0/0/0 | 0/0/0 |
|  | 21:25891784 | C | A | 0/1/0 | 0/0/0 | 0/0/0 | 0/0/0 | 0/0/0 |

Variants shown as count in AD control/case/unknown individuals.

**Supplementary Table e-11: Carrier Status, Demographics, and Study among Individuals with Unknown AD Status.**

|  | **Clinical Variant Carriers** | **Conflicting Clinical Variant Carriers** | **Additional DM Variant Carriers** | **Total** |
| --- | --- | --- | --- | --- |
|  | **(N=13)** | **(N=56)** | **(N=70)** | **(N=2,950)** |
| **Sex** |  |  |  |  |
| Female | 7 (53.9%) | 23 (41.07%) | 30 (42.86%) | 1439 (48.78%) |
| Male | 6 (46.2%) | 33 (58.93%) | 40 (57.14%) | 1511 (51.22%) |
| **Ethnicity/Race** |  |  |  |  |
| Non-Hispanic |  |  |  |  |
| White | 6 (46.2%) | 44 (78.57%) | 39 (55.71%) | 2334 (79.12%) |
| Black | 0 (0%) | 8 (14.29%) | 16 (22.86%) | 307 (10.41%) |
| Unknown/Other | 0 (0%) | 0 (0%) | 1 (1.43%) | 23 (0.78%) |
| Hispanic |  |  |  |  |
| Black | 1 (7.7%) | 0 (0%) | 0 (0%) | 2 (0.07%) |
| Other/Unknown | 3 (23.1%) | 4 (7.14%) | 12 (17.14%) | 248 (8.41%) |
| White | 3 (23.1%) | 0 (0%) | 2 (2.86%) | 36 (1.22%) |
| **NIAGADS Defined Study Site** |  |  |  |  |
| **Whole Genome Sequencing (WGS)** | |  |  |  |
| ADNI | 0 (0%) | 7 (12.5%) | 10 (14.3%) | 313 (10.6%) |
| ADSP Discovery Family | 0 (0%) | 1 (1.8%) | 1 (1.4%) | 31 (1.1%) |
| ADSP Extension Family | 0 (0%) | 0 (0%) | 0 (0%) | 17 (0.6%) |
| ADSP Extension Case-Control | 0 (0%) | 1 (1.8%) | 5 (7.1%) | 115 (3.9%) |
| ADSP Follow-up ADC | 1 (7.7%) | 5 (8.9%) | 0 (0%) | 98 (3.3%) |
| ADSP Follow-up ADGC AA | 0 (0%) | 1 (1.8%) | 1 (1.4%) | 31 (1.1%) |
| ADSP Follow-up ADNI | 0 (0%) | 3 (5.4%) | 8 (11.4%) | 194 (6.6%) |
| ADSP Follow-up APOE extreme | 7 (53.9%) | 4 (7.1%) | 6 (8.6%) | 231 (7.8%) |
| ADSP Follow-up HIHGBB | 0 (0%) | 0 (0%) | 0 (0%) | 5 (0.2%) |
| ADSP Follow-up PR1066 | 1 (7.7%) | 4 (7.1%) | 5 (7.1%) | 178 (6.03%) |
| ADSP Follow-up SteEPAD | 0 (0%) | 0 (0%) | 0 (0%) | 4 (0.1%) |
| AMPAD Mayo | 0 (0%) | 1 (1.8%) | 0 (0%) | 38 (1.3%) |
| FASe families | 0 (0%) | 0 (0%) | 0 (0%) | 4 (0.1%) |
| Knight ADRC | 0 (0%) | 0 (0%) | 0 (0%) | 7 (0.2%) |
| NACC Genetech | 0 (0%) | 0 (0%) | 0 (0%) | 2 (0.1%) |
| PSP | 0 (0%) | 0 (0%) | 1 (1.4%) | 2 (0.1%) |
| **Whole Exome Sequencing (WES)** | |  |  |  |
| ADSP | 0 (0%) | 5 (8.9%) | 6 (8.6%) | 341 (11.6%) |
| ADGC | 0 (0%) | 7 (12.5%) | 6 (8.6%) | 209 (7.1%) |
| FASe | 0 (0%) | 1 (1.8%) | 1 (1.4%) | 134 (4.5%) |
| Brkanac families | 3 (23.1%) | 0 (0%) | 0 (0%) | 16 (0.5%) |
| Miami families | 0 (0%) | 0 (0%) | 0 (0%) | 3 (0.1%) |
| WHICAP | 1 (7.7%) | 0 (0%) | 5 (7.1%) | 47 (1.6%) |
| Knight ADRC | 0 (0%) | 0 (0%) | 1 (1.4%) | 46 (1.6%) |
| CBD | 0 (0%) | 6 (10.7%) | 3 (4.3%) | 335 (11.4%) |
| PSP | 0 (0%) | 10 (17.9%) | 11 (15.7%) | 549 (18.6%) |

**Supplementary Table e-12. Original AD Status by Cohort among the AD Unknown Participants.**

|  | **ADNI (N=313)** | **Case-control (N=779)** | **Family (N=202)** | **Missing in Phenotype File (N=775)** | **PSP/CBD (N=881)** | **Overall (N=2,950)** |
| --- | --- | --- | --- | --- | --- | --- |
| **Base_MCI** |  |  |  |  |  |  |
| Yes | 313 (100%) | 0 (0%) | 0 (0%) | 0 (0%) | 0 (0%) | 313 (10.61%) |
| Missing | 0 (0%) | 779 (100%) | 202 (100%) | 775 (100%) | 881 (100%) | 2,637 (89.39%) |
| **Base_AD  (Family)** |  |  |  |  |  |  |
| No dementia | 0 (0%) | 0 (0%) | 2 (1.0%) | 0 (0%) | 0 (0%) | 2 (0.07%) |
| Definite AD | 0 (0%) | 0 (0%) | 1 (0.5%) | 0 (0%) | 0 (0%) | 1 (0.03%) |
| Probable AD | 0 (0%) | 0 (0%) | 7 (3.5%) | 0 (0%) | 0 (0%) | 7 (0.2%) |
| Possible AD | 0 (0%) | 0 (0%) | 18 (8.9%) | 0 (0%) | 0 (0%) | 18 (0.6%) |
| Family Reported AD | 0 (0%) | 0 (0%) | 53 (26.2%) | 0 (0%) | 0 (0%) | 53 (1.8%) |
| Other Dementia | 0 (0%) | 0 (0%) | 50 (24.8%) | 0 (0%) | 0 (0%) | 51 (1.7%) |
| Unknown | 0 (0%) | 0 (0%) | 62 (30.7%) | 0 (0%) | 0 (0%) | 64 (2.2%) |
| Family Reported No AD | 0 (0%) | 0 (0%) | 9 (4.5%) | 0 (0%) | 0 (0%) | 9 (0.3%) |
| Missing | 313 (100%) | 779 (100%) | 0(0%) | 775 (100%) | 881 (100%) | 2,745 (93.1%) |
| **Base_AD**  **(Case-control)** |  |  |  |  |  |  |
| No | 0 (0%) | 339 (43.5%) | 0 (0%) | 0 (0%) | 0 (0%) | 339 (11.5%) |
| Yes | 0 (0%) | 120 (15.4%) | 0 (0%) | 0 (0%) | 0 (0%) | 120 (4.07%) |
| Missing | 313 (100%) | 320 (41.1%) | 202 (100%) | 775 (100%) | 881 (100%) | 2,491 (84.4%) |
| **Diagnosis** |  |  |  |  |  |  |
| PSP | 0 (0%) | 0 (0%) | 0 (0%) | 0 (0%) | 549 (62.3%) | 549 (18.6%) |
| CBD | 0 (0%) | 0 (0%) | 0 (0%) | 0 (0%) | 332 (37.7%) | 332 (11.3%) |
| Missing | 313 (100%) | 779 (100%) | 202 (100%) | 775 (100%) | 0 (0%) | 2,069 (70.1%) |
| **Base_PrevAD** |  |  |  |  |  |  |
| No | 313 (100%) | 387 (49.7%) | 0 (0%) | 0 (0%) | 0 (0%) | 700 (23.7%) |
| Yes | 0 (0%) | 72 (9.2%) | 0 (0%) | 0 (0%) | 0 (0%) | 72 (2.4%) |
| Missing | 0 (0%) | 320 (41.1%) | 202 (100%) | 775 (100%) | 881 (100%) | 2,178 (73.8%) |
| **Base_IncAD** |  |  |  |  |  |  |
| No | 313 (100%) | 408 (52.4%) | 0 (0%) | 0 (0%) | 0 (0%) | 721 (24.4%) |
| Yes | 0 (0%) | 51 (6.6%) | 0 (0%) | 0 (0%) | 0 (0%) | 51 (1.7%) |
| Missing | 0 (0%) | 320 (41.1%) | 202 (100%) | 775 (100%) | 881 (100%) | 2,178 (73.8%) |
| **PrevAD** |  |  |  |  |  |  |
| No | 313 (100%) | 0 (0%) | 0 (0%) | 0 (0%) | 0 (0%) | 313 (10.6%) |
| Missing | 0 (0%) | 779 (100%) | 0 (0%) | 0 (0%) | 0 (0%) | 2,637 (89.4%) |
| **IncAD** |  |  |  |  |  |  |
| No | 313 (100%) | 0 (0%) | 0 (0%) | 0 (0%) | 0 (0%) | 313 (10.6%) |
| Missing | 0 (0%) | 779 (100%) | 202 (100%) | 775 (100%) | 881 (100%) | 2,637 (89.4%) |

**Acknowledgments:**

### sa000001 - Alzheimer’s Disease Sequencing Project (ADSP)

The Alzheimer’s Disease Sequencing Project (ADSP) is comprised of two Alzheimer’s Disease (AD) genetics consortia and three National Human Genome Research Institute (NHGRI) funded Large Scale Sequencing and Analysis Centers (LSAC). The two AD genetics consortia are the Alzheimer’s Disease Genetics Consortium (ADGC) funded by NIA (U01 AG032984), and the Cohorts for Heart and Aging Research in Genomic Epidemiology (CHARGE) funded by NIA (R01 AG033193), the National Heart, Lung, and Blood Institute (NHLBI), other National Institute of Health (NIH) institutes and other foreign governmental and non-governmental organizations. The Discovery Phase analysis of sequence data is supported through UF1AG047133 (to Drs. Schellenberg, Farrer, Pericak-Vance, Mayeux, and Haines); U01AG049505 to Dr. Seshadri; U01AG049506 to Dr. Boerwinkle; U01AG049507 to Dr. Wijsman; and U01AG049508 to Dr. Goate and the Discovery Extension Phase analysis is supported through U01AG052411 to Dr. Goate, U01AG052410 to Dr. Pericak-Vance and U01 AG052409 to Drs. Seshadri and Fornage.

Sequencing for the Follow Up Study (FUS) is supported through U01AG057659 (to Drs. PericakVance, Mayeux, and Vardarajan) and U01AG062943 (to Drs. Pericak-Vance and Mayeux). Data generation and harmonization in the Follow-up Phase is supported by U54AG052427 (to Drs. Schellenberg and Wang). The FUS Phase analysis of sequence data is supported through U01AG058589 (to Drs. Destefano, Boerwinkle, De Jager, Fornage, Seshadri, and Wijsman), U01AG058654 (to Drs. Haines, Bush, Farrer, Martin, and Pericak-Vance), U01AG058635 (to Dr. Goate), RF1AG058066 (to Drs. Haines, Pericak-Vance, and Scott), RF1AG057519 (to Drs. Farrer and Jun), R01AG048927 (to Dr. Farrer), and RF1AG054074 (to Drs. Pericak-Vance and Beecham).

The ADGC cohorts include: Adult Changes in Thought (ACT) (U01 AG006781, U19 AG066567), the Alzheimer’s Disease Research Centers (ADRC) (P30 AG062429, P30 AG066468, P30 AG062421, P30 AG066509, P30 AG066514, P30 AG066530, P30 AG066507, P30 AG066444, P30 AG066518, P30 AG066512, P30 AG066462, P30 AG072979, P30 AG072972, P30 AG072976, P30 AG072975, P30 AG072978, P30 AG072977, P30 AG066519, P30 AG062677, P30 AG079280, P30 AG062422, P30 AG066511, P30 AG072946, P30 AG062715, P30 AG072973, P30 AG066506, P30 AG066508, P30 AG066515, P30 AG072947, P30 AG072931, P30 AG066546, P20 AG068024, P20 AG068053, P20 AG068077, P20 AG068082, P30 AG072958, P30 AG072959), the Chicago Health and Aging Project (CHAP) (R01 AG11101, RC4 AG039085, K23 AG030944), Indiana Memory and Aging Study (IMAS) (R01 AG019771), Indianapolis Ibadan (R01 AG009956, P30 AG010133), the Memory and Aging Project (MAP) ( R01 AG17917), Mayo Clinic (MAYO) (R01 AG032990, U01 AG046139, R01 NS080820, RF1 AG051504, P50 AG016574), Mayo Parkinson’s Disease controls (NS039764, NS071674, 5RC2HG005605), University of Miami (R01 AG027944, R01 AG028786, R01 AG019085, IIRG09133827, A2011048), the Multi-Institutional Research in Alzheimer’s Genetic Epidemiology Study (MIRAGE) (R01 AG09029, R01 AG025259), the National Centralized Repository for Alzheimer’s Disease and Related Dementias (NCRAD) (U24 AG021886), the National Institute on Aging Late Onset Alzheimer’s Disease Family Study (NIA- LOAD) (U24 AG056270), the Religious Orders Study (ROS) (P30 AG10161, R01 AG15819), the Texas Alzheimer’s Research and Care Consortium (TARCC) (funded by the Darrell K Royal Texas Alzheimer’s Initiative), Vanderbilt University/Case Western Reserve University (VAN/CWRU) (R01 AG019757, R01 AG021547, R01 AG027944, R01 AG028786, P01 NS026630, and Alzheimer’s Association), the Washington Heights-Inwood Columbia Aging Project (WHICAP) (RF1 AG054023), the University of Washington Families (VA Research Merit Grant, NIA: P50AG005136, R01AG041797, NINDS: R01NS069719), the Columbia University Hispanic Estudio Familiar de Influencia Genetica de Alzheimer (EFIGA) (RF1 AG015473), the University of Toronto (UT) (funded by Wellcome Trust, Medical Research Council, Canadian Institutes of Health Research), and Genetic Differences (GD) (R01 AG007584). The CHARGE cohorts are supported in part by National Heart, Lung, and Blood Institute (NHLBI) infrastructure grant HL105756 (Psaty), RC2HL102419 (Boerwinkle) and the neurology working group is supported by the National Institute on Aging (NIA) R01 grant AG033193.

The CHARGE cohorts participating in the ADSP include the following: Austrian Stroke Prevention Study (ASPS), ASPS-Family study, and the Prospective Dementia Registry-Austria (ASPS/PRODEM-Aus), the Atherosclerosis Risk in Communities (ARIC) Study, the Cardiovascular Health Study (CHS), the Erasmus Rucphen Family Study (ERF), the Framingham Heart Study (FHS), and the Rotterdam Study (RS). ASPS is funded by the Austrian Science Fond (FWF) grant number P20545-P05 and P13180 and the Medical University of Graz. The ASPS-Fam is funded by the Austrian Science Fund (FWF) project I904), the EU Joint Programme – Neurodegenerative Disease Research (JPND) in frame of the BRIDGET project (Austria, Ministry of Science) and the Medical University of Graz and the Steiermärkische Krankenanstalten Gesellschaft. PRODEM-Austria is supported by the Austrian Research Promotion agency (FFG) (Project No. 827462) and by the Austrian National Bank (Anniversary Fund, project 15435. ARIC research is carried out as a collaborative study supported by NHLBI contracts (HHSN268201100005C, HHSN268201100006C, HHSN268201100007C, HHSN268201100008C, HHSN268201100009C, HHSN268201100010C, HHSN268201100011C, and HHSN268201100012C). Neurocognitive data in ARIC is collected by U01 2U01HL096812, 2U01HL096814, 2U01HL096899, 2U01HL096902, 2U01HL096917 from the NIH (NHLBI, NINDS, NIA and NIDCD), and with previous brain MRI examinations funded by R01-HL70825 from the NHLBI. CHS research was supported by contracts HHSN268201200036C, HHSN268200800007C, N01HC55222, N01HC85079, N01HC85080, N01HC85081, N01HC85082, N01HC85083, N01HC85086, and grants U01HL080295 and U01HL130114 from the NHLBI with additional contribution from the National Institute of Neurological Disorders and Stroke (NINDS). Additional support was provided by R01AG023629, R01AG15928, and R01AG20098 from the NIA. FHS research is supported by NHLBI contracts N01-HC-25195, HHSN268201500001I and 75N92019D00031. This study was also supported by additional grants from the NIA (R01s AG054076, AG049607 and AG033040 and NINDS (R01 NS017950). The ERF study as a part of EUROSPAN (European Special Populations Research Network) was supported by European Commission FP6 STRP grant number 018947 (LSHG-CT-2006-01947) and also received funding from the European Community’s Seventh Framework Programme (FP7/2007-2013)/grant agreement HEALTH-F4- 2007-201413 by the European Commission under the programme “Quality of Life and Management of the Living Resources” of 5th Framework Programme (no. QLG2-CT-2002- 01254). High-throughput analysis of the ERF data was supported by a joint grant from the Netherlands Organization for Scientific Research and the Russian Foundation for Basic Research (NWO-RFBR 047.017.043). The Rotterdam Study is funded by Erasmus Medical Center and Erasmus University, Rotterdam, the Netherlands Organization for Health Research and Development (ZonMw), the Research Institute for Diseases in the Elderly (RIDE), the Ministry of Education, Culture and Science, the Ministry for Health, Welfare and Sports, the European Commission (DG XII), and the municipality of Rotterdam. Genetic datasets are also supported by the Netherlands Organization of Scientific Research NWO Investments (175.010.2005.011, 911-03-012), the Genetic Laboratory of the Department of Internal Medicine, Erasmus MC, the Research Institute for Diseases in the Elderly (014-93-015; RIDE2), and the Netherlands Genomics Initiative (NGI)/Netherlands Organization for Scientific Research (NWO) Netherlands Consortium for Healthy Aging (NCHA), project 050-060-810. All studies are grateful to their participants, faculty and staff. The content of these manuscripts is solely the responsibility of the authors and does not necessarily represent the official views of the National Institutes of Health or the U.S. Department of Health and Human Services.

The FUS cohorts include: the Alzheimer’s Disease Research Centers (ADRC) (P30 AG062429, P30 AG066468, P30 AG062421, P30 AG066509, P30 AG066514, P30 AG066530, P30 AG066507, P30 AG066444, P30 AG066518, P30 AG066512, P30 AG066462, P30 AG072979, P30 AG072972, P30 AG072976, P30 AG072975, P30 AG072978, P30 AG072977, P30 AG066519, P30 AG062677, P30 AG079280, P30 AG062422, P30 AG066511, P30 AG072946, P30 AG062715, P30 AG072973, P30 AG066506, P30 AG066508, P30 AG066515, P30 AG072947, P30 AG072931, P30 AG066546, P20 AG068024, P20 AG068053, P20 AG068077, P20 AG068082, P30 AG072958, P30 AG072959), Alzheimer’s Disease Neuroimaging Initiative (ADNI) (U19AG024904), Amish Protective Variant Study (RF1AG058066), Cache County Study (R01AG11380, R01AG031272, R01AG21136, RF1AG054052), Case Western Reserve University Brain Bank (CWRUBB) (P50AG008012), Case Western Reserve University Rapid Decline (CWRURD) (RF1AG058267, NU38CK000480), CubanAmerican Alzheimer’s Disease Initiative (CuAADI) (3U01AG052410), Estudio Familiar de Influencia Genetica en Alzheimer (EFIGA) (5R37AG015473, RF1AG015473, R56AG051876), Genetic and Environmental Risk Factors for Alzheimer Disease Among African Americans Study (GenerAAtions) (2R01AG09029, R01AG025259, 2R01AG048927), Gwangju Alzheimer and Related Dementias Study (GARD) (U01AG062602), Hillblom Aging Network (2014-A-004-NET, R01AG032289, R01AG048234), Hussman Institute for Human Genomics Brain Bank (HIHGBB) (R01AG027944, Alzheimer’s Association “Identification of Rare Variants in Alzheimer Disease”), Ibadan Study of Aging (IBADAN) (5R01AG009956), Longevity Genes Project (LGP) and LonGenity (R01AG042188, R01AG044829, R01AG046949, R01AG057909, R01AG061155, P30AG038072), Mexican Health and Aging Study (MHAS) (R01AG018016), Multi-Institutional Research in Alzheimer’s Genetic Epidemiology (MIRAGE) (2R01AG09029, R01AG025259, 2R01AG048927), Northern Manhattan Study (NOMAS) (R01NS29993), Peru Alzheimer’s Disease Initiative (PeADI) (RF1AG054074), Puerto Rican 1066 (PR1066) (Wellcome Trust (GR066133/GR080002), European Research Council (340755)), Puerto Rican Alzheimer Disease Initiative (PRADI) (RF1AG054074), Reasons for Geographic and Racial Differences in Stroke (REGARDS) (U01NS041588), Research in African American Alzheimer Disease Initiative (REAAADI) (U01AG052410), the Religious Orders Study (ROS) (P30 AG10161, P30 AG72975, R01 AG15819, R01 AG42210), the RUSH Memory and Aging Project (MAP) (R01 AG017917, R01 AG42210Stanford Extreme Phenotypes in AD (R01AG060747), University of Miami Brain Endowment Bank (MBB), University of Miami/Case Western/North Carolina A&T African American (UM/CASE/NCAT) (U01AG052410, R01AG028786), and Wisconsin Registry for Alzheimer’s Prevention (WRAP) (R01AG027161 and R01AG054047).

The four LSACs are: the Human Genome Sequencing Center at the Baylor College of Medicine (U54 HG003273), the Broad Institute Genome Center (U54HG003067), The American Genome Center at the Uniformed Services University of the Health Sciences (U01AG057659), and the Washington University Genome Institute (U54HG003079). Genotyping and sequencing for the ADSP FUS is also conducted at John P. Hussman Institute for Human Genomics (HIHG) Center for Genome Technology (CGT).

Biological samples and associated phenotypic data used in primary data analyses were stored at Study Investigators institutions, and at the National Centralized Repository for Alzheimer’s Disease and Related Dementias (NCRAD, U24AG021886) at Indiana University funded by NIA. Associated Phenotypic Data used in primary and secondary data analyses were provided by Study Investigators, the NIA funded Alzheimer’s Disease Centers (ADCs), and the National Alzheimer’s Coordinating Center (NACC, U24AG072122) and the National Institute on Aging Genetics of Alzheimer’s Disease Data Storage Site (NIAGADS, U24AG041689) at the University of Pennsylvania, funded by NIA. Harmonized phenotypes were provided by the ADSP Phenotype Harmonization Consortium (ADSP-PHC), funded by NIA (U24 AG074855, U01 AG068057 and R01 AG059716) and Ultrascale Machine Learning to Empower Discovery in Alzheimer’s Disease Biobanks (AI4AD, U01 AG068057). This research was supported in part by the Intramural Research Program of the National Institutes of health, National Library of Medicine. Contributors to the Genetic Analysis Data included Study Investigators on projects that were individually funded by NIA, and other NIH institutes, and by private U.S. organizations, or foreign governmental or nongovernmental organizations.

### sa000002 - Alzheimer’s Disease Neuroimaging Initiative (ADNI)

Data collection and sharing for this project was funded by the Alzheimer's Disease Neuroimaging Initiative (ADNI) (National Institutes of Health Grant U01 AG024904) and DOD ADNI (Department of Defense award number W81XWH-12-2-0012). ADNI is funded by the National Institute on Aging, the National Institute of Biomedical Imaging and Bioengineering, and through generous contributions from the following: AbbVie, Alzheimer’s Association; Alzheimer’s Drug Discovery Foundation; Araclon Biotech; BioClinica, Inc.; Biogen; Bristol-Myers Squibb Company; CereSpir, Inc.; Cogstate; Eisai Inc.; Elan Pharmaceuticals, Inc.; Eli Lilly and Company; EuroImmun; F. Hoffmann-La Roche Ltd and its affiliated company Genentech, Inc.; Fujirebio; GE Healthcare; IXICO Ltd.; Janssen Alzheimer Immunotherapy Research & Development, LLC.; Johnson & Johnson Pharmaceutical Research & Development LLC.; Lumosity; Lundbeck; Merck & Co., Inc.; Meso Scale Diagnostics, LLC.; NeuroRx Research; Neurotrack Technologies; Novartis Pharmaceuticals Corporation; Pfizer Inc.; Piramal Imaging; Servier; Takeda Pharmaceutical Company; and Transition Therapeutics. The Canadian Institutes of Health Research is providing funds to support ADNI clinical sites in Canada. Private sector contributions are facilitated by the Foundation for the National Institutes of Health (www.fnih.org). The grantee organization is the Northern California Institute for Research and Education, and the study is coordinated by the Alzheimer’s Therapeutic Research Institute at the University of Southern California. ADNI data are disseminated by the Laboratory for Neuro Imaging at the University of Southern California.

Additional information to include in an acknowledgment statement can be found on the LONI site: https://adni.loni.usc.edu/wp-content/uploads/how_to_apply/ADNI_Data_Use_Agreement.pdf.

sa000003- Alzheimer’s Disease Genetics Consortium: African Americans (ADGC AA)

The Alzheimer’s Disease Genetics Consortium (ADGC) supported sample preparation, whole exome sequencing and data processing through NIA grant U01AG032984. Sequencing data generation and harmonization is supported by the Genome Center for Alzheimer’s Disease, U54AG052427, and data sharing is supported by NIAGADS, U24AG041689. Samples from the National Centralized Repository for Alzheimer’s Disease and Related Dementias (NCRAD), which receives government support under a cooperative agreement grant (U24 AG021886) awarded by the National Institute on Aging (NIA), were used in this study. We thank contributors who collected samples used in this study, as well as patients and their families, whose help and participation made this work possible. NIH grants supported enrollment and data collection for the individual studies including: GenerAAtions R01AG20688 (PI M. Daniele Fallin, PhD); Miami/Duke R01 AG027944, R01 AG028786 (PI Margaret A. Pericak-Vance, PhD); NC A&T P20 MD000546, R01 AG28786-01A1 (PI Goldie S. Byrd, PhD); Case Western (PI Jonathan L. Haines, PhD); MIRAGE R01 AG009029 (PI Lindsay A. Farrer, PhD); ROS P30AG10161, R01AG15819, R01AG30146, TGen (PI David A. Bennett, MD); MAP R01AG17917, R01AG15819, TGen (PI David A. Bennett, MD). The NACC database is funded by NIA/NIH Grant U01 AG016976. NACC data are contributed by the NIA-funded ADCs: P30 AG019610 (PI Eric Reiman, MD), P30 AG013846 (PI Neil Kowall, MD), P30 AG062428-01 (PI James Leverenz, MD) P50 AG008702 (PI Scott Small, MD), P50 AG025688 (PI Allan Levey, MD, PhD), P50 AG047266 (PI Todd Golde, MD, PhD), P30 AG010133 (PI Andrew Saykin, PsyD), P50 AG005146 (PI Marilyn Albert, PhD), P30 AG062421-01 (PI Bradley Hyman, MD, PhD), P30 AG062422-01 (PI Ronald Petersen, MD, PhD), P50 AG005138 (PI Mary Sano, PhD), P30 AG008051 (PI Thomas Wisniewski, MD), P30 AG013854 (PI Robert Vassar, PhD), P30 AG008017 (PI Jeffrey Kaye, MD), P30 AG010161 (PI David Bennett, MD), P50 AG047366 (PI Victor Henderson, MD, MS), P30 AG010129 (PI Charles DeCarli, MD), P50 AG016573 (PI Frank LaFerla, PhD), P30 AG062429-01(PI James Brewer, MD, PhD), P50 AG023501 (PI Bruce Miller, MD), P30 AG035982 (PI Russell Swerdlow, MD), P30 AG028383 (PI Linda Van Eldik, PhD), P30 AG053760 (PI Henry Paulson, MD, PhD), P30 AG010124 (PI John Trojanowski, MD, PhD), P50 AG005133 (PI Oscar Lopez, MD), P50 AG005142 (PI Helena Chui, MD), P30 AG012300 (PI Roger Rosenberg, MD), P30 AG049638 (PI Suzanne Craft, PhD), P50 AG005136 (PI Thomas Grabowski, MD), P30 AG062715-01 (PI Sanjay Asthana, MD, FRCP), P50 AG005681 (PI John Morris, MD), P50 AG047270 (PI Stephen Strittmatter, MD, PhD).

### sa000004 - The Familial Alzheimer Sequencing (FASe) project

This work was supported by grants from the National Institutes of Health (R01AG044546, P01AG003991, RF1AG053303, R01AG058501, U01AG058922, RF1AG058501 and R01AG057777). The recruitment and clinical characterization of research participants at Washington University were supported by NIH P50 AG05681, P01 AG03991, and P01 AG026276. This work was supported by access to equipment made possible by the Hope Center for Neurological Disorders, and the Departments of Neurology and Psychiatry at Washington University School of Medicine.

We thank the contributors who collected samples used in this study, as well as patients and their families, whose help and participation made this work possible. This work was supported by access to equipment made possible by the Hope Center for Neurological Disorders, and the Departments of Neurology and Psychiatry at Washington University School of Medicine

sa000005- Brkanac – Family-based genome scan for AAO of LOAD

This work was partially supported by grant funding from NIH R01 AG039700 and NIH P50 AG005136. Subjects and samples used here were originally collected with grant funding from NIH U24 AG026395, U24 AG021886, P50 AG008702, P01 AG007232, R37 AG015473, P30 AG028377, P50 AG05128, P50 AG16574, P30 AG010133, P50 AG005681, P01 AG003991, U01MH046281, U01 MH046290 and U01 MH046373. The funders had no role in study design, analysis or preparation of the manuscript. The authors declare no competing interests.

sa000006- HIHG Miami Families with AD

This work was supported by the National Institutes of Health (R01 AG027944, R01 AG028786 to MAPV, R01 AG019085 to JLH, P20 MD000546); a joint grant from the Alzheimer’s Association (SG-14-312644) and the Fidelity Biosciences Research Initiative to MAPV; the BrightFocus Foundation (A2011048 to MAPV). NIA-LOAD Family-Based Study supported the collection of samples used in this study through NIH grants U24 AG026395 and R01 AG041797 and the MIRAGE cohort was supported through the NIH grants R01 AG025259 and R01 AG048927. We thank contributors, including the Alzheimer’s disease Centers who collected samples used in this study, as well as patients and their families, whose help and participation made this work possible. Study design: HNC, BWK, JLH, MAPV; Sample collection: MLC, JMV, RMC, LAF, JLH, MAPV; Whole exome sequencing and Sanger sequencing: SR, PLW; Sequencing data analysis: HNC, BWK, KLHN, SR, MAK, JRG, ERM, GWB, MAPV; Statistical analysis: BWK, KLHN, JMJ, MAPV; Preparation of manuscript: HNC, BWK. The authors jointly discussed the experimental results throughout the duration of the study. All authors read and approved the final manuscript.

sa000007- Washington Heights/Inwood Columbia Aging Project (WHICAP)

Data collection and sharing for this project was supported by the Washington Heights-Inwood Columbia Aging Project (WHICAP, PO1AG07232, R01AG037212, RF1AG054023) funded by the National Institute on Aging (NIA) and by the National Center for Advancing Translational Sciences, National Institutes of Health, through Grant Number UL1TR001873. This manuscript has been reviewed by WHICAP investigators for scientific content and consistency of data interpretation with previous WHICAP Study publications. We acknowledge the WHICAP study participants and the WHICAP research and support staff for their contributions to this study.

sa000008- Charles F. and Joanne Knight Alzheimer’s Disease Research Center (Knight ADRC)

This work was supported by grants from the National Institutes of Health (R01AG044546, P01AG003991, RF1AG053303, R01AG058501, U01AG058922, RF1AG058501 and R01AG057777). The recruitment and clinical characterization of research participants at Washington University were supported by NIH P50 AG05681, P01 AG03991, and P01 AG026276. This work was supported by access to equipment made possible by the Hope Center for Neurological Disorders, and the Departments of Neurology and Psychiatry at Washington University School of Medicine.

We thank the contributors who collected samples used in this study, as well as patients and their families, whose help and participation made this work possible. This work was supported by access to equipment made possible by the Hope Center for Neurological Disorders, and the Departments of Neurology and Psychiatry at Washington University School of Medicine.

sa000009- Corticobasal degeneration Study (CBD)

CBD Solutions funded the WES, data processing, and analysis. Assembled samples are from University College London (John Hardy), Mayo Clinic Jacksonville (Dennis Dickson), University of Pennsylvania (John Trojanowski), Emory University (Marla Gearing), Johns Hopkins University (Alex Pantelyat), Indiana University (Bernadino Ghetti), New York Brain Bank (Jean Paul Vonsattel), McClean Brain Bank (Elaine Benes), University of Texas Southwestern (Charles White), University of California Los Angeles (William Tourtelloute), and European collaborators at University Munich and Neurobiobank Munich (Gunter Hoglinger, Ulrich Muller, Hans Kretzschmr), Newcastle University, University of Barcelona (Charles Gaig), MRC London Brain Bank, Australian Brain Bank, and the University of Madrid (Alberto Rábano Gutiérrez).

sa000010- Progressive Supranuclear Palsy Study (PSP)

This work was funded by the following NIH grants: P01 AG017586 (VM-YL, GDS, JQT), U54 NS100693 (OR, DD, GDS), UG3 NS104095 (GDS, L-SW, OR), U54 AG052427 (l-SW, GDS), P30 AG010133 (B.G.), R01 AG057516 (AC, AM, AW, JAP, SG), R01 HL143790 (AC, SG), R01 HG010067 (SG), RF1 AG055477 (CB), P01 AG017586 (VM-YL, GDS, JQT, VMV), UG3 NS104095 and CWOW grant U54 NS100693 (DD), AG025688 and NS055077 (MG), P30 AG012300 (CLW), P30 AG053760 (APL and RA), 1P50NS091856 (RA), 5 P50 AG005134 (MPF), AG005131 (DRG), Johns Hopkins University Morris K. Udall Parkinson’s Disease Research Center of Excellence grant P50 NS038377 and Alzheimer’s Disease Research Center grant P50 AG05146 (JCT), U24 NS072026 and P30 AG19610 (TGB). This work was also funded by Cure PSP (GDS), the Rainwater Foundation (GDS), the Daniel B. Burke Endowed Chair for Diabetes Research (SG), the CHOP Center for Spatial and Functional Genomics (AW, SFG), a CUREPSP research grant (Cure PSP Grant # 515-14; 2013-2015) to P.P., the Reta Lila Weston Trust for Medical Research, the PSP Association (RdS), and the Michael J. Fox Foundation for Parkinson’s Research (TGB). G. Höglinger was funded by the German Research Foundation (DFG) under Germany’s Excellence Strategy within the framework of the Munich Cluster for Systems Neurology (EXC 2145 SyNergy – ID 390857198), the German Federal Ministry of Education and Research (BMBF, 01KU1403A EpiPD; 01EK1605A HitTau), and the NOMIS foundation (FTLD project). J. Hardy was partly funded by UKDRI limited which receives its funding from the MRC, the Alzheimer’s Society and Alzheimer Research UK. The London Neurodegenerative Diseases Brain Bank receives funding from the UK Medical Research Council (MR/L016397/1) and as part of the Brains for Dementia Research programme, jointly funded by Alzheimer’s Research UK and the Alzheimer’s Society. Queen Square Brain Bank is supported by the Reta Lila Weston Institute for Neurological Studies and the Medical Research Council UK. Newcastle Brain Tissue Resource is funded in part by a grant from the UK Medical Research Council (MR/L016451/1) and by Brains for Dementia Research, a joint venture between Alzheimer’s Society and Alzheimer’s Research UK (CMM) and National Institute of Health Research Biomedical Research Centre at Newcastle upon Tyne Hospitals NHS Foundation Trust and Newcastle University (CMM). This work was partly funded by UKDRI limited which receives its funding from the MRC, the Alzheimer’s Society and Alzheimer Research UK (JH). The Mayo Clinic Florida had support from a Morris K. Udall Parkinson’s Disease Research Center of Excellence (NINDS P50 #NS072187), CurePSP and the Tau Consortium. OAR is supported by a NINDS Tau Center without Walls (U54-NS100693), NINDS R01-NS078086 and the Mayo Clinic Center for Individualized Medicine. Funding provided by CurePSP through the generous support of the Peebler PSP Research Foundation in memory of Charles D. Peebler Jr. and Drs. Jeffrey S. and Jennifer R. Friedman in memory of Morton L. Friedman.

### sa000011 - Accelerating Medicines Partnership- Alzheimer’s Disease (AMP-AD)

Mayo RNAseq Study- Study data were provided by the following sources: The Mayo Clinic Alzheimer's Disease Genetic Studies, led by Dr. Nilufer Ertekin-Taner and Dr. Steven G. Younkin, Mayo Clinic, Jacksonville, FL using samples from the Mayo Clinic Study of Aging, the Mayo Clinic Alzheimer's Disease Research Center, and the Mayo Clinic Brain Bank. Data collection was supported through funding by NIA grants P50 AG016574, R01 AG032990, U01 AG046139, R01 AG018023, U01 AG006576, U01 AG006786, R01 AG025711, R01 AG017216, R01 AG003949, NINDS grant R01 NS080820, CurePSP Foundation, and support from Mayo Foundation. Study data includes samples collected through the Sun Health Research Institute Brain and Body Donation Program of Sun City, Arizona. The Brain and Body Donation Program is supported by the National Institute of Neurological Disorders and Stroke (U24 NS072026 National Brain and Tissue Resource for Parkinson's Disease and Related Disorders), the National Institute on Aging (P30 AG19610 Arizona Alzheimer's Disease Core Center), the Arizona Department of Health Services (contract 211002, Arizona Alzheimer's Research Center), the Arizona Biomedical Research Commission (contracts 4001, 0011, 05-901 and 1001 to the Arizona Parkinson's Disease Consortium) and the Michael J. Fox Foundation for Parkinson's Research

ROSMAP- We are grateful to the participants in the Religious Order Study, the Memory and Aging Project. This work is supported by the US National Institutes of Health [U01 AG046152, R01 AG043617, R01 AG042210, R01 AG036042, R01 AG036836, R01 AG032990, R01 AG18023, RC2 AG036547, P50 AG016574, U01 ES017155, KL2 RR024151, K25 AG041906-01, R01 AG30146, P30 AG10161, R01 AG17917, R01 AG15819, K08 AG034290, P30 AG10161 and R01 AG11101.

Mount Sinai Brain Bank (MSBB)- This work was supported by the grants R01AG046170, RF1AG054014, RF1AG057440 and R01AG057907 from the NIH/National Institute on Aging (NIA). R01AG046170 is a component of the AMP-AD Target Discovery and Preclinical Validation Project. Brain tissue collection and characterization was supported by NIH HHSN271201300031C.

### sa000012 - [University of Pittsburgh- Kamboh WGS](https://dss.niagads.org/studies/sa000012/)

This study was supported by the National Institute on Aging (NIA) grants AG030653, AG041718, AG064877 and P30-AG066468.

### sa000013 - NACC Genentech WGS

We would like to thank study participants, their families, and the sample collectors for their invaluable contributions. This research was supported in part by the National Institute on Aging grant U01AG049508 (PI Alison M. Goate). This research was supported in part by Genentech, Inc. (PI Alison M. Goate, Robert R. Graham).

The NACC database is funded by NIA/NIH Grant U01 AG016976. NACC data are contributed by these NIA-funded ADCs: P30 AG013846 (PI Neil Kowall, MD), P50 AG008702 (PI Scott Small, MD), P50 AG025688 (PI Allan Levey, MD, PhD), P30 AG010133 (PI Andrew Saykin, PsyD), P50 AG005146 (PI Marilyn Albert, PhD), P50 AG005134 (PI Bradley Hyman, MD, PhD), P50 AG016574 (PI Ronald Petersen, MD, PhD), P30 AG013854 (PI M. Marsel Mesulam, MD), P30 AG008017 (PI Jeffrey Kaye, MD), P30 AG010161 (PI David Bennett, MD), P30 AG010129 (PI Charles DeCarli, MD), P50 AG016573 (PI Frank LaFerla, PhD), P50 AG005131 (PI Douglas Galasko, MD), P30 AG028383 (PI Linda Van Eldik, PhD), P30 AG010124 (PI John Trojanowski, MD, PhD), P50 AG005142 (PI Helena Chui, MD), P30 AG012300 (PI Roger Rosenberg, MD), P50 AG005136 (PI Thomas Grabowski, MD), P50 AG005681 (PI John Morris, MD), P30 AG028377 (Kathleen Welsh-Bohmer, PhD), and P50 AG008671 (PI Henry Paulson, MD, PhD).

Samples from the National Cell Repository for Alzheimer’s Disease (NCRAD), which receives government support under a cooperative agreement grant (U24 AG21886) awarded by the National Institute on Aging (NIA), were used in this study. We thank contributors who collected samples used in this study, as well as patients and their families, whose help and participation made this work possible.

The Alzheimer's Disease Genetics Consortium supported the collection of samples used in this study through National Institute on Aging (NIA) grants U01AG032984 and RC2AG036528.

### sa000014 - Cache County Study

We acknowledge the generous contributions of the Cache County Memory Study participants. Sequencing for this study was funded by RF1AG054052 (PI: John S.K. Kauwe).

### sa000015 - NIH, CurePSP and Tau Consortium PSP WGS

This project was funded by the NIH grant UG3NS104095 and supported by grants U54NS100693 and U54AG052427. Queen Square Brain Bank is supported by the Reta Lila Weston Institute for Neurological Studies and the Medical Research Council UK. The Mayo Clinic Florida had support from a Morris K. Udall Parkinson's Disease Research Center of Excellence (NINDS P50 #NS072187), CurePSP and the Tau Consortium. The samples from the University of Pennsylvania are supported by NIA grant P01AG017586.

### sa000016 - CurePSP and Tau Consortium PSP WGS

This project was funded by the Tau Consortium, Rainwater Charitable Foundation, and CurePSP. It was also supported by NINDS grant U54NS100693 and NIA grants U54NS100693 and U54AG052427. Queen Square Brain Bank is supported by the Reta Lila Weston Institute for Neurological Studies and the Medical Research Council UK. The Mayo Clinic Florida had support from a Morris K. Udall Parkinson's Disease Research Center of Excellence (NINDS P50 #NS072187), CurePSP and the Tau Consortium. The samples from the University of Pennsylvania are supported by NIA grant P01AG017586. Tissues were received from the Victorian Brain Bank, supported by The Florey Institute of Neuroscience and Mental Health, The Alfred and the Victorian Forensic Institute of Medicine and funded in part by Parkinson’s Victoria and MND Victoria. We are grateful to the Sun Health Research Institute Brain and Body Donation Program of Sun City, Arizona for the provision of human biological materials (or specific description, e.g. brain tissue, cerebrospinal fluid). The Brain and Body Donation Program is supported by the National Institute of Neurological Disorders and Stroke (U24 NS072026 National Brain and Tissue Resource for Parkinson's Disease and Related Disorders), the National Institute on Aging (P30 AG19610 Arizona Alzheimer's Disease Core Center), the Arizona Department of Health Services ( contract 211002, Arizona Alzheimer's Research Center), the Arizona Biomedical Research Commission (contracts 4001, 0011, 05-901 and 1001 to the Arizona Parkinson's Disease Consortium) and the Michael J. Fox Foundation for Parkinson's Research. Biomaterial was provided by the Study Group DESCRIBE of theClinical Research of the German Center for Neurodegenerative Diseases (DZNE).

### sa000017 - UCLA Progressive Supranuclear Palsy

If data are used for a publication, “on behalf of the AL-108-231 investigators” should be included in the authorship list.

ADSP Consortium Contributors

**University of Texas Houston**

Eric Boerwinkle

Myriam Fornage

Jennifer E. Below

Xueqiu Jian

**Boston University CHARGE**

Sudha Seshadri

Anita L. DeStefano

Adrienne Cupples

Honghuang Lin

Claudia Satizabal

Josée Dupuis

Alexa S Beiser

Chloe Sarnowski

Yuning Chen

Ching Ti Liu

Fangui Jenny Sun

**Broad Institute**

Seung Hoan Choi

**University of Washington**

Ellen M. Wijsman

Bruce M. Psaty

Joshua C. Bis

Daniela Witten

Kenneth Rice

Timothy Thornton

Elizabeth Blue

Patrick A Navas

Michael O. Dorschner

Debby Tsuang

Mohamad Saad

Alejandro Q. Nato Jr.

Rafael A. Nafikov

Andrea R Horimoto

Harkirat Sohi

Lisa Brown

Hiep Nguyen

Bowen Wang

Tyler Day

**Erasmus Medical University/Rotterdam**

Cornelia van Duijn

Shahzad Ahmad

Najaf Amin

M Afran Ikram

Sven van der Lee

Ashley Vanderspek

**Medical University Graz, Austria**

Helena Schmidt

Reinhold Schmidt

**Mount Sinai School of Medicine**

Alison M Goate

Alan Renton

Manav Kapoor

Edoardo Marcora

**Washington University**

Carlos Cruchaga

**University of Pennsylvania**

Adam C Naj

Gerard D Schellenberg

John Malamon

Weixin Wang

Nancy Zhang

Laura Cantwell

Yuk Yee Leung

Elisabeth Mlynarski

**Stanford University**

Li Charlie Xia

**University of Miami**

Brian W. Kunkle

Eden R Martin

James Jaworski

Gary W Beecham

Margaret A Pericak-Vance

Kara L Hamilton-Nelson

Michael Schmidt

Farid Rajabli

Jeffery M Vance

Michael L Cuccaro

Pedro Mena

Larry D Adams

**Columbia University**

Badri Vardarajan

Christiane Reitz

Giuseppe Tosto

Dolly Reyes-Dumeyer

Sandra Barral

Richard Mayeux

Phillip L De Jager

**Case Western Reserve University**

Jonathan L Haines

Yeunjoo Song

William S. Bush

Penelope Benchek

Nicholas Wheeler

**Boston University CASA**

Lindsay A Farrer

Kathryn L. Lunetta

Xiaoling Zhang

Dan Lancour

John Farrell

Yiyi Ma

Devanshi Patel

Jaeyoon Chung

Gyungah R Jun

Congcong Zhu

Jesse Mez

**Baylor College of Medicine**

Shannon Dugan-Perez

Simon White

Divya Kalra

Waleed Nasser

Viktoriya Korchina

Yue Liu

Sandra Lee

Adam English

Evette Skinner

Xiuping Liu

Joy Jayaseelan

Huyen Dinh

Yi Han

Jireh Santibanez

Ziad Khan

Harsha Doddapeneni

Jianhong Hu

William Salerno

Kim C. Worley

Donna Muzny

Richard A. Gibbs

Michelle Bellair

Yiming Zhu

**Broad Institute**

Namrata Gupta

Stacey Gabriel

Eric Banks

**Washington University St. Louis**

Robert S. Fulton

Jason Waligorski

Elizabeth Appelbaum

Lucinda Antonacci-Fulton

Richard K. Wilson

Daniel C. Koboldt

David E. Larson

**Indiana University**

Kelley M Faber

Tatiana M Foroud

**University of Pennslyvania**

Li-San Wang

Amanda B Kuzma

Otto Valladares

Prabhakaran Gangadharan

Yi Zhao

Liming Qu

Yi-Fan Chou

Han-Jen Lin

Briana M Vogel

Heather A Issen

**NIA**

Lenore J. Launer

**Rush University**

David A Bennett

**UT/Baylor**

Jan Bressler

Xiaoming Liu

**NCBI**

Adam Stine

Michael Feolo

**ARIC**

Thomas H. Mosley
